# Proton magnetic resonance spectroscopy of N-acetyl aspartate in first depressive episode and chronic major depressive disorder: a systematic review and meta-analysis

**DOI:** 10.1101/2022.07.01.22277107

**Authors:** Luigi F. Saccaro, Matteo Tassone, Francesca Tozzi, Grazia Rutigliano

## Abstract

N-acetyl aspartate (NAA) is a marker of neuronal integrity and metabolism. Deficiency in neuronal plasticity and hypometabolism are implicated in the pathophysiology of Major Depressive Disorder (MDD). To test if cerebral NAA concentrations decrease progressively over the MDD course, we conducted a meta-analysis of Proton Magnetic Resonance Spectroscopy (^1^H-MRS) studies comparing NAA concentrations in chronic MDD (cMDD) and first episode of depression (FED) to healthy controls. We searched Scopus® and Web of Knowledge □ using search terms related to depression and NAA. Hedges’ g was used as effect size measure, together with heterogeneity analyses, test of moderators and publication bias and quality assessment. The protocol is registered in PROSPERO (CRD42020221050). Sixty-two studies were included and meta-analyzed using a random-effect model for each brain region. NAA concentrations were significantly reduced in cMDD compared to healthy controls within the frontal lobe (n=26, Hedges’ g= -0.330, 95% CI -0.598 to -0.062; p= 0.018), the occipital lobe (n = 4, Hedges’ g= -0.677, 95% CI -1.013 to -0.341; p = 0.007), the thalamus (n= 4, Hedges’ g= -0.673, 95% CI -1.108 to -0.238; p = 0.016) and the frontal (n = 6, Hedges’ g= -0.471, 95% CI -0.891 to -0.052; p= 0.034) and periventricular white matter (n= 3, Hedges’ g= -0.478, 95% CI -0.938 to -0.018; p= 0.047). We highlighted a gap of knowledge regarding NAA levels in FED. Sensitivity analyses indicated that antidepressant treatment may reverse NAA alterations in the frontal lobe. Our findings are in line with previous evidence showing alterations in the aforementioned brain areas in MDD. Future studies should assess NAA alterations in the early stages of the illness and their longitudinal progression, also considering our preliminary results on the modifying effect of antidepressant treatment.

## 1. Introduction

Major Depressive Disorder (MDD) is a prevalent and serious mood disorder. Accounting for 10% of the total non-fatal disease burden worldwide and affecting more than 300 million people, MDD is globally responsible for more years lost to disability than any other disease according to the World Health Organization (WHO), and is a major contributor to deaths by suicide(1). In MDD, a malfunction of multiple brain areas, including the limbic system and the prefrontal cortex, has been postulated, but, despite significant advances, its pathophysiology and the molecular basis of treatments are still poorly understood. This is further complicated by the heterogeneity of disease phases, as clinical(2) and radiological(3) markers differ between First Episode of Depression (FED) and chronic MDD (cMDD). Differentiating MDD stages has important implications for patient care and clinical research(4). Current treatments for depression do not effectively or sufficiently reduce the associated morbidity and mortality(5). Indeed, up to 50% of individuals treated with antidepressant medications for MDD do not achieve full remission(6). These data highlight the need for better pathophysiologic insight in MDD, as well as for diagnostic and prognostic markers.

Proton Magnetic Resonance Spectroscopy (^1^H-MRS) is a non-invasive technique that allows the in vivo measurement of biochemical changes in the brain, to produce a regionally specific molecular fingerprint. Recent technological advances allow higher signal-to-noise ratios than ever before and finer metabolite analysis. ^1^H-MRS is thus becoming increasingly clinically relevant, contributing to our understanding of psychiatric disorders, such as schizophrenia(7), bipolar disorder(8), and anxiety(9).

N-acetyl aspartate (NAA) is the main peak in the ^1^H-MRS spectra, which makes it easily measured, also at lower and more available field strengths(7, 10). As NAA takes part in lipid biosynthesis, including myelin, it is believed to be a marker of viable neuronal tissue, neuronal health, and neuronal energy metabolism(11). In fact, a permanent NAA decrease is observed in cerebral stroke(12) and neurodegenerative diseases(13), while a transient decrease is observed in acute demyelinating diseases(14) and in the ischemic penumbra(15). Accordingly, cerebral NAA concentrations positively correlate with other parameters of neuronal metabolism in animal models and humans(16-18), and an NAA reduction is associated with neuronal loss or damage(19), or lower neuronal metabolic function(7). For instance, inhibition of mitochondrial complex one decreases NAA mitochondrial production in vitro (7, 20). While NAA may be a marker of several processes besides neuronal metabolism, it is noteworthy that cerebral hypometabolism is thought to be an important player in MDD pathophysiology, potentially underlying MDD symptoms. Evidence for this includes a metanalysis of Positron Emission Tomography (PET) studies on 188 MDD patients and 169 healthy controls (HC) showing reduced metabolism in multiple brain regions in MDD patients(21). Thus, NAA has been studied in MDD as a marker of brain regions (dys)function or metabolism.

To the best of our knowledge, the two most recent and comprehensive meta-analysis of ^1^H-MRS in MDD, involving multiple brain regions and metabolites, were published in 2015(22) and 2006(23), and did not report detectable changes in NAA levels. However, they included only 11(22) and 14(23) studies, respectively.

Our meta-analysis aims at comparing NAA levels, measured in every brain region using ^1^H-MRS, between patients with a diagnosis of MDD (FED or cMDD) and HC. We hypothesize that cerebral NAA concentrations should be lower in MDD patients relative to HC. Our hypothesis is that NAA reduction will be larger in cMDD as compared to FED, in accordance with progressive neuronal damage or hypometabolic changes over the course of the illness(3).

## 2. Materials and methods

### 2.1 Information sources, search strategy, and selection criteria

This Systematic Review was conducted following the Preferred Reporting Items for Systematic Reviews and Meta-analyses (PRISMA) guidelines (**Supplementary Table 1**)(24), and is pre-registered (PROSPERO: CRD42020221050).

We used a two-step approach to identify articles assessing NAA concentration in MDD patients and HC using ^1^H-MRS.

First, we performed an automatic search of two electronic databases: a) Scopus® (www.scopus.com/) Advanced Search, with the following search formula: *“TITLE-ABS-KEY (n-acetylaspartate) OR TITLE-ABS-KEY (naa) AND TITLE-ABS-KEY (major AND depressive AND disorder) OR TITLE-ABS-KEY (mdd) OR TITLE-ABS-KEY (depression)”;* b) Web of Knowledge □ database by Thomas Reuters ® (including Web of Science™, Current Contents Connect, Data Citation Index, Derwent Innovation Index KCI-Korean Journal Database, MEDLINE®, Russian Science Citation Index, SciELO Citation Index, www.webofknowledge.com/), with the following search formula: “TS=(n-acetylaspartate OR NAA) AND TS=(Major depressive disorder OR depression OR MDD)”. The search was extended until the 15^th^ of October 2021. In the second step, we conducted a manual search of the reference lists of all retrieved articles to check for studies potentially missing in the first step. Duplicate references were removed manually. The identified articles were first screened by title and abstract, and the full texts of surviving articles were further inspected for eligibility against the a priori-defined inclusion and exclusion criteria. Candidate articles were independently screened and scrutinized by MT and FT. Discrepancies in study selection were resolved by discussion with an independent arbiter (GR).

We included original articles written in English that employed ^1^H-MRS to compare brain levels of NAA between adult (>18 years old) patients with a diagnosis of MDD (as assessed by DSM, ICD, or consensus expert evaluation) and HC and reported enough data to compute effect sizes. We excluded thus studies focusing on children, adolescents, or patients with diagnoses of any other mental disorder, or that compared MDD patients to patients with diagnoses of any other mental disorder and not with HC. We included cross-sectional and randomized controlled trials (RCT), excluding, for instance, case studies, case series, pilot studies, and reviews. In the case of RCT, we used the NAA measures prior to treatment allocation. Our outcome was NAA concentration in different brain regions (both absolute or scaled to creatine). The Participants, Interventions, Comparators, Outcomes, and Study design (PICOS) criteria are detailed in **Supplementary Table 2**, and the selection in the PRISMA flow diagram (**Figure 1**).

**Figure 1.**
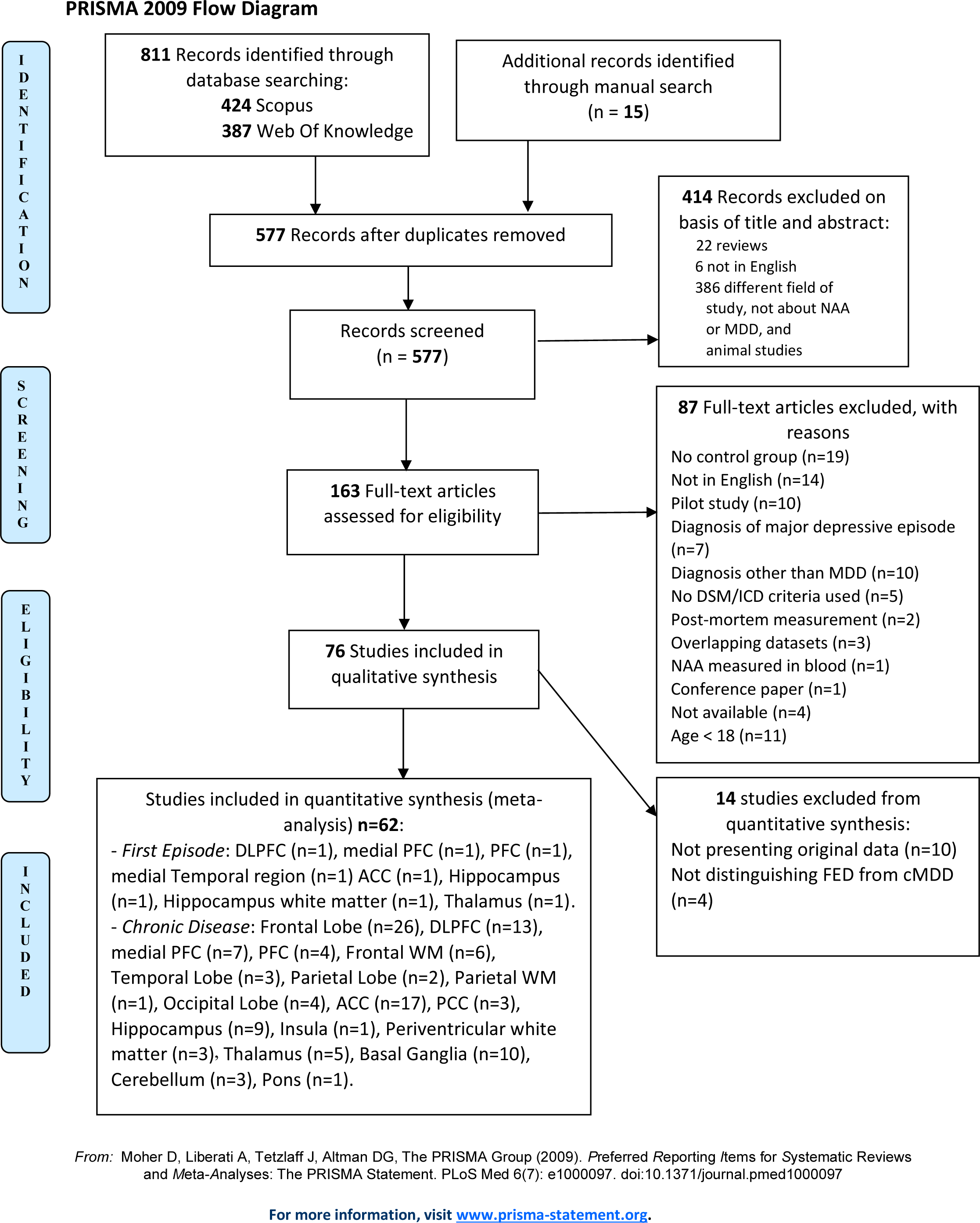
Preferred reporting items for systematic reviews and meta-analyses diagram for study search, 2009. ACC, anterior cingulate cortex; cMDD, chronic major depressive disorder; DLPFC, dorsolateral prefrontal cortex; DSM, diagnostic and statistical manual of mental disorders; FED, first episode of depression; ICD, international classification of diseases; MDD, major depressive disorder; NAA, n-acetyl aspartate; PCC, posterior cingulate cortex; PFC, prefrontal cortex; WM, white matter.

### 2.2. Data extraction

Data were independently extracted by MT and FT. The extracted data were cross-checked, and discrepancies were resolved by discussion between MT, FT, and the independent arbiter (GR).

We extracted sample size, mean NAA concentration and standard deviation(25) or standard error of the mean(26) for MDD patient and HC groups. If the normality assumption allowed parametric statistics in the original paper, t-test or p-value were extracted alongside with direction of the effect size (**Supplementary Methods**).

Where available, we extracted values for: the frontal lobe, including dorsolateral prefrontal cortex (DLPFC), prefrontal cortex (PFC), medial prefrontal cortex (mPFC), and frontal white matter (FWM); the parietal lobe, including parietal white matter (PWM); the temporal lobe, including medial temporal region; the occipital lobe; the limbic lobe, including anterior cingulate cortex (ACC), posterior cingulate cortex (PCC), hippocampus and amygdala; the insular cortex; subcortical structures, such as the thalamus and basal ganglia (BG); the cerebellum; the brain stem; and the periventricular white matter (PVWM). Studies reporting voxels in the striatum, putamen, caudate or lentiform nucleus were counted with studies reporting voxels in the BG and analyzed together. When data from bilateral lobes were reported separately, data from the left lobe was used as the left lobe is examined in most studies(27). We also extracted publication year, information about ^1^H-MRS technique such as field strength, acquisition sequence, echo time (TE) and relaxation time(28), NAA quantification (Cr scaling, vs “absolute” concentration, i.e. relative to tissue water), evidence of correction for cerebrospinal fluid (CSF) partial volume (yes vs no), age of patients and controls, percentage of female subjects, any psychoactive therapy (antidepressants, benzodiazepines, mood stabilizers, and antipsychotics), values of the available depression score such as Hamilton Depression Rating Scale 17,21 or 24 items (HAMD 17, 21 or 24), Montgomery-Asberg Depression Rating Scale (MADRS) and Beck Inventory Scale (BDI). When patient or control groups were split into separately presented subgroups, values were pooled to provide a single value for the entire patient and control groups, using the supplementary formula in **Supplementary Table3**.

### 2.3. Quality assessment

The quality of the selected studies was assessed independently by two reviewers (MT and FT) with the Newcastle-Ottawa Scale (NOS)(29). Studies were evaluated using NOS considering three aspects: patient selection, comparability, and exposure (**Supplementary Table 14**).

### 2.4. Statistical analysis

#### 2.4.1. Main analysis

We performed the meta-analysis using the *meta*(30), *metaphor*(31) and *dmetar*(32) packages in R (version 4.0.5), following the guide in Harrer et al.(33). We ran separate meta-analyses by brain region and illness stage. Studies pooling data of illness phases and/or brain regions were excluded from the analyses if data was inseparable. Effect sizes were pooled using a random-effect model to account for sources of heterogeneity in the combined analysis. Where studies reported more than one NAA quantification method (e.g. Cr scaling and absolute concentration), we selected Cr scaling for analysis.

We report the main outcome, i.e. differences in NAA levels between MDD patients and HC, as Hedge’s g with a significance threshold of p < 0.05(34). Results were visualized through forest plots and tables. We assessed between-study heterogeneity using *Cochran’s Q statistics*(34) and quantified total variability by the *I*^*2*^ *index*(35). We identified outliers and ran sensitivity analyses without them. Through influence analyses, we detected studies with a large impact on the pooled effect size. We employed the Graphic Display of Heterogeneity (GOSH) plots(36) to explore heterogeneity when at least nine studies were available. To assess the robustness of the results, we performed sensitivity analyses by sequentially removing each study and re-running the analysis(31).

#### 2.4.2. Subgroup analyses and meta-regressions

The impact of categorical moderators was evaluated through subgroup analyses. A-priori defined categorical moderators were: ^1^H-MRS acquisition sequence; NAA quantification (Cr scaling vs absolute concentration); cerebrospinal fluid (CSF) quantification (yes vs no); antidepressant treatment (yes vs no); and field strength (>1.5 T vs <1.5 T). Subgroup analyses were conducted where each subgroup had at least 4 studies(34), using the fixed-effects plural model.

For groups with 10 or more studies, we fitted meta-regression models to investigate the influence of pre-defined continuous moderators: year of publication, age, female percentage, ^1^H-MRS field strength, TE, time relaxation, illness duration, and HAMD scores.

We conducted supplementary sensitivity analyses to explore the impact of geriatric populations (> 65 years old) on the meta-analytical estimates. Because of numerical constraints, this was possible only in five brain regions in cMDD patients: medial prefrontal region, anterior cingulate, hippocampus, BG, and FWM.

#### 2.4.3. Publication bias

We tested publication bias using the P-Curve Analysis and the Small Sample Bias Method. We plotted p-curves and funnel plots. For groups with 10 or more studies, we quantitatively assessed publication bias using Egger’s Test(37). When Egger’s test was significant, we used the Duval and Tweedie Trim-and-Fill procedure to estimate true effects controlling for any detected bias(38).

## 3. Results

As described in the PRISMA flowchart (**Figure 1)**, we identified 811 records through database searching (424 from Scopus and 387 from WOS) and 15 records through manual search. After duplicates were removed, we screened 577 records and we excluded 414 records based on title and abstract, so that 163 full-text articles were assessed for eligibility. We excluded 87 full-text articles, leaving 76 studies eligible for the meta-analysis. We excluded 10 articles because they did not present enough data, and data could not be retrieved despite attempts to contact the authors(39-48). Four articles were excluded(49-52) because they did not distinguish FED from cMDD patients in the analyses. Thus, 62 studies were included in 15 separate meta-analysis, according to illness phase and cerebral region(28, 52-113).

The NOS score ranged from 2 to 6 and the mean was 5.43, which suggests that the quality of the included studies was good on average (**Supplementary Table 14**).

### 3.1. Studies characteristics

Characteristics of the included studies are presented in **Table 1**. We included 62 studies: 37 studies measured NAA levels as Cr scaling, the rest (n=25) as absolute concentration. 29 studies were performed at a magnetic field strength of 1.5T, 29 studies at 3T, 2 studies at 7T, 1 at 4T and 1 at 2.1T. MRI protocols and methodological information, including measurement technique and parameters, for each study are described in **Table 1**.

**Table 1.**
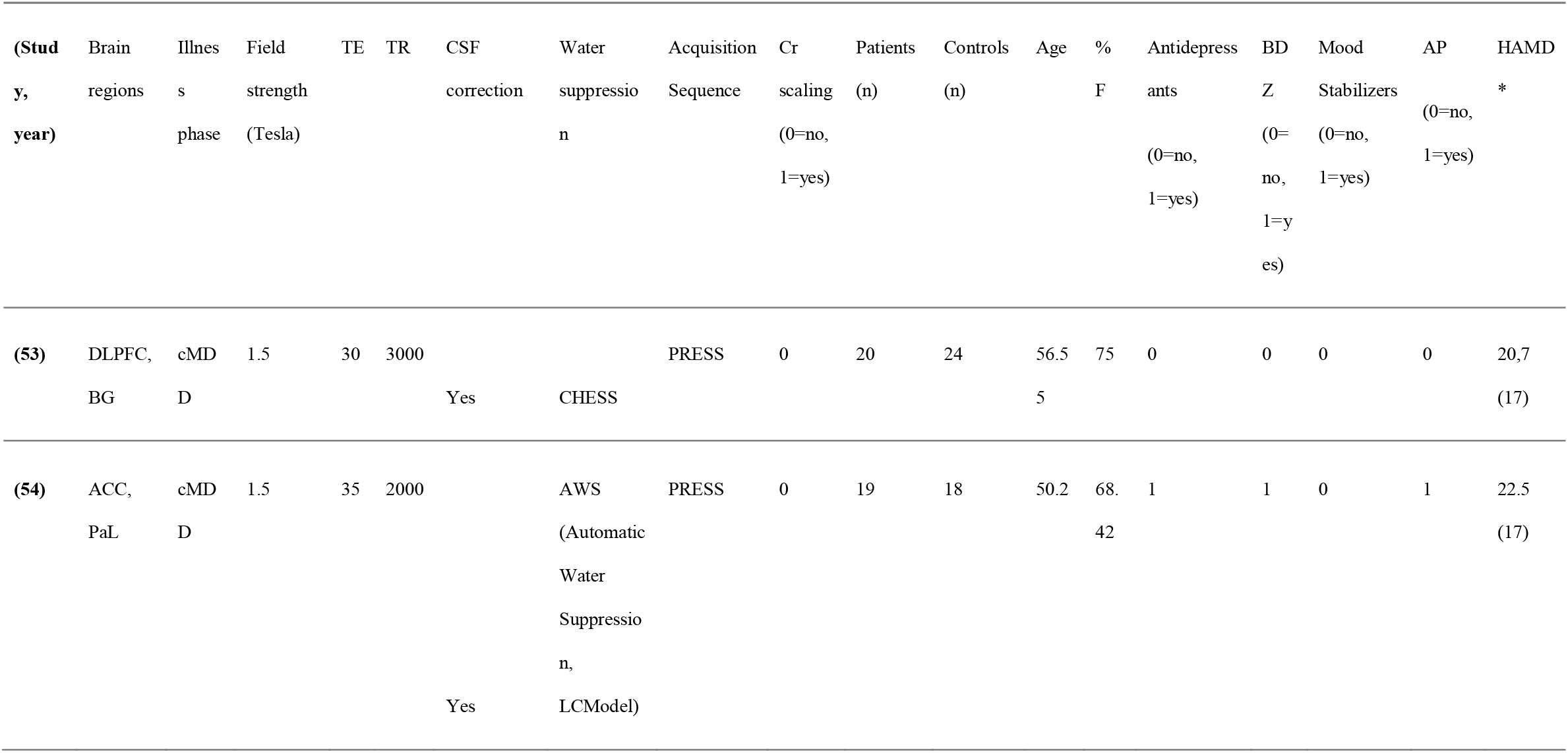
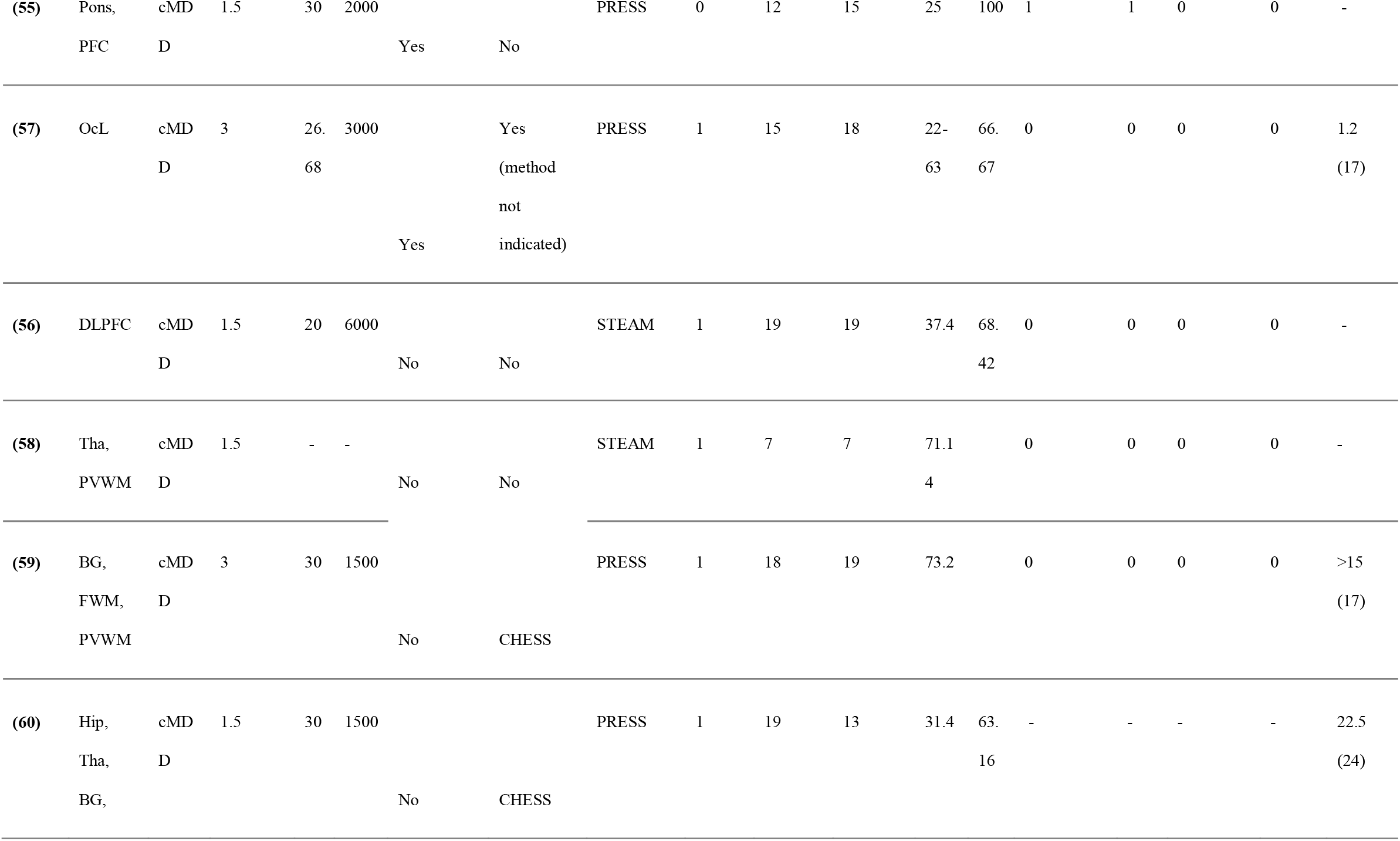
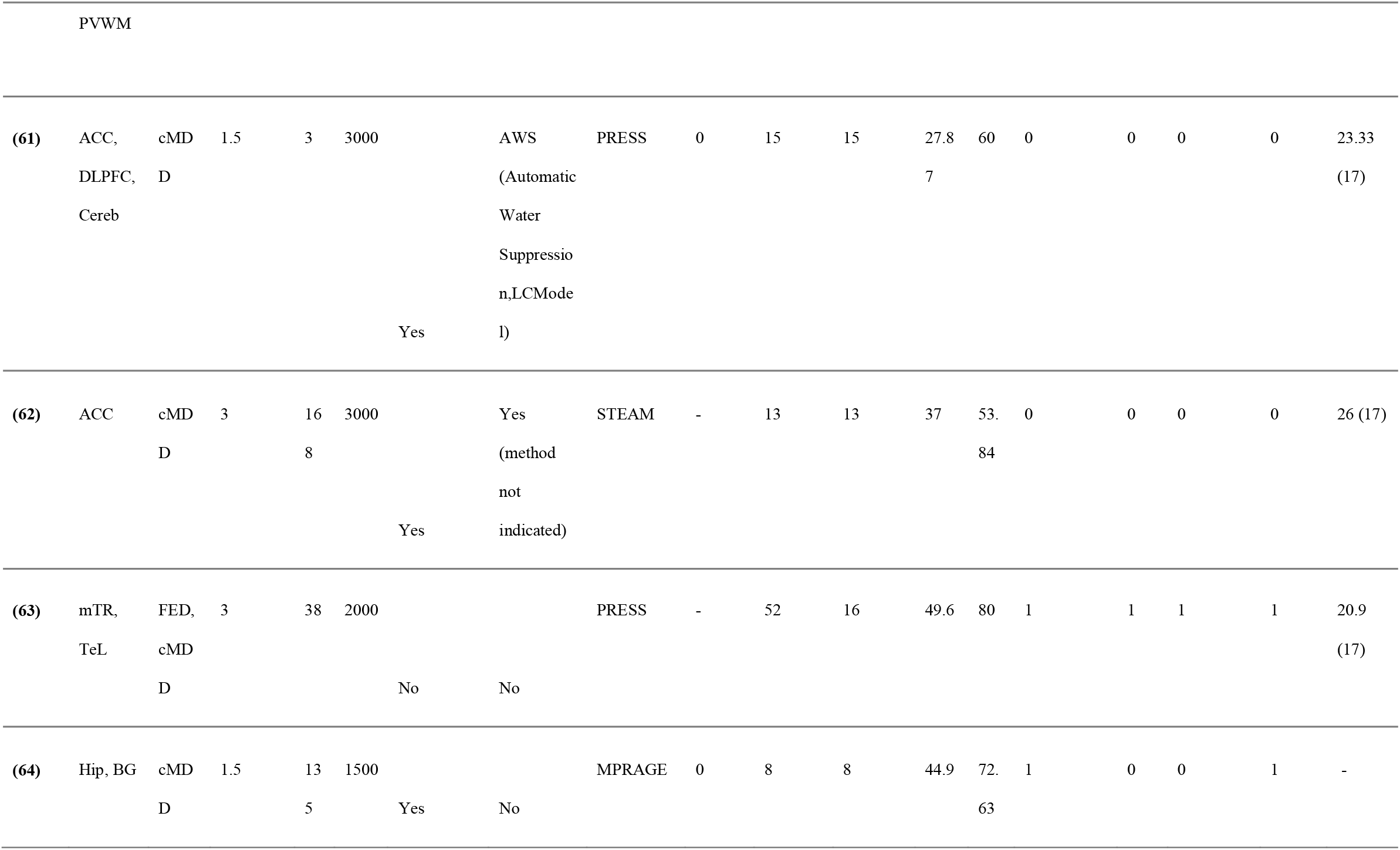
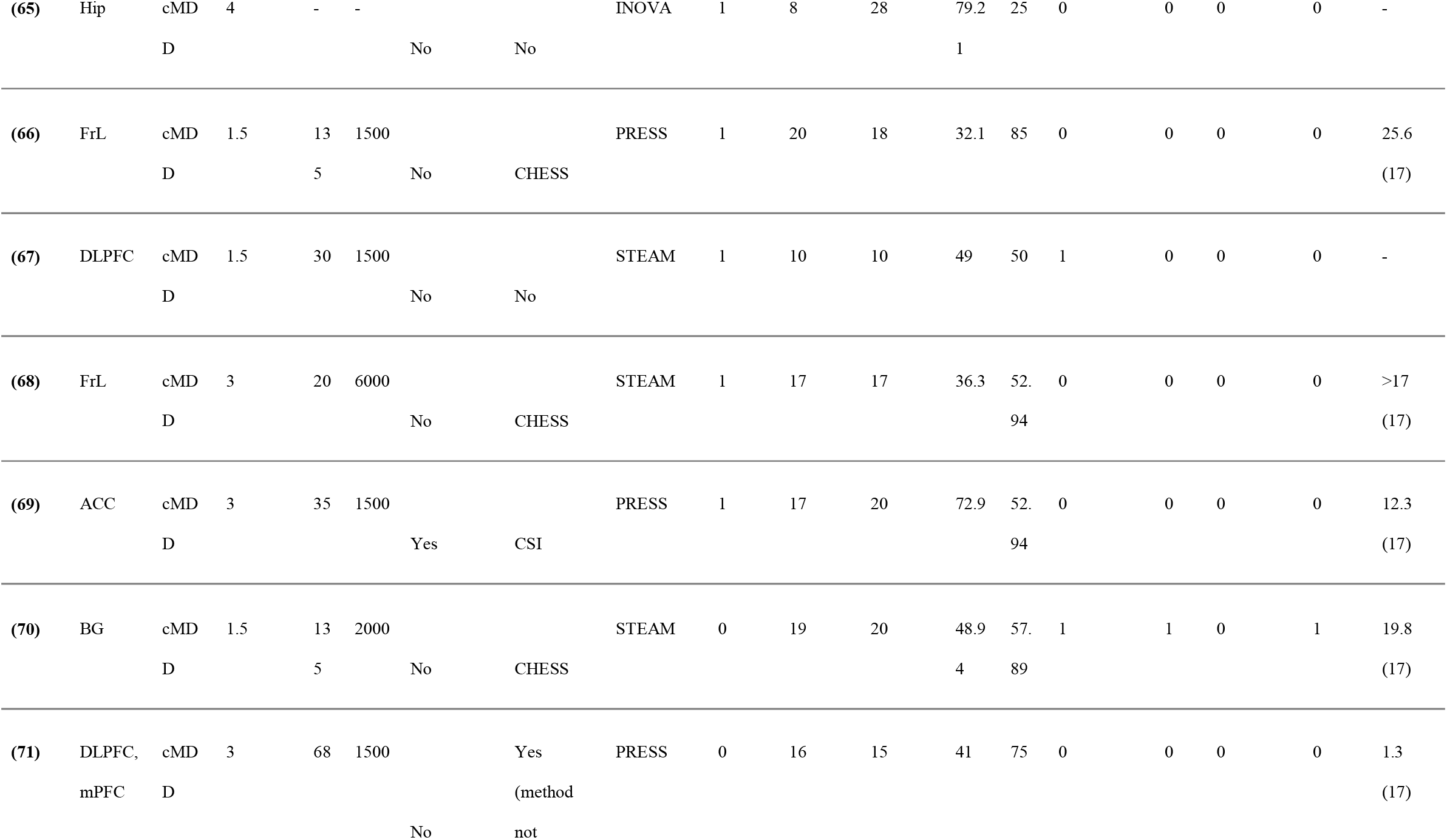
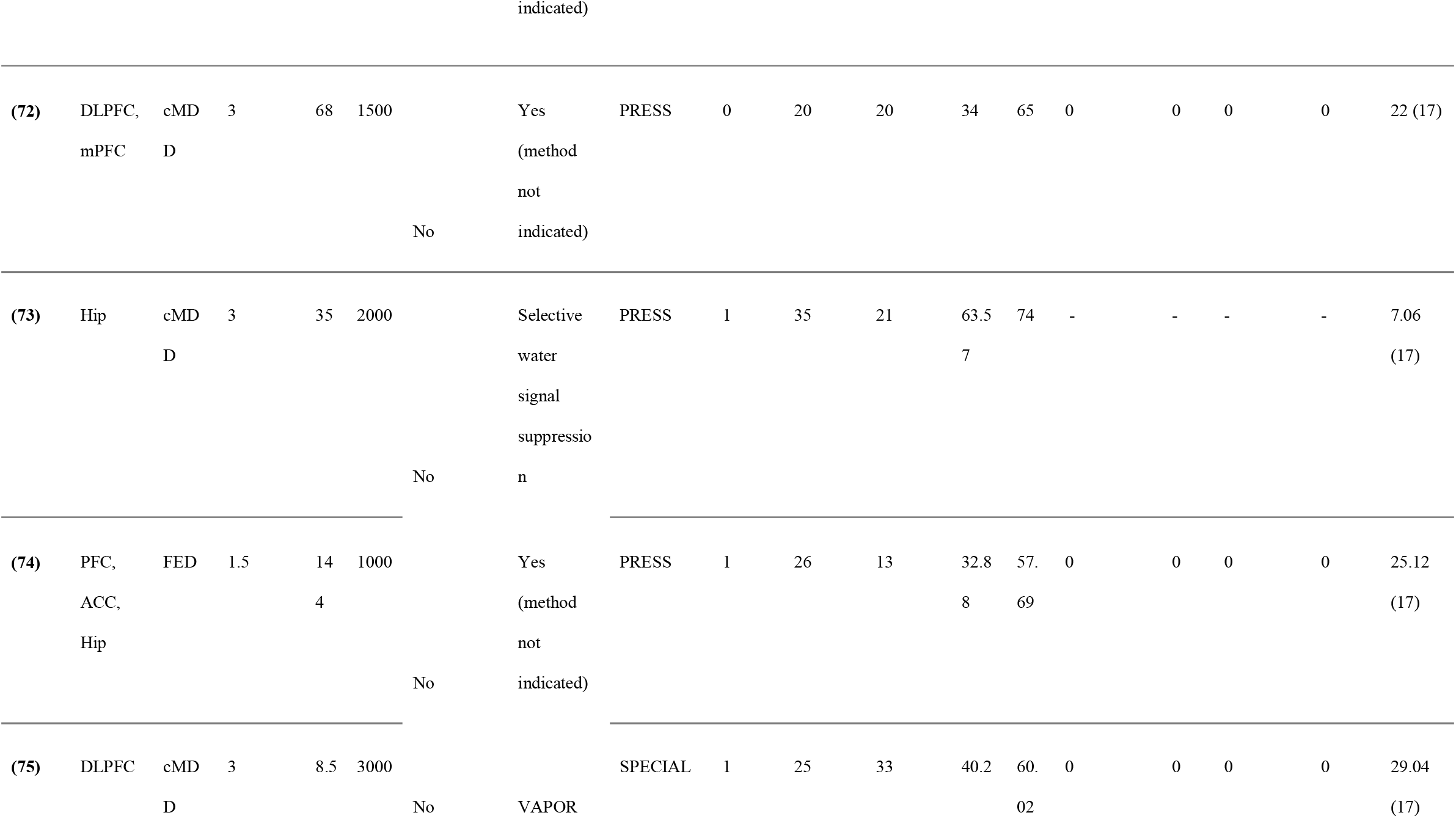
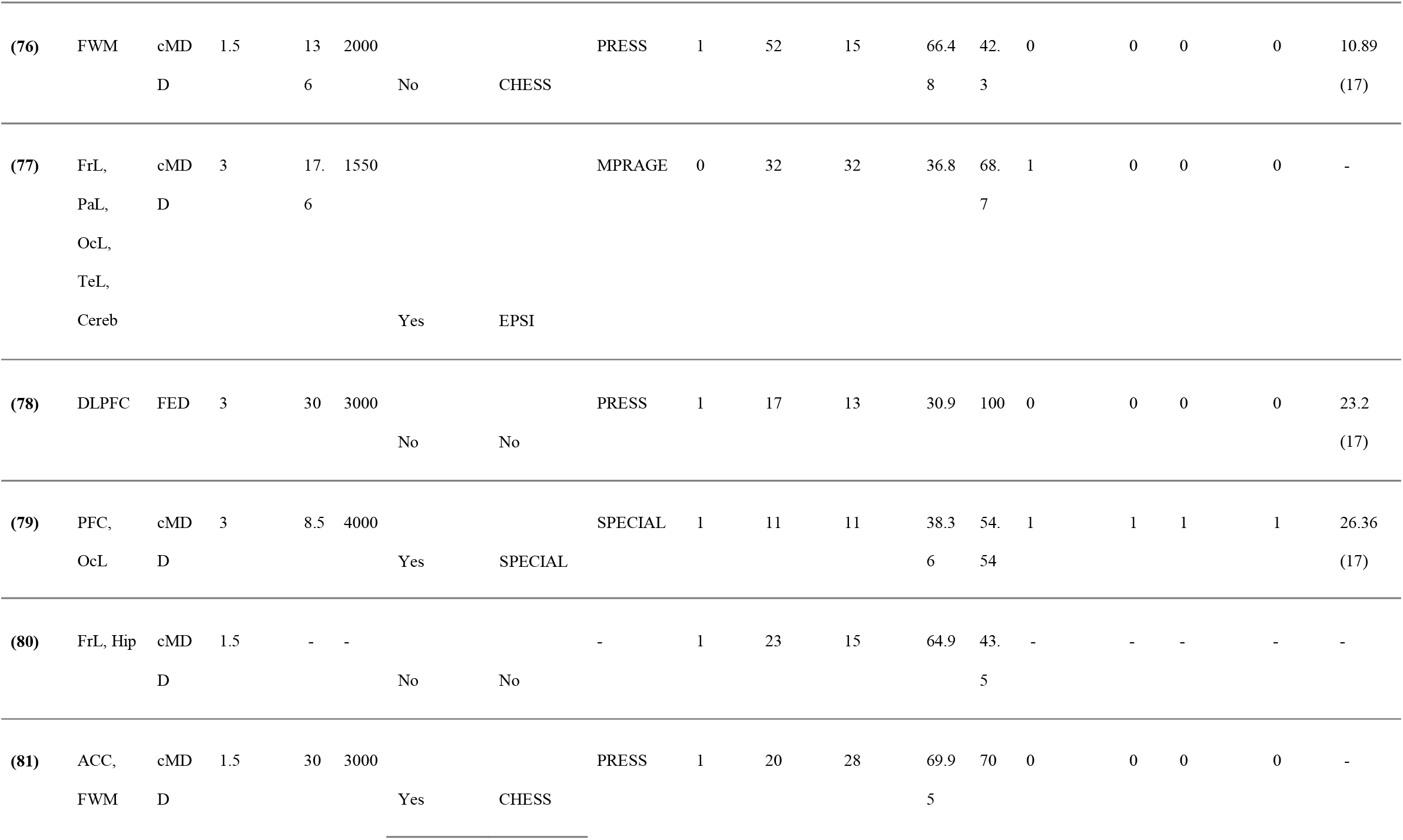
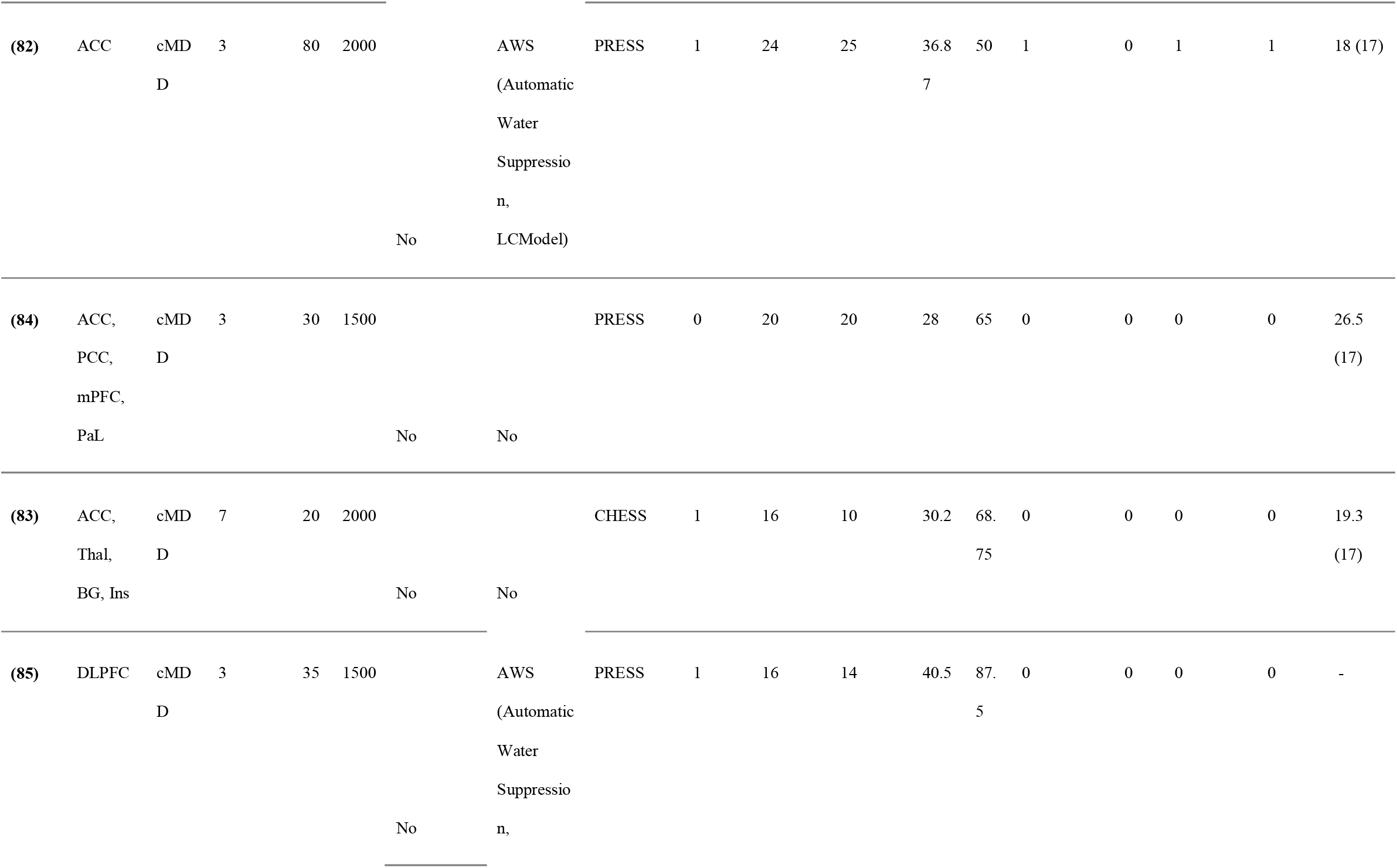
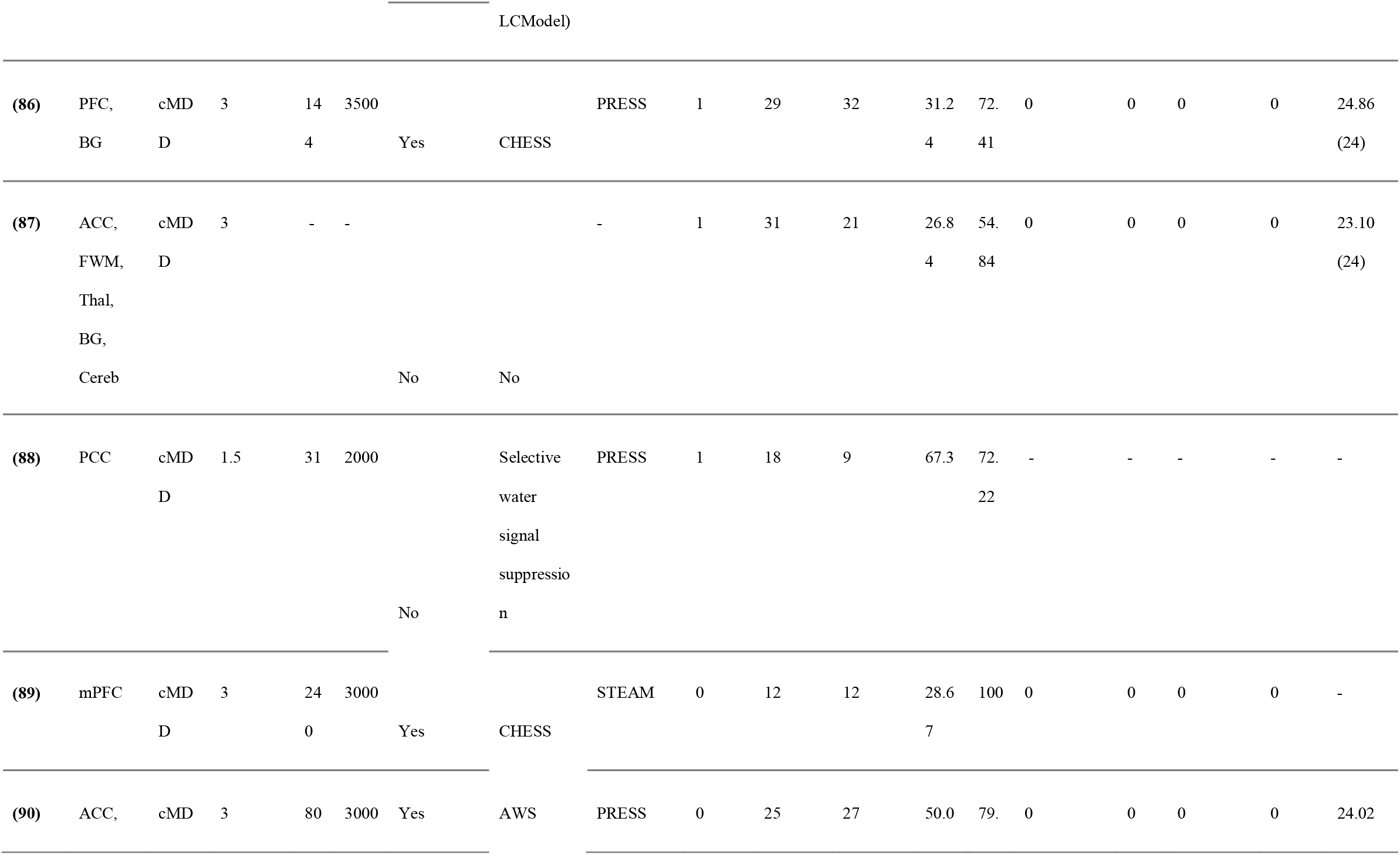
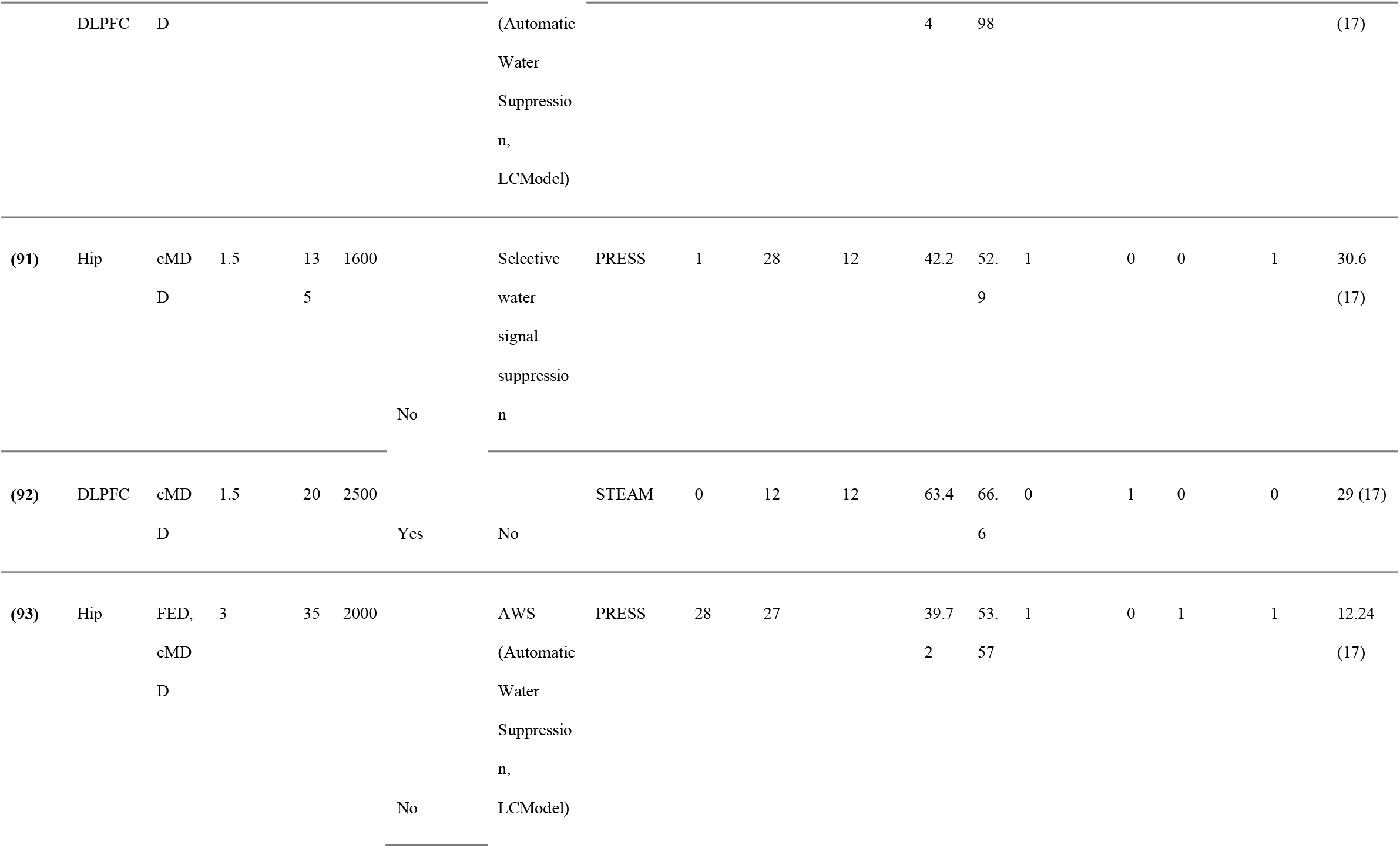
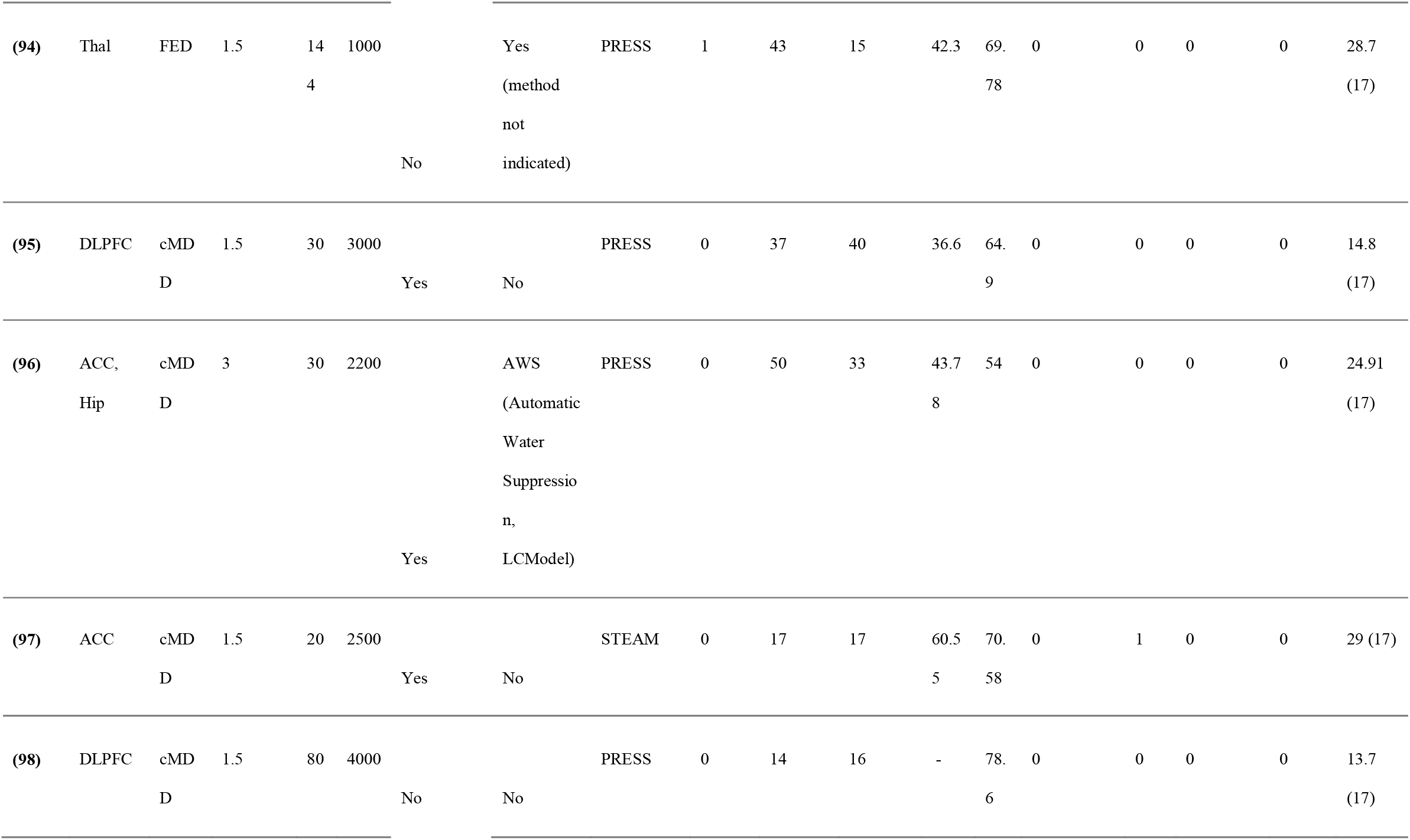
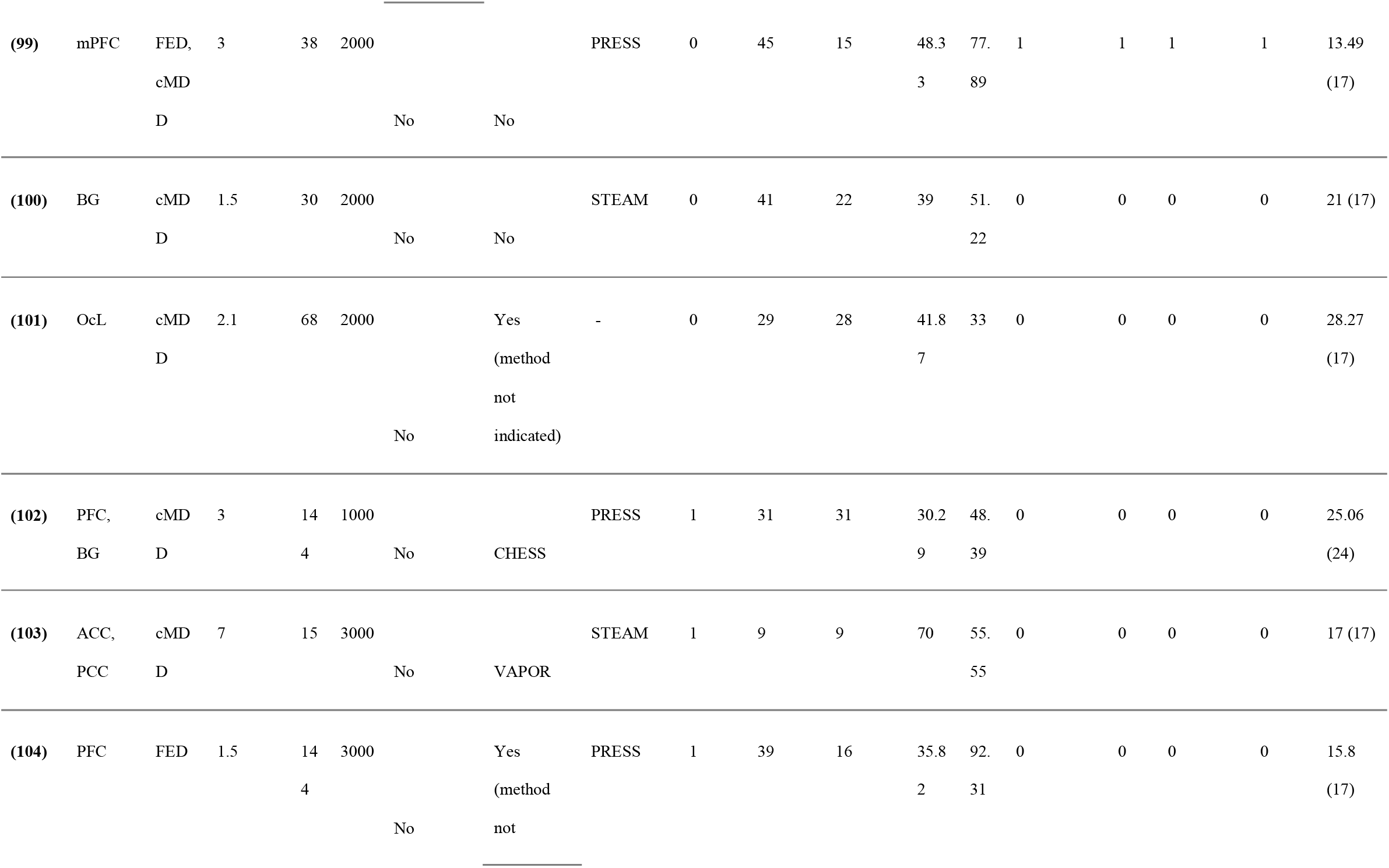
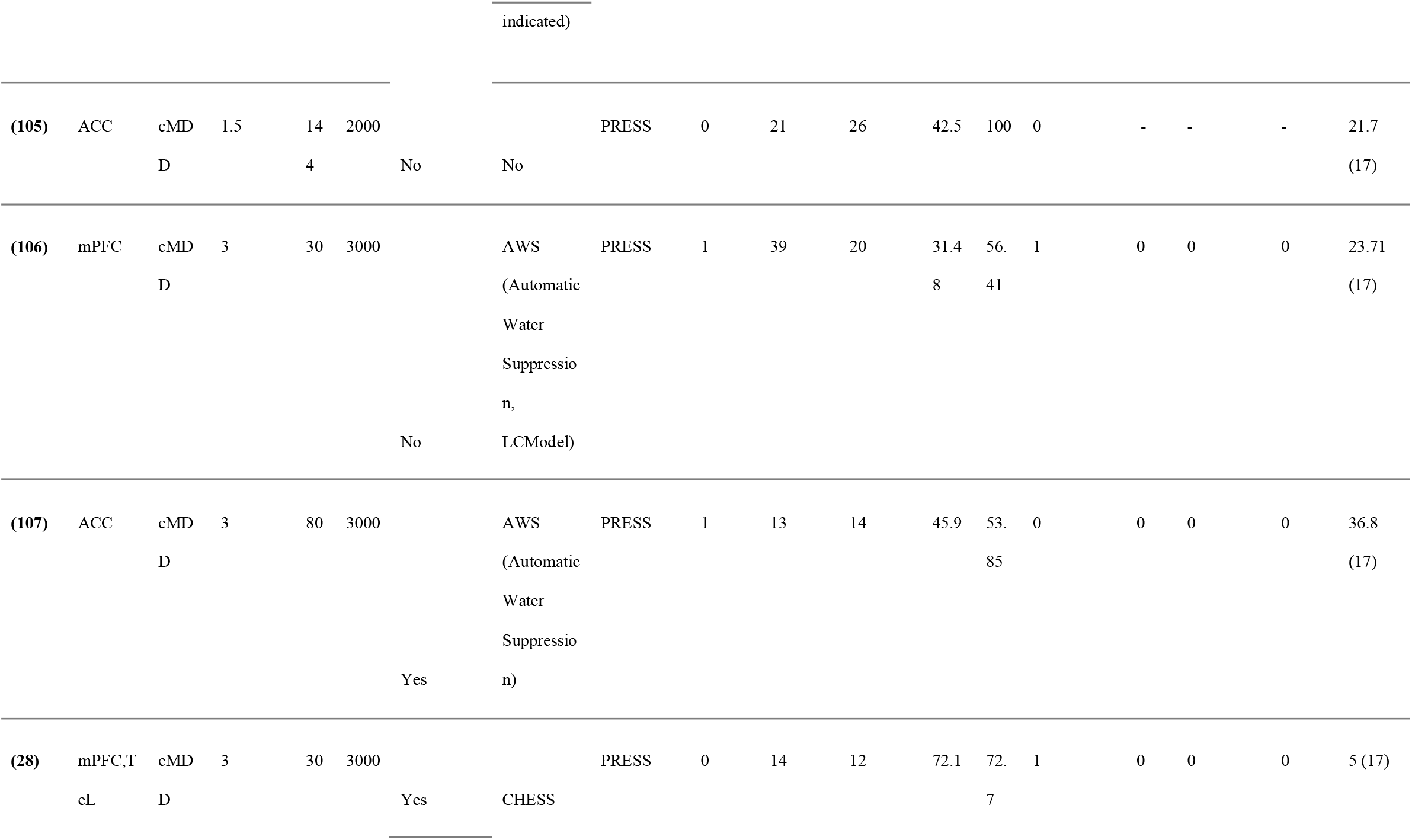
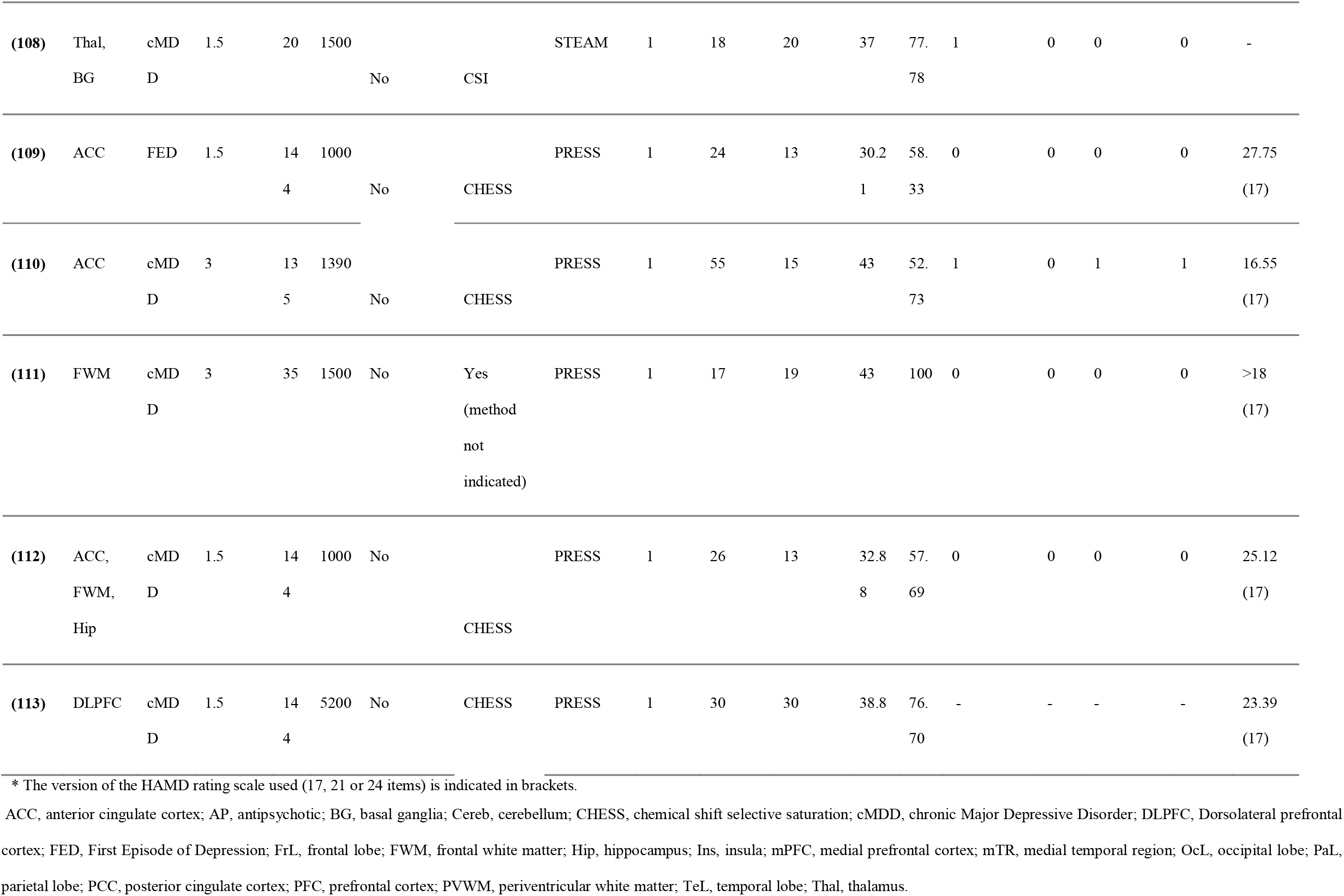
Characteristics of the studies included in the meta-analysis.

Three studies analyzed patients in total remission(28, 57, 71), 7 studies analyzed geriatric subjects(28, 59, 65, 69, 81, 88, 103), and 1 study patients with age > 50 years(73). One study analyzed post-partum depression(89). One study analyzed subjects with type 2 diabetes(53), 4 studies analyzed subjects with comorbid Alzheimer’s disease or mild cognitive impairment(69, 80, 88, 103), 1 study analyzed subjects with migraine(85) and 1 study subjects with chronic back pain(67).

In **Table 2** we report the results of the meta-analyses, while the main results are visually summarized in **Figure 2**. In the following paragraphs, we summarize the results in FED and in cMDD. The detailed results, including forest plots, influence analyses, sensitivity analyses, funnel plots and p-curve plots, are presented in **Supplementary Figures 1-51**.

**Table 2.**
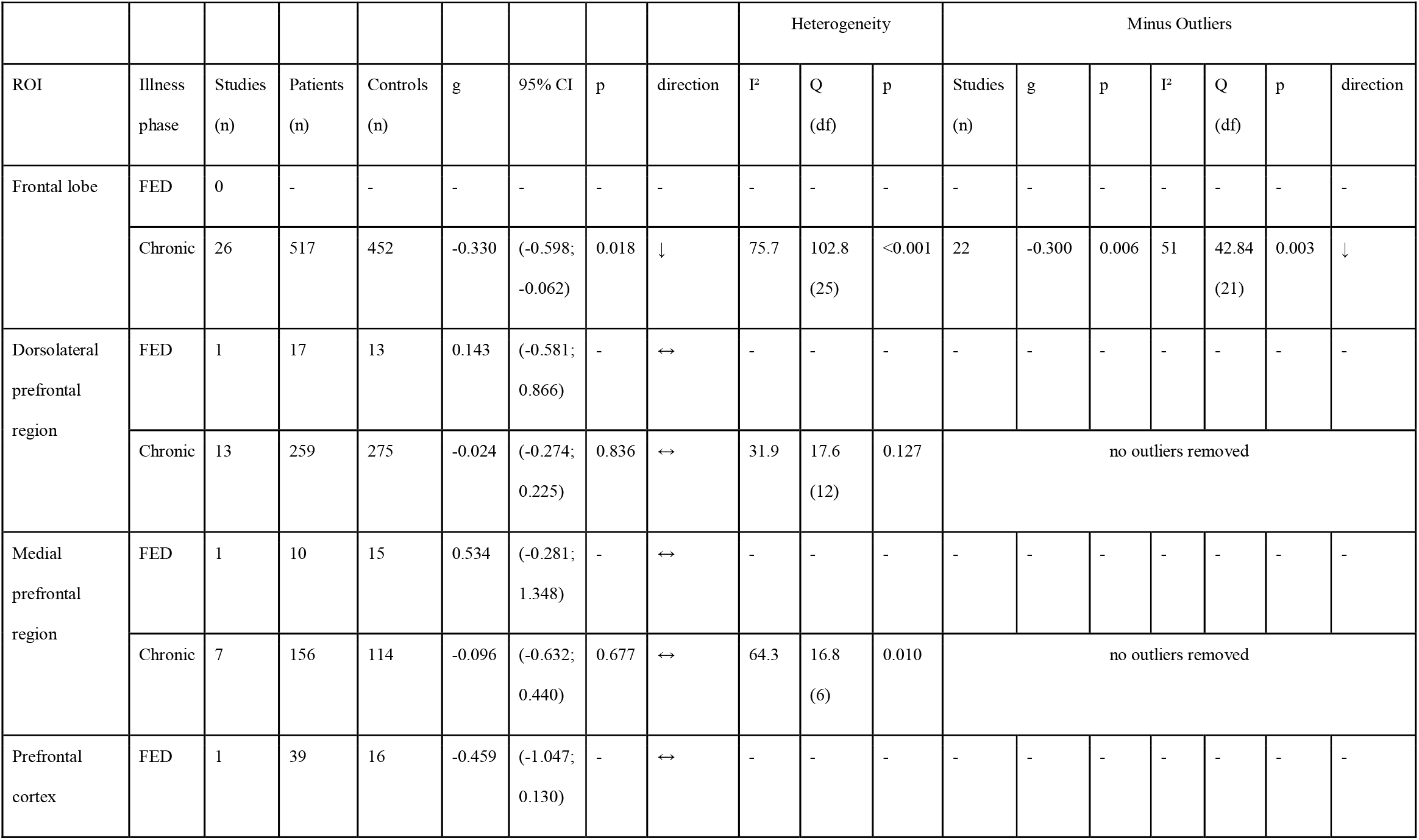
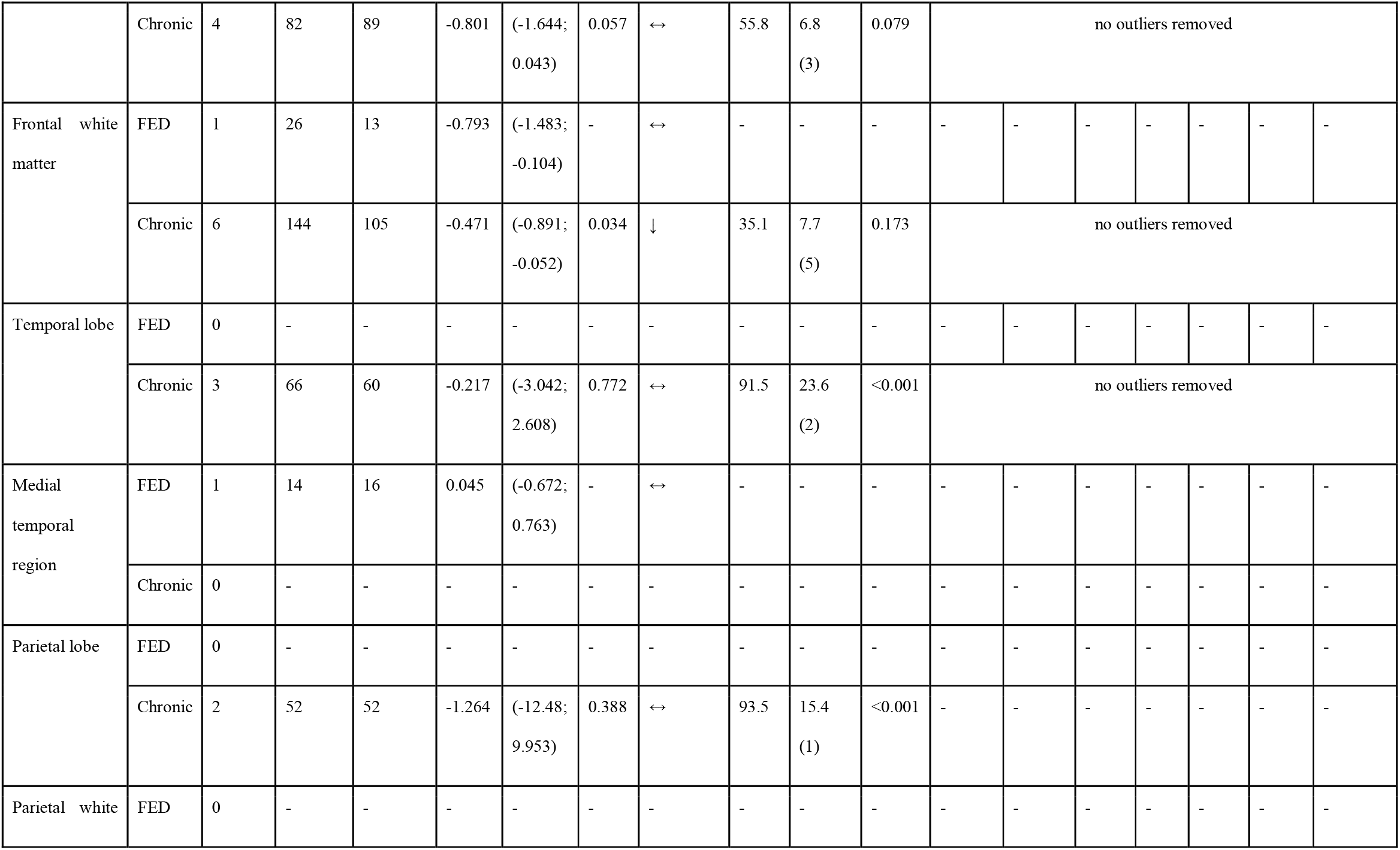
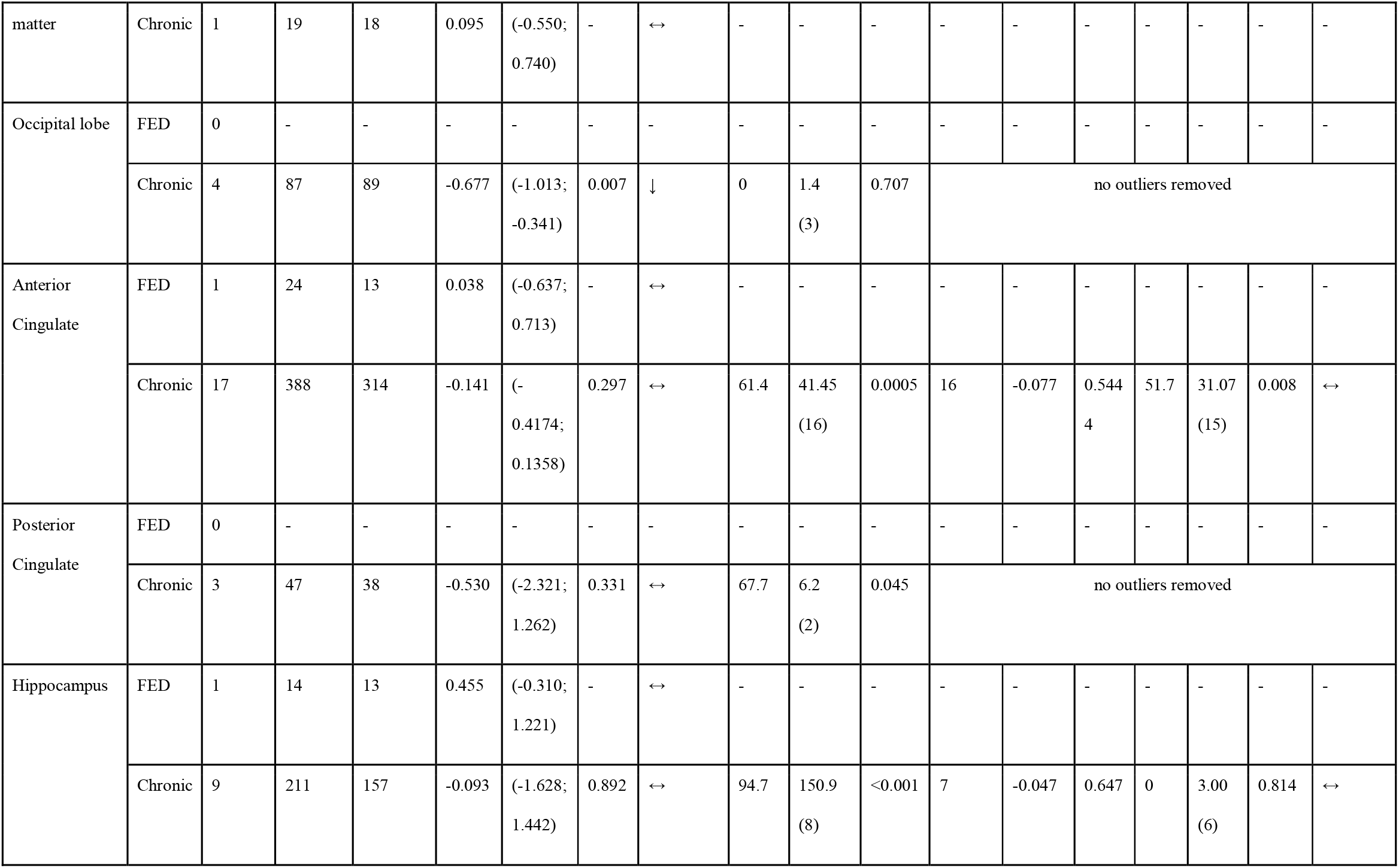
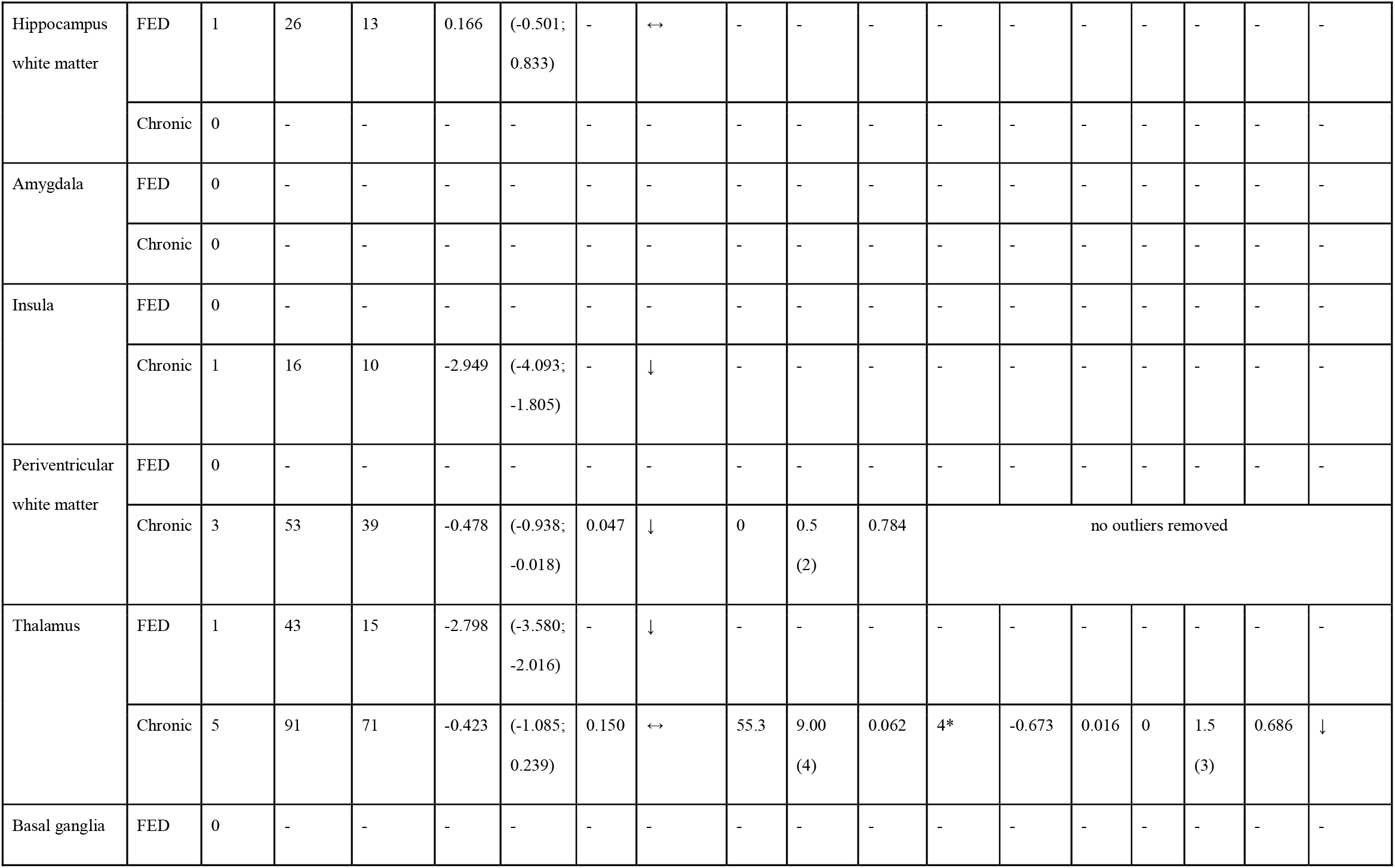
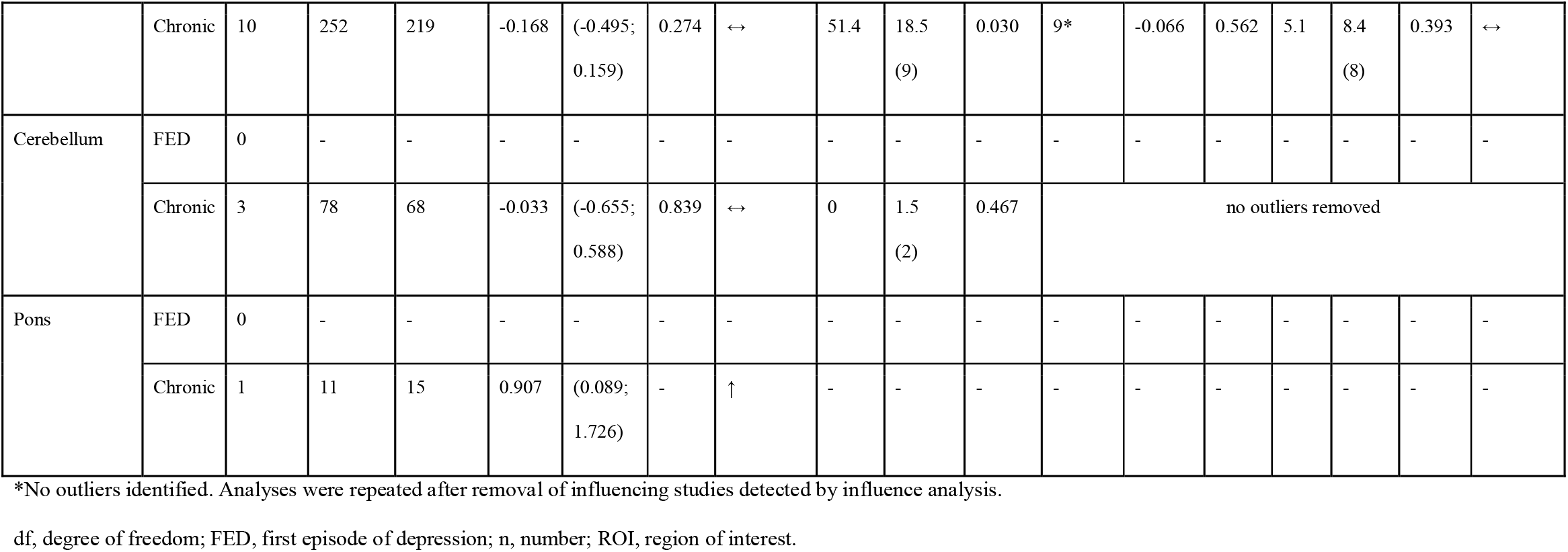
Summary of meta-analysis results.

**Figure 2.**
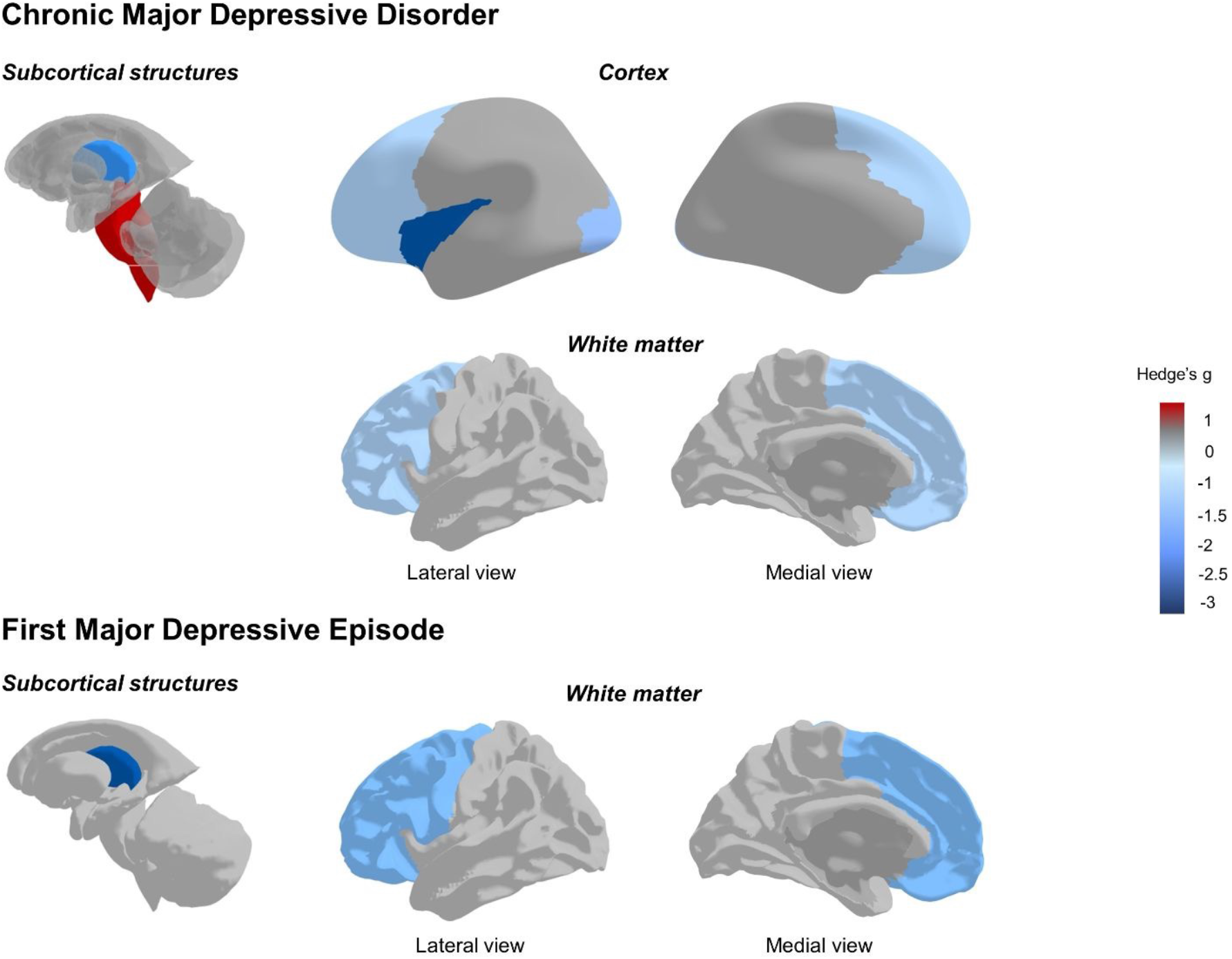
Visual summary of meta-analysis results. Areas of significant difference in NAA between patients with MDD and controls are colored, while non-significant or untested areas are shown in grey. Negative Hedges’ g (lower NAA levels in MDD than controls) are depicted in shades of blue; positive Hedges’ g (higher NAA levels in MDD than controls) are depicted in red.

### 3.2. First episode depression

We extracted information on NAA in 242 FED patients and 128 HC from 8 studies. We could not pool effect sizes in any brain region, since we retrieved a single study for each brain region (i.e. DLPFC and mPFC, PFC, ACC, FWM, medial temporal region, hippocampus, and thalamus). Primary studies showed significantly lower NAA concentrations in FEP than controls in the thalamus (n = 1, Hedges’ g = -2.789, 95% CI - 3.580 to -2.016) and FWM (n = 1, Hedges’ g = -0.793, 95% CI -1.483 to -0.104).

### 3.3. Chronic Major Depression Disorder

NAA levels were measured in 1308 patients with cMDD and 1114 HC from 57 studies. Relative to controls, cMDD patients had significantly lower cortical NAA levels, in the frontal lobe (n = 26, Hedges’ g = -0.330, 95% CI -0.598 to -0.062; p = 0.018; Q = 102.84, I^2^ = 75.7%, p < 0.001), and the occipital lobe (n = 4, Hedges’ g = -0.677, 95% CI -1.013 to -0.341; p = 0.007; Q = 1.39, I^2^ = 0%, p = 0.707). Sub-analyses in frontal lobe subregions - DLPFC (n = 13, Hedges’ g = -0.024, 95% CI -0.274 to 0.225; p = 0.836; Q = 17.63, I^2^ = 31.9%, p = 0.127), PFC (n = 4, Hedges’ g = -0.801, 95% CI -1.644 to -0.043; p = 0.057; Q = 6.79, I^2^ = 55.8%, p = 0.079), and mPFC (n = 7, Hedges’ g = -0.096, 95% CI -0.632 to 0.440; p = 0.677; Q = 16.81, I2 = 64.3%, p = 0.01)-did not show any difference between cMDD and controls. A single study compared NAA levels in the insula between cMDD and controls (n = 1, Hedges’ g = -2.949, 95% CI -4.093 to -1.805)(83). There were no significant differences between cMDD patients and HC in the parietal lobe, temporal lobe, and limbic lobe.

Lower NAA levels were also found within the white matter (WM), both in the FWM (n = 6, Hedges’ g = -0.471, 95% CI -0.891 to -0.052; p = 0.034; Q = 7.71, I^2^ = 35.1%, p = 0.173) and in the PVWM (n = 3, Hedges’ g = -0.478, 95% CI -0.938 to -0.018; p = 0.047; Q = 0.49, I^2^ = 0%, p = 0.784).

Finally, we observed some evidence of a difference in NAA levels between cMDD patients and controls in subcortical structures. In the thalamus, the meta-analysis of the 5 included studies could not detect any significant difference (n = 5, Hedges’ g = -0.423, 95% CI -1.085 to 0.234; p = 0.150; Q = 8.95, I^2^ = 55.3%, p = 0.062). The results became significant after removal of one influencing study (n = 4, Hedges’ g = -0.673, 95% CI -1.108 to -0.238; p = 0.016; Q = 1.49, I^2^ = 0%, p = 0.686)(108). A single primary study showed an increase in NAA levels in the pons of cMDD patients as compared to controls (n = 1, Hedges’ g = 0.907, 95% CI 0.089 to 1.726). No significant differences were found in the BG and cerebellum.

When sensitivity analysis based on the *leave-one-out* method found studies heavily distorting the meta-analytical estimates (i.e. in the occipital lobe and hippocampus), the pooled effect size remained significant after re-running the meta-analysis omitting the influential studies (**Table 2**).

#### 3.3.1. Sensitivity analysis for the effect of antidepressants on NAA levels in cMDD patients

Sensitivity analyses for the effect of antidepressants on meta-analytical estimates were conducted when at least 4 studies were available for each of the subgroups (i.e. patients treated or not with antidepressants), i.e., in the frontal lobe and BG. These studies focused on patients in remission, during a depressive episode, or on a mixed sample. Results are summarized in **Supplementary Table 13** and **Supplementary Figure 51**.

##### 3.3.1.1. Frontal lobe

The studies that did not specify whether patients were under antidepressant treatment or not (n = 2) were excluded from the analysis. Thus, 24 studies were finally included, of which 7 allowed antidepressant drugs, while 17 were on patients not taking antidepressant drugs. The difference in NAA levels between cMDD patients and HC remained significant only in the subgroup of studies not allowing antidepressant drugs (n = 17, Hedges’ g = 0.373, 95% CI 0.110 to 0.636, p _within subgroup_ = 0.005). Of note, a significant between-group heterogeneity could not be demonstrated (p _between subgroups_ = 0.900;), likely due to the overall large heterogeneity (Q = 102.84, *I*^*2*^ = 75.7%, p < 0.0001) (**Supplementary Table 13**).

##### 3.3.1.2. Basal ganglia

The studies that did not specify whether patients were under antidepressant treatment or not (n = 2) were excluded from the analysis. Thus, 8 studies were included, of which 4 allowed antidepressant drugs while 4 were on patients not taking antidepressant drugs. No significant difference emerged between subgroups (p_between subgroups_= 0.990; **Supplementary Table 13**).

#### 3.3.2. Other subgroup and sensitivity analyses, and meta-regressions in cMDD patients

The frontal lobe, ACC, hippocampus, and BG had enough studies to conduct subgroup analyses. None of the categorical clinical and methodological moderators significantly modified any analyses in these brain regions, as shown in **Supplementary Tables 4-1**4 As shown in **Supplementary Figure 4** and **10**, the only continuous moderator that significantly, albeit slightly, affected the meta-analytical estimates was publication year, with effect sizes growing more negative in more recent studies. Of note, although the subgroup analysis for field strength did not detect a significant difference between the subgroups, the meta-analytical estimate of the difference in NAA levels in the frontal lobe between cMDD and HC remained significant in the subgroup of studies conducted at field strength > 1.5 T only.

The sensitivity analysis on the influence of geriatric populations did not uncover significant changes in the results in each brain region after the exclusion of geriatric patients (**Supplementary Results**).

#### 3.3.3. Publication bias

The inspection of the funnel plots did not suggest the presence of publication bias for any of the brain regions analyzed but the FWM, where a certain asymmetry could be observed, with small studies having non-significant positive effect sizes missing (**Supplementary Figure 48**). However, there were not enough studies to conduct the Egger’s test or p-curve analysis. Egger’s tests were non-significant for all the analyses, indicating the absence of publication bias. The only p-curve plot not showing a rightward skew in p-values (i.e., the pattern associated with true effects) was the DLPFC (**Supplementary Figure 12)**.

## 4. Discussion

In this study, we compared NAA levels in broad brain regions between patients with MDD and HC, while also considering factors that can affect NAA levels, such as disease stage (FED versus cMDD) and severity, age, medication status, and ^1^H-MRS-related methodological factors. With medium effect sizes, NAA levels were lower within the frontal and occipital lobe, the thalamus, FWM, and PVWM in patients with cMDD as compared to HC. We highlighted a gap of knowledge regarding NAA levels in FED, which needs to be addressed to elucidate if NAA can be considered a marker of MDD neuroprogression. We observed that NAA levels were lower within the frontal lobe in unmedicated patients with cMDD as compared to HC, while no significant differences were found between medicated patients and HC. We found a relationship between the effect sizes of NAA levels in the frontal lobe and publication year, with effect sizes growing more negative (i.e. lower NAA levels in patients with cMDD than controls) in recent studies. This association may be explained by recent technological advances that allow higher field strengths. In fact, the meta-analytical estimate of the difference in NAA levels between cMDD and HC remained significant in the subgroup of studies employing field strength above 1.5 T only. None of the other clinical or ^1^H-MRS-related methodological moderators had a significant impact on the effect sizes.

Previous analyses of ^1^H-MRS-derived NAA levels in MDD found no convincing evidence of cerebral NAA alterations in MDD(22, 23). Contrary to what we show here, two previous reports could not demonstrate a reduction of NAA levels in the frontal lobe of patients with MDD as compared to HC, either because their analysis lacked power (n = 8 and n = 11 versus n = 26) or because more negative effect sizes emerged from more recent studies(77, 86). Our findings expand and confirm their negative findings within the BG(23).

NAA is broadly regarded as a marker of neuronal integrity and trophism. While historically considered a disorder of monoamine unbalance(114), the pathophysiology of MDD now appears to include deficiency in neuronal plasticity, and neuronal and astro-glial atrophy(115-117). More recently, NAA has been proposed as a marker of neuronal metabolic function(7). Using functional MRI and PET, hypoactivity/hypometabolism has been demonstrated in MDD patients within several brain regions(21, 118). Our findings of reduced NAA levels in the aforementioned brain regions thus converge with the existing evidence supporting impairment or abnormalities in the frontal and occipital(119-122) lobe, insula(123), thalamus(124-127), and WM(128, 129) in MDD. In particular, we found evidence for significantly reduced NAA levels in both the frontal lobe and the FWM in cMDD. The only study reporting on NAA levels in the FWM in FED patients showed a significant reduction. These findings are in line with solid evidence arguing for dysfunction of circuits involving the frontal lobe in MDD(119), which, critically, may underlie pivotal MDD symptoms such as anhedonia symptom(130) or rumination(26, 131). In fact, even though our results do not support a direct link between NAA levels and MDD symptoms, anhedonia has been linked with dysfunction in the reward circuits involving the frontal lobe(132-134). Similarly, a meta-analysis(135) showed that alterations in the dorsomedial prefrontal cortex subsystem of the Default Mode Network (DMN) may be a neural substrate of rumination, and a recent dynamic-functional connectivity transdiagnostic study on unipolar and bipolar depression highlighted that clinical depression levels modulated DMN duration, i.e. the time during which brain activity takes on a configuration corresponding to the DMN(136).

In our subgroup analysis of the effect of antidepressant treatment, we observed that NAA levels were lower within the frontal lobe in unmedicated patients with cMDD as compared to HC, while no significant differences were found between medicated patients and HC. This finding may suggest that antidepressant treatment could modify cerebral NAA levels. Several lines of preclinical and clinical evidence support that antidepressant administration may reverse neurobiological changes associated with MDD(116). However, the effect of antidepressant treatment on NAA levels has been investigated by a few original articles(61, 103, 111) with inconclusive results due to the small sample size.

### 4.1. Strengths and limitations

Our meta-analysis has several strengths. First, we separately analyzed FED and cMDD to discriminate if NAA should be considered a marker of either vulnerability or neurodegenerative processes in MDD. We highlighted a knowledge gap regarding NAA levels in FED. We could only retrieve one study per brain area, which did not allow to draw conclusions on whether cerebral NAA levels vary with MDD progression. Future longitudinal studies are needed to better elucidate whether NAA may represent an early biomarker of MDD neuroprogression. Understanding the pathophysiology underlying NAA alterations in MDD might help build an integrated clinicopathological staging model for MDD, with important clinical implications.

On the other hand, we had a reasonable sample size and statistical power to obtain significant findings in multiple brain regions in cMDD. As we were expecting high heterogeneity, we used a random-effects model which is highly robust to heterogeneity. We found significant moderate to high heterogeneity in all the global effect size estimates but the occipital lobe, the cerebellum, the FWM and the PVWM. After removal of outliers or influential studies, null or low heterogeneity remained in the hippocampus, thalamus and BG, while heterogeneity remained significant and moderate in the frontal lobe and ACC. Removal of outliers or influential studies had no major impact on our findings, except for the analysis of thalamic NAA levels in cMDD that went from non-significant to significant. We analyzed the impact of several categorical and continuous moderators on the effect sizes by means of subgroup analyses and meta-regressions. We defined an a-priori requirement for four datapoints per subgroup analysis and ten datapoints per meta-regression, so as to have enough statistical power to detect significant effects. We observed a significant relationship between effect size and publication year in the frontal lobe. We speculated that technical advances may have improved sensitivity. On this line, in our subgroup analysis of field strength, the difference between NAA levels in the frontal lobe of patients with cMDD vs HC remained significant in the subgroup of studies conducted at field strength above 1.5 T while it was no longer detectable in studies conducted at 1.5 T, although the two subgroups were not significantly different. The use of devices with higher magnetic field (3 T and higher) can enable smaller voxel size, which in turn may allow the acquisition of signal from smaller anatomical structures reducing partial volume effects due to inclusion of surrounding tissue and cerebral fluid in addiction to target structures. For in vivo spectroscopy, signal-to-noise ratio is expected to increase linearly with field strength(137). However, previous studies showed that such improvements may be offset by other factors, such as T2 relaxation time differences, line-broadening due to magnetic susceptibility effects, and radiofrequency coil efficiency(138). Unfortunately, our meta-analysis was not designed to assess the impact of the interaction of such a plethora of factors on spectra recorded at different field strengths. Future experiments should ascertain if using field strengths of 3 T or higher offer an advantage in terms of spectra resolution. None of the methodological moderators tested individually influenced our findings, therefore heterogeneity in the frontal lobe and ACC remained largely unexplained.

It has been hypothesized that lower NAA levels found in patients with psychotic disorders (schizophrenia, schizoaffective disorder, or bipolar disorder) may not reflect truly reduced NAA concentrations, but that they may rather be an effect of abnormal T2 relaxation times in these populations, in particular at longer TE due to differences in decay of resonance signal (139). There is limited evidence that MDD, too, may be associated with abnormal MRI T2 relaxation times in certain brain regions (99). Our meta-regressions evaluating TE as a continuous moderator in the frontal lobe, DLPFC, ACC, and BG do not support an impact of TE on the meta-analytical estimates.

## 4.2. Conclusions

The results of this meta-analysis suggest that cMDD is associated with lower NAA levels in the frontal and occipital lobes, thalamus, FWM, and PVWM. Due to the small number of studies reporting NAA levels in FED, we could not establish if NAA alterations are already present in the early stages of the disease. We found preliminary evidence that antidepressant treatment may reverse NAA alterations in the frontal lobe. Our findings support the hypometabolism hypothesis of depression. Although NAA is easily measured also at lower and more available MRI field strengths (7), it is possible that technical advances will be pivotal to establishing NAA as a biomarker of MDD and treatment response.

## Data Availability

All data produced in the present study are available upon reasonable request to the authors

https://www.crd.york.ac.uk/prospero/display_record.php?ID=CRD42020221050

## Acknowledgements

The authors would like to thank Noël Harris for her valuable help in proofreading the manuscript.

## Availability of Data and Materials

Data and materials will be provided upon request.

## Supplementary information

Supplementary information is available.

## REFERENCES

1. GBD2017Collaborators. Global, regional, and national age-sex-specific mortality for 282 causes of death in 195 countries and territories, 1980-2017: a systematic analysis for the Global Burden of Disease Study 2017. Lancet. 2018;392(10159):1736–88.

2. Ley P, Helbig-Lang S, Czilwik S, Lang T, Worlitz A, Brücher K, et al. Phenomenological differences between acute and chronic forms of major depression in inpatients. Nord J Psychiatry. 2011;65(5):330–7.

3. Liu J, Fan Y, Ling-Li Z, Liu B, Ju Y, Wang M, et al. The neuroprogressive nature of major depressive disorder: evidence from an intrinsic connectome analysis. Translational Psychiatry. 2021;11(1):102.

4. Dodd S, Berk M, Kelin K, Mancini M, Schacht A. Treatment response for acute depression is not associated with number of previous episodes: lack of evidence for a clinical staging model for major depressive disorder. J Affect Disord. 2013;150(2):344–9.

5. Unutzer J, Park M. Strategies to improve the management of depression in primary care. Prim Care. 2012;39(2):415–31.

6. Al-Harbi KS. Treatment-resistant depression: therapeutic trends, challenges, and future directions. Patient Prefer Adherence. 2012;6:369–88.

7. Whitehurst TS, Osugo M, Townsend L, Shatalina E, Vava R, Onwordi EC, et al. Proton Magnetic Resonance Spectroscopy of N-acetyl Aspartate in Chronic Schizophrenia, First Episode of Psychosis and High-Risk of Psychosis: A Systematic Review and Meta-Analysis. Neurosci Biobehav Rev. 2020;119:255–67.

8. Atagun MI, Sikoglu EM, Can SS, Ugurlu GK, Kaymak SU, Caykoylu A, et al. Neurochemical differences between bipolar disorder type I and II in superior temporal cortices: A proton magnetic resonance spectroscopy study. J Affect Disord. 2018;235:15–9.

9. Delvecchio G, Stanley JA, Altamura AC, Brambilla P. Metabolic alterations in generalised anxiety disorder: a review of proton magnetic resonance spectroscopic studies. Epidemiol Psychiatr Sci. 2017;26(6):587–95.

10. Jenkinson M, Chappell M. Introduction to neuroimaging analysis: Oxford University Press; 2018.

11. Maddock RJ, Buonocore MH. MR spectroscopic studies of the brain in psychiatric disorders. Curr Top Behav Neurosci. 2012;11:199–251.

12. Kang DW, Chu K, Yoon BW, Song IC, Chang KH, Roh JK. Diffusion-weighted imaging in Wallerian degeneration. J Neurol Sci. 2000;178(2):167–9.

13. Zhu X, Cao L, Hu X, Dong Y, Wang H, Liu F, et al. Brain metabolism assessed via proton magnetic resonance spectroscopy in patients with amnestic or vascular mild cognitive impairment. Clin Neurol Neurosurg. 2015;130:80–5.

14. Saindane AM, Cha S, Law M, Xue X, Knopp EA, Zagzag D. Proton MR spectroscopy of tumefactive demyelinating lesions. AJNR Am J Neuroradiol. 2002;23(8):1378–86.

15. Bivard A, Yassi N, Krishnamurthy V, Lin L, Levi C, Spratt NJ, et al. A comprehensive analysis of metabolic changes in the salvaged penumbra. Neuroradiology. 2016;58(4):409–15.

16. Demougeot C, Garnier P, Mossiat C, Bertrand N, Giroud M, Beley A, et al. N-Acetylaspartate, a marker of both cellular dysfunction and neuronal loss: its relevance to studies of acute brain injury. J Neurochem. 2001;77(2):408–15.

17. Veeramuthu V, Seow P, Narayanan V, Wong JHD, Tan LK, Hernowo AT, et al. Neurometabolites Alteration in the Acute Phase of Mild Traumatic Brain Injury (mTBI): An In Vivo Proton Magnetic Resonance Spectroscopy (1H-MRS) Study. Acad Radiol. 2018;25(9):1167–77.

18. Cvoro V, Marshall I, Armitage PA, Bastin ME, Carpenter T, Rivers CS, et al. MR diffusion and perfusion parameters: relationship to metabolites in acute ischaemic stroke. J Neurol Neurosurg Psychiatry. 2010;81(2):185–91.

19. Schuff N, Meyerhoff DJ, Mueller S, Chao L, Sacrey DT, Laxer K, et al. N-acetylaspartate as a marker of neuronal injury in neurodegenerative disease. Adv Exp Med Biol. 2006;576:241–62; discussion 361-3.

20. Clark J. N-acetyl aspartate: a marker for neuronal loss or mitochondrial dysfunction. Developmental neuroscience. 1998;20(4-5):271.

21. Su L, Cai Y, Xu Y, Dutt A, Shi S, Bramon E. Cerebral metabolism in major depressive disorder: a voxel-based meta-analysis of positron emission tomography studies. BMC Psychiatry. 2014;14(1):321.

22. Arnone D, Mumuni AN, Jauhar S, Condon B, Cavanagh J. Indirect evidence of selective glial involvement in glutamate-based mechanisms of mood regulation in depression: meta-analysis of absolute prefrontal neuro-metabolic concentrations. Eur Neuropsychopharmacol. 2015;25(8):1109–17.

23. Yildiz-Yesiloglu A, Ankerst DP. Review of 1H magnetic resonance spectroscopy findings in major depressive disorder: a meta-analysis. Psychiatry Res. 2006;147(1):1–25.

24. Moher D, Liberati A, Tetzlaff J, Altman DG, Group P. Preferred reporting items for systematic reviews and meta-analyses: the PRISMA statement. J Clin Epidemiol. 2009;62(10):1006–12.

25. Kumari V, Mitterschiffthaler MT, Teasdale JD, Malhi GS, Brown RG, Giampietro V, et al. Neural abnormalities during cognitive generation of affect in treatment-resistant depression. Biol Psychiatry. 2003;54(8):777–91.

26. Nolen-Hoeksema S, Wisco BE, Lyubomirsky S. Rethinking Rumination. Perspect Psychol Sci. 2008;3(5):400–24.

27. Moriguchi S, Takamiya A, Noda Y, Horita N, Wada M, Tsugawa S, et al. Glutamatergic neurometabolite levels in major depressive disorder: a systematic review and meta-analysis of proton magnetic resonance spectroscopy studies. Mol Psychiatry. 2019;24(7):952–64.

28. Venkatraman TN, Krishnan RR, Steffens DC, Song AW, Taylor WD. Biochemical abnormalities of the medial temporal lobe and medial prefrontal cortex in late-life depression. Psychiatry Res. 2009;172(1):49–54.

29. Stang A. Critical evaluation of the Newcastle-Ottawa scale for the assessment of the quality of nonrandomized studies in meta-analyses. Eur J Epidemiol. 2010;25(9):603–5.

30. Balduzzi S, Rucker G, Schwarzer G. How to perform a meta-analysis with R: a practical tutorial. Evid Based Ment Health. 2019;22(4):153–60.

31. Viechtbauer W, Cheung MW. Outlier and influence diagnostics for meta-analysis. Res Synth Methods. 2010;1(2):112–25.

32. Harrer M, Cuijpers P, Furukawa TA, Ebert DD. Doing Meta-Analysis with R: A Hands-On Guide. Boca Raton, FL and London: Chapmann & Hall/CRC [Internet]. 2021.

33. Harrer M, Cuijpers P, Furukawa TA, Ebert DD. Doing Meta-Analysis with R: A Hands-On Guide. Boca Raton, FL and London: Chapmann & Hall/CRC [Internet]. 2021.

34. Schwarzer G, Carpenter James R., Rücker, Gerta. meta-analysis with R 2015.

35. Higgins JP, Thompson SG. Quantifying heterogeneity in a meta-analysis. Stat Med. 2002;21(11):1539–58.

36. Olkin I, Dahabreh IJ, Trikalinos TA. GOSH - a graphical display of study heterogeneity. Res Synth Methods. 2012;3(3):214–23.

37. Egger M, Davey Smith G, Schneider M, Minder C. Bias in meta-analysis detected by a simple, graphical test. BMJ. 1997;315(7109):629–34.

38. Schwarzer G, Carpenter JR, Rücker G. meta-analysis with R 2015.

39. Capizzano AA, Jorge RE, Robinson RG. Limbic metabolic abnormalities in remote traumatic brain injury and correlation with psychiatric morbidity and social functioning. J Neuropsychiatry Clin Neurosci. 2010;22(4):370–7.

40. Urrila AS, Hakkarainen A, Castaneda A, Paunio T, Marttunen M, Lundbom N. Frontal Cortex Myo-Inositol Is Associated with Sleep and Depression in Adolescents: A Proton Magnetic Resonance Spectroscopy Study. Neuropsychobiology. 2017;75(1):21–31.

41. Chagas MHN, Tumas V, Pena-Pereira MA, Machado-de-Sousa JP, Carlos Dos Santos A, Sanches RF, et al. Neuroimaging of major depression in Parkinson’s disease: Cortical thickness, cortical and subcortical volume, and spectroscopy findings. J Psychiatr Res. 2017;90:40–5.

42. Zhang J, Narr KL, Woods RP, Phillips OR, Alger JR, Espinoza RT. Glutamate normalization with ECT treatment response in major depression. Mol Psychiatry. 2013;18(3):268–70.

43. Tollkotter M, Pfleiderer B, Soros P, Michael N. Effects of antidepressive therapy on auditory processing in severely depressed patients: a combined MRS and MEG study. J Psychiatr Res. 2006;40(4):293–306.

44. Hermens DF, Lagopoulos J, Naismith SL, Tobias-Webb J, Hickie IB. Distinct neurometabolic profiles are evident in the anterior cingulate of young people with major psychiatric disorders. Transl Psychiatry. 2012;2:e110.

45. Blasi G, Bertolino A, Brudaglio F, Sciota D, Altamura M, Antonucci N, et al. Hippocampal neurochemical pathology in patients at first episode of affective psychosis: a proton magnetic resonance spectroscopic imaging study. Psychiatry Res. 2004;131(2):95–105.

46. Xu J, Tang Y, Cecilio Baro C, Zhang X, Meng Z, Li Y. Left fimbria atrophy is associated with hippocampal metabolism in female major depressive disorder patients. Annu Int Conf IEEE Eng Med Biol Soc. 2018;2018:1136–9.

47. Wyckoff N, Kumar A, Gupta RC, Alger J, Hwang S, Thomas MA. Magnetization transfer imaging and magnetic resonance spectroscopy of normal-appearing white matter in late-life major depression. J Magn Reson Imaging. 2003;18(5):537–43.

48. Michael N, Erfurth A, Ohrmann P, Arolt V, Heindel W, Pfleiderer B. Neurotrophic effects of electroconvulsive therapy: a proton magnetic resonance study of the left amygdalar region in patients with treatment-resistant depression. Neuropsychopharmacology. 2003;28(4):720–5.

49. Jarnum H, Eskildsen SF, Steffensen EG, Lundbye-Christensen S, Simonsen CW, Thomsen IS, et al. Longitudinal MRI study of cortical thickness, perfusion, and metabolite levels in major depressive disorder. Acta Psychiatr Scand. 2011;124(6):435–46.

50. Yoon S, Kim JE, Hwang J, Kim TS, Kang HJ, Namgung E, et al. Effects of Creatine Monohydrate Augmentation on Brain Metabolic and Network Outcome Measures in Women With Major Depressive Disorder. Biol Psychiatry. 2016;80(6):439–47.

51. Block W, Traber F, von Widdern O, Metten M, Schild H, Maier W, et al. Proton MR spectroscopy of the hippocampus at 3 T in patients with unipolar major depressive disorder: correlates and predictors of treatment response. Int J Neuropsychopharmacol. 2009;12(3):415–22.

52. Rosa CE, Soares JC, Figueiredo FP, Cavalli RC, Barbieri MA, Schaufelberger MS, et al. Glutamatergic and neural dysfunction in postpartum depression using magnetic resonance spectroscopy. Psychiatry Res Neuroimaging. 2017;265:18–25.

53. Ajilore O, Haroon E, Kumaran S, Darwin C, Binesh N, Mintz J, et al. Measurement of brain metabolites in patients with type 2 diabetes and major depression using proton magnetic resonance spectroscopy. Neuropsychopharmacology. 2007;32(6):1224–31.

54. Auer DP, Putz B, Kraft E, Lipinski B, Schill J, Holsboer F. Reduced glutamate in the anterior cingulate cortex in depression: an in vivo proton magnetic resonance spectroscopy study. Biol Psychiatry. 2000;47(4):305–13.

55. Bernier D, Bartha R, Devarajan S, Macmaster FP, Schmidt MH, Rusak B. Effects of overnight sleep restriction on brain chemistry and mood in women with unipolar depression and healthy controls. J Psychiatry Neurosci. 2009;34(5):352–60.

56. Brambilla P, Stanley JA, Nicoletti MA, Sassi RB, Mallinger AG, Frank E, et al. 1H Magnetic resonance spectroscopy study of dorsolateral prefrontal cortex in unipolar mood disorder patients. Psychiatry Res. 2005;138(2):131–9.

57. Bhagwagar Z, Wylezinska M, Jezzard P, Evans J, Ashworth F, Sule A, et al. Reduction in occipital cortex gamma-aminobutyric acid concentrations in medication-free recovered unipolar depressed and bipolar subjects. Biol Psychiatry. 2007;61(6):806–12.

58. Charles HC, Lazeyras F, Krishnan KR, Boyko OB, Payne M, Moore D. Brain choline in depression: in vivo detection of potential pharmacodynamic effects of antidepressant therapy using hydrogen localized spectroscopy. Prog Neuropsychopharmacol Biol Psychiatry. 1994;18(7):1121–7.

59. Chen CS, Chiang IC, Li CW, Lin WC, Lu CY, Hsieh TJ, et al. Proton magnetic resonance spectroscopy of late-life major depressive disorder. Psychiatry Res. 2009;172(3):210–4.

60. Chen S, Lai L, Kang Z, Luo X, Zhang J, Li J. Imaging changes in neural circuits in patients with depression using (1)H-magnetic resonance spectroscopy and diffusion tensor imaging. Neural Regen Res. 2012;7(24):1881–8.

61. Chen LP, Dai HY, Dai ZZ, Xu CT, Wu RH. Anterior cingulate cortex and cerebellar hemisphere neurometabolite changes in depression treatment: A 1H magnetic resonance spectroscopy study. Psychiatry Clin Neurosci. 2014;68(5):357–64.

62. Coupland NJ, Ogilvie CJ, Hegadoren KM, Seres P, Hanstock CC, Allen PS. Decreased prefrontal Myo-inositol in major depressive disorder. Biol Psychiatry. 2005;57(12):1526–34.

63. de Diego-Adelino J, Portella MJ, Gomez-Anson B, Lopez-Moruelo O, Serra-Blasco M, Vives Y, et al. Hippocampal abnormalities of glutamate/glutamine, N-acetylaspartate and choline in patients with depression are related to past illness burden. J Psychiatry Neurosci. 2013;38(2):107–16.

64. Ende G, Demirakca T, Walter S, Wokrina T, Sartorius A, Wildgruber D, et al. Subcortical and medial temporal MR-detectable metabolite abnormalities in unipolar major depression. Eur Arch Psychiatry Clin Neurosci. 2007;257(1):36–9.

65. Ezzati A, Zimmerman ME, Katz MJ, Lipton RB. Hippocampal correlates of depression in healthy elderly adults. Hippocampus. 2013;23(12):1137–42.

66. Gonul AS, Kitis O, Ozan E, Akdeniz F, Eker C, Eker OD, et al. The effect of antidepressant treatment on N-acetyl aspartate levels of medial frontal cortex in drug-free depressed patients. Prog Neuropsychopharmacol Biol Psychiatry. 2006;30(1):120–5.

67. Grachev ID, Ramachandran TS, Thomas PS, Szeverenyi NM, Fredrickson BE. Association between dorsolateral prefrontal N-acetyl aspartate and depression in chronic back pain: an in vivo proton magnetic resonance spectroscopy study. J Neural Transm (Vienna). 2003;110(3):287–312.

68. Gruber S, Frey R, Mlynarik V, Stadlbauer A, Heiden A, Kasper S, et al. Quantification of metabolic differences in the frontal brain of depressive patients and controls obtained by 1H-MRS at 3 Tesla. Invest Radiol. 2003;38(7):403–8.

69. Guo Z, Zhang J, Liu X, Hou H, Cao Y, Wei F, et al. Neurometabolic characteristics in the anterior cingulate gyrus of Alzheimer’s disease patients with depression: a (1)H magnetic resonance spectroscopy study. BMC Psychiatry. 2015;15:306.

70. Hamakawa H, Kato T, Murashita J, Kato N. Quantitative proton magnetic resonance spectroscopy of the basal ganglia in patients with affective disorders. Eur Arch Psychiatry Clin Neurosci. 1998;248(1):53–8.

71. Hasler G, Neumeister A, van der Veen JW, Tumonis T, Bain EE, Shen J, et al. Normal prefrontal gamma-aminobutyric acid levels in remitted depressed subjects determined by proton magnetic resonance spectroscopy. Biol Psychiatry. 2005;58(12):969–73.

72. Hasler G, van der Veen JW, Tumonis T, Meyers N, Shen J, Drevets WC. Reduced prefrontal glutamate/glutamine and gamma-aminobutyric acid levels in major depression determined using proton magnetic resonance spectroscopy. Arch Gen Psychiatry. 2007;64(2):193–200.

73. Jayaweera HK, Lagopoulos J, Duffy SL, Lewis SJ, Hermens DF, Norrie L, et al. Spectroscopic markers of memory impairment, symptom severity and age of onset in older people with lifetime depression: Discrete roles of N-acetyl aspartate and glutamate. J Affect Disord. 2015;183:31–8.

74. Jia Y, Zhong S, Wang Y, Liu T, Liao X, Huang L. The correlation between biochemical abnormalities in frontal white matter, hippocampus and serum thyroid hormone levels in first-episode patients with major depressive disorder. J Affect Disord. 2015;180:162–9.

75. Jollant F, Near J, Turecki G, Richard-Devantoy S. Spectroscopy markers of suicidal risk and mental pain in depressed patients. Prog Neuropsychopharmacol Biol Psychiatry. 2016.

76. Kado H, Kimura H, Murata T, Nagata K, Kanno I. Depressive psychosis: clinical usefulness of MR spectroscopy data in predicting prognosis. Radiology. 2006;238(1):248–55.

77. Kahl KG, Atalay S, Maudsley AA, Sheriff S, Cummings A, Frieling H, et al. Altered neurometabolism in major depressive disorder: A whole brain (1)H-magnetic resonance spectroscopic imaging study at 3T. Prog Neuropsychopharmacol Biol Psychiatry. 2020;101:109916.

78. Kaymak SU, Demir B, Oguz KK, Senturk S, Ulug B. Antidepressant effect detected on proton magnetic resonance spectroscopy in drug-naive female patients with first-episode major depression. Psychiatry Clin Neurosci. 2009;63(3):350–6.

79. Knudsen MK, Near J, Blicher AB, Videbech P, Blicher JU. Magnetic resonance (MR) spectroscopic measurement of gamma-aminobutyric acid (GABA) in major depression before and after electroconvulsive therapy. Acta Neuropsychiatr. 2019;31(1):17–26.

80. Kotb MA, Kamal AM, Aldossary NM. Value of magnetic resonance spectroscopy in geriatric patients with cognitive impairment. Egypt J Neurol Psychiatry Neurosurg. 2020;56, 10.

81. Kumar A, Thomas A, Lavretsky H, Yue K, Huda A, Curran J, et al. Frontal white matter biochemical abnormalities in late-life major depression detected with proton magnetic resonance spectroscopy. Am J Psychiatry. 2002;159(4):630–6.

82. Li M, Metzger CD, Li W, Safron A, van Tol MJ, Lord A, et al. Dissociation of glutamate and cortical thickness is restricted to regions subserving trait but not state markers in major depressive disorder. J Affect Disord. 2014;169:91–100.

83. Li Y, Jakary A, Gillung E, Eisendrath S, Nelson SJ, Mukherjee P, et al. Evaluating metabolites in patients with major depressive disorder who received mindfulness-based cognitive therapy and healthy controls using short echo MRSI at 7 Tesla. MAGMA. 2016;29(3):523–33.

84. Li H, Xu H, Zhang Y, Guan J, Zhang J, Xu C, et al. Differential neurometabolite alterations in brains of medication-free individuals with bipolar disorder and those with unipolar depression: a two-dimensional proton magnetic resonance spectroscopy study. Bipolar Disord. 2016;18(7):583–90.

85. Lirng JF, Chen HC, Fuh JL, Tsai CF, Liang JF, Wang SJ. Increased myo-inositol level in dorsolateral prefrontal cortex in migraine patients with major depression. Cephalalgia. 2015;35(8):702–9.

86. Liu X, Zhong S, Li Z, Chen J, Wang Y, Lai S, et al. Serum copper and zinc levels correlate with biochemical metabolite ratios in the prefrontal cortex and lentiform nucleus of patients with major depressive disorder. Prog Neuropsychopharmacol Biol Psychiatry. 2020;99:109828.

87. Liu X, Zhong S, Yan L, Zhao H, Wang Y, Hu Y, et al. Correlations Among mRNA Expression Levels of ATP7A, Serum Ceruloplasmin Levels, and Neuronal Metabolism in Unmedicated Major Depressive Disorder. Int J Neuropsychopharmacol. 2020;23(10):642–52.

88. Martinez-Bisbal MC, Arana E, Marti-Bonmati L, Molla E, Celda B. Cognitive impairment: classification by 1H magnetic resonance spectroscopy. Eur J Neurol. 2004;11(3):187–93.

89. McEwen AM, Burgess DT, Hanstock CC, Seres P, Khalili P, Newman SC, et al. Increased glutamate levels in the medial prefrontal cortex in patients with postpartum depression. Neuropsychopharmacology. 2012;37(11):2428–35.

90. Merkl A, Schubert F, Quante A, Luborzewski A, Brakemeier EL, Grimm S, et al. Abnormal cingulate and prefrontal cortical neurochemistry in major depression after electroconvulsive therapy. Biol Psychiatry. 2011;69(8):772–9.

91. Mervaala E, Fohr J, Kononen M, Valkonen-Korhonen M, Vainio P, Partanen K, et al. Quantitative MRI of the hippocampus and amygdala in severe depression. Psychol Med. 2000;30(1):117–25.

92. Michael N, Erfurth A, Ohrmann P, Arolt V, Heindel W, Pfleiderer B. Metabolic changes within the left dorsolateral prefrontal cortex occurring with electroconvulsive therapy in patients with treatment resistant unipolar depression. Psychol Med. 2003;33(7):1277–84.

93. Milne A, MacQueen GM, Yucel K, Soreni N, Hall GB. Hippocampal metabolic abnormalities at first onset and with recurrent episodes of a major depressive disorder: a proton magnetic resonance spectroscopy study. Neuroimage. 2009;47(1):36–41.

94. Mohamed MA, Smith MA, Schlund MW, Nestadt G, Barker PB, Hoehn-Saric R. Proton magnetic resonance spectroscopy in obsessive-compulsive disorder: a pilot investigation comparing treatment responders and non-responders. Psychiatry Res. 2007;156(2):175–9.

95. Nery FG, Stanley JA, Chen HH, Hatch JP, Nicoletti MA, Monkul ES, et al. Normal metabolite levels in the left dorsolateral prefrontal cortex of unmedicated major depressive disorder patients: a single voxel (1)H spectroscopy study. Psychiatry Res. 2009;174(3):177–83.

96. Njau S, Joshi SH, Espinoza R, Leaver AM, Vasavada M, Marquina A, et al. Neurochemical correlates of rapid treatment response to electroconvulsive therapy in patients with major depression. J Psychiatry Neurosci. 2017;42(1):6–16.

97. Pfleiderer B, Michael N, Erfurth A, Ohrmann P, Hohmann U, Wolgast M, et al. Effective electroconvulsive therapy reverses glutamate/glutamine deficit in the left anterior cingulum of unipolar depressed patients. Psychiatry Res. 2003;122(3):185–92.

98. Pigoni A, Delvecchio G, Squarcina L, Bonivento C, Girardi P, Finos L, et al. Sex differences in brain metabolites in anxiety and mood disorders. Psychiatry Res Neuroimaging. 2020;305:111196.

99. Portella MJ, de Diego-Adelino J, Gomez-Anson B, Morgan-Ferrando R, Vives Y, Puigdemont D, et al. Ventromedial prefrontal spectroscopic abnormalities over the course of depression: a comparison among first episode, remitted recurrent and chronic patients. J Psychiatr Res. 2011;45(4):427–34.

100. Renshaw PF, Lafer B, Babb SM, Fava M, Stoll AL, Christensen JD, et al. Basal ganglia choline levels in depression and response to fluoxetine treatment: an in vivo proton magnetic resonance spectroscopy study. Biol Psychiatry. 1997;41(8):837–43.

101. Sanacora G, Gueorguieva R, Epperson CN, Wu YT, Appel M, Rothman DL, et al. Subtype-specific alterations of gamma-aminobutyric acid and glutamate in patients with major depression. Arch Gen Psychiatry. 2004;61(7):705–13.

102. Shan Y, Jia Y, Zhong S, Li X, Zhao H, Chen J, et al. Correlations between working memory impairment and neurometabolites of prefrontal cortex and lenticular nucleus in patients with major depressive disorder. J Affect Disord. 2018;227:236–42.

103. Smith GS, Oeltzschner G, Gould NF, Leoutsakos JS, Nassery N, Joo JH, et al. Neurotransmitters and Neurometabolites in Late-Life Depression: A Preliminary Magnetic Resonance Spectroscopy Study at 7T. J Affect Disord. 2021;279:417–25.

104. Sozeri-Varma G, Kalkan-Oguzhanoglu N, Efe M, Kiroglu Y, Duman T. Neurochemical metabolites in prefrontal cortex in patients with mild/moderate levels in first-episode depression. Neuropsychiatr Dis Treat. 2013;9:1053–9.

105. Tae WS, Kim SS, Lee KU, Nam EC, Koh SH. Progressive decrease of N-acetylaspartate to total creatine ratio in the pregenual anterior cingulate cortex in patients with major depressive disorder: longitudinal 1H-MR spectroscopy study. Acta Radiol. 2014;55(5):594–603.

106. Taylor MJ, Godlewska BR, Norbury R, Selvaraj S, Near J, Cowen PJ. Early increase in marker of neuronal integrity with antidepressant treatment of major depression: 1H-magnetic resonance spectroscopy of N-acetyl-aspartate. Int J Neuropsychopharmacol. 2012;15(10):1541–6.

107. Tosun S, Tosun M, Akansel G, Gokbakan AM, Unver H, Tural U. Proton magnetic resonance spectroscopic analysis of changes in brain metabolites following electroconvulsive therapy in patients with major depressive disorder. Int J Psychiatry Clin Pract. 2020;24(1):96–101.

108. Vythilingam M, Charles HC, Tupler LA, Blitchington T, Kelly L, Krishnan KR. Focal and lateralized subcortical abnormalities in unipolar major depressive disorder: an automated multivoxel proton magnetic resonance spectroscopy study. Biol Psychiatry. 2003;54(7):744–50.

109. Wang Y, Jia Y, Xu G, Ling X, Liu S, Huang L. Frontal white matter biochemical abnormalities in first-episode, treatment-naive patients with major depressive disorder: a proton magnetic resonance spectroscopy study. J Affect Disord. 2012;136(3):620–6.

110. Zavorotnyy M, Zollner R, Rekate H, Dietsche P, Bopp M, Sommer J, et al. Intermittent theta-burst stimulation moderates interaction between increment of N-Acetyl-Aspartate in anterior cingulate and improvement of unipolar depression. Brain Stimul. 2020;13(4):943–52.

111. Zhang Y, Han Y, Wang Y, Zhang Y, Li L, Jin E, et al. A MRS study of metabolic alterations in the frontal white matter of major depressive disorder patients with the treatment of SSRIs. BMC Psychiatry. 2015;15:99.

112. Zhong S, Wang Y, Zhao G, Xiang Q, Ling X, Liu S, et al. Similarities of biochemical abnormalities between major depressive disorder and bipolar depression: a proton magnetic resonance spectroscopy study. J Affect Disord. 2014;168:380–6.

113. Sendur I, Kalkan Oguzhanoglu N, Sozeri Varma G. Study on Dorsolateral Prefrontal Cortex Neurochemical Metabolite Levels of Patients with Major Depression Using H-MRS Technique. Turk Psikiyatri Derg. 2020;31(2):75–83.

114. Mulinari S. Monoamine theories of depression: historical impact on biomedical research. J Hist Neurosci. 2012;21(4):366–92.

115. Duman RS, Malberg J, Nakagawa S, D’Sa C. Neuronal plasticity and survival in mood disorders. Biol Psychiatry. 2000;48(8):732–9.

116. Tartt AN, Mariani MB, Hen R, Mann JJ, Boldrini M. Dysregulation of adult hippocampal neuroplasticity in major depression: pathogenesis and therapeutic implications. Molecular Psychiatry. 2022;27(6):2689–99.

117. Ongür D, Drevets WC, Price JL. Glial reduction in the subgenual prefrontal cortex in mood disorders. Proc Natl Acad Sci U S A. 1998;95(22):13290–5.

118. Fales CL, Barch DM, Rundle MM, Mintun MA, Mathews J, Snyder AZ, et al. Antidepressant treatment normalizes hypoactivity in dorsolateral prefrontal cortex during emotional interference processing in major depression. J Affect Disord. 2009;112(1-3):206–11.

119. Zhong X, Pu W, Yao S. Functional alterations of fronto-limbic circuit and default mode network systems in first-episode, drug-naive patients with major depressive disorder: A meta-analysis of resting-state fMRI data. J Affect Disord. 2016;206:280–6.

120. Wang Y-m, Yang Z-y. Aberrant pattern of cerebral blood flow in patients with major depressive disorder: A meta-analysis of arterial spin labelling studies. Psychiatry Research: Neuroimaging. 2022;321:111458.

121. Maller JJ, Thomson RH, Rosenfeld JV, Anderson R, Daskalakis ZJ, Fitzgerald PB. Occipital bending in depression. Brain. 2014;137(Pt 6):1830–7.

122. Lee JS, Kang W, Kang Y, Kim A, Han K-M, Tae W-S, et al. Alterations in the Occipital Cortex of Drug-Naïve Adults With Major Depressive Disorder: A Surface-Based Analysis of Surface Area and Cortical Thickness. Psychiatry Investig. 2021;18(10):1025–33.

123. Li X, Wang J. Abnormal neural activities in adults and youths with major depressive disorder during emotional processing: a meta-analysis. Brain Imaging Behav. 2021;15(2):1134–54.

124. Peng W, Chen Z, Yin L, Jia Z, Gong Q. Essential brain structural alterations in major depressive disorder: A voxel-wise meta-analysis on first episode, medication-naive patients. J Affect Disord. 2016;199:114–23.

125. Du MY, Wu QZ, Yue Q, Li J, Liao Y, Kuang WH, et al. Voxelwise meta-analysis of gray matter reduction in major depressive disorder. Prog Neuropsychopharmacol Biol Psychiatry. 2012;36(1):11–6.

126. Greicius MD, Flores BH, Menon V, Glover GH, Solvason HB, Kenna H, et al. Resting-state functional connectivity in major depression: abnormally increased contributions from subgenual cingulate cortex and thalamus. Biol Psychiatry. 2007;62(5):429–37.

127. Hamilton JP, Etkin A, Furman DJ, Lemus MG, Johnson RF, Gotlib IH. Functional neuroimaging of major depressive disorder: a meta-analysis and new integration of base line activation and neural response data. Am J Psychiatry. 2012;169(7):693–703.

128. Liao Y, Huang X, Wu Q, Yang C, Kuang W, Du M, et al. Is depression a disconnection syndrome? Meta-analysis of diffusion tensor imaging studies in patients with MDD. J Psychiatry Neurosci. 2013;38(1):49–56.

129. Shen X, Reus LM, Cox SR, Adams MJ, Liewald DC, Bastin ME, et al. Subcortical volume and white matter integrity abnormalities in major depressive disorder: findings from UK Biobank imaging data. Sci Rep. 2017;7(1):5547.

130. Admon R, Pizzagalli DA. Dysfunctional Reward Processing in Depression. Curr Opin Psychol. 2015;4:114–8.

131. Berman MG, Peltier S, Nee DE, Kross E, Deldin PJ, Jonides J. Depression, rumination and the default network. Soc Cogn Affect Neurosci. 2011;6(5):548–55.

132. Smoski MJ, Felder J, Bizzell J, Green SR, Ernst M, Lynch TR, et al. fMRI of alterations in reward selection, anticipation, and feedback in major depressive disorder. J Affect Disord. 2009;118(1-3):69–78.

133. Rizvi SJ, Salomons TV, Konarski JZ, Downar J, Giacobbe P, McIntyre RS, et al. Neural response to emotional stimuli associated with successful antidepressant treatment and behavioral activation. J Affect Disord. 2013;151(2):573–81.

134. Keedwell PA, Andrew C, Williams SC, Brammer MJ, Phillips ML. A double dissociation of ventromedial prefrontal cortical responses to sad and happy stimuli in depressed and healthy individuals. Biol Psychiatry. 2005;58(6):495–503.

135. Zhou HX, Chen X, Shen YQ, Li L, Chen NX, Zhu ZC, et al. Rumination and the default mode network: Meta-analysis of brain imaging studies and implications for depression. Neuroimage. 2020;206:116287.

136. Piguet C, Karahanoglu FI, Saccaro LF, Van De Ville D, Vuilleumier P. Mood disorders disrupt the functional dynamics, not spatial organization of brain resting state networks. Neuroimage Clin. 2021;32:102833.

137. Hoult DI, Lauterbur PC. The sensitivity of the zeugmatographic experiment involving human samples. Journal of Magnetic Resonance (1969). 1979;34(2):425–33.

138. Barker PB, Hearshen DO, Boska MD. Single-voxel proton MRS of the human brain at 1.5T and 3.0T. Magn Reson Med. 2001;45(5):765–9.

139. Kuan E, Chen X, Du F, Ongur D. N-acetylaspartate concentration in psychotic disorders: T2-relaxation effects. Schizophr Res. 2021;232:42–4.

