## Supplementary information for "Proton magnetic resonance spectroscopy of N-acetyl aspartate in first depressive episode and chronic major depressive disorder: a systematic review and meta-analysis"

**Running title**

N-acetyl aspartate in depression: a meta-analysis

Table of Tables

Table of Figures

### **Supplementary Methods**

Supplementary Table 1. PRISMA guidelines for systematic reviews and meta-analysis

| **#** | **Section/topic** | **Checklist item and brief description of how the criteria were handled** | **Section, page** |
| --- | --- | --- | --- |
| **TITLE** | | | |
| 1 | Title | *Identify the report as a systematic review, meta-analysis, or both.*  The study has been identified as a systematic review and meta-analysis of the studies that assessed brain levels of NAA in MDD patients using ^1^H-MRS. | Title |
| **ABSTRACT** | | | |
| 2 | Structured summary | *Provide a structured summary including, as applicable: background; objectives; data sources; study eligibility criteria, participants and interventions; study appraisal and synthesis methods; results; limitations; conclusions and implications of key findings; systematic review registration number.*  All relevant information has been included in the abstract. | Abstract |
| **INTRODUCTION** | | | |
| 3 | Rationale | *Describe the rationale for the review in the context of what is already known.*  MDD is a common and serious mood disorder that must be recognized by all physicians and health professionals, since its progressive nature towards cognitive and functional decline is not unstoppable. Recent advances in ^1^H-MRS technique allowed an increasingly accurate measurement of brain neuro-metabolites, in particular NAA, considered a marker of integrity and metabolic function of neuron cells. The role of NAA as a marker and/or pathogenetic factor in MDD is still unclear, since previous studies showed discordant results. | Introduction |
| 4 | Objectives | *Provide an explicit statement of questions being addressed with reference to participants, interventions, comparisons, outcomes, and study design (PICOS).*  We aimed to conduct a meta-analysis of the studies that assessed brain levels of NAA in FED and cMDD using ^1^H-MRS. We tested the hypothesis that NAA concentration could be lower in different brain regions in FED and cMDD patients relative to healthy controls. | Introduction |
| **METHODS** | | | |
| 5 | Protocol and registration | *Indicate if a review protocol exists, if and where it can be accessed (e.g., Web address), and, if available, provide registration information including registration number.*  The protocol has been registered on PROSPERO (ID CRD 42020221050). | Methods, Search strategy and selection criteria |
| 6 | Eligibility criteria | *Specify study characteristics (e.g., PICOS, length of follow-up) and report characteristics (e.g., years considered, language, publication status) used as criteria for eligibility, giving rationale.*  We included: a) randomized and case-control studies written in English; b) recruiting adult subjects (age>18) with a diagnosis of MDD according to DSM or ICD criteria and healthy controls (HC); c) measuring NAA levels in MDD and HC using ^1^H-MRS. We excluded: a) non-original studies written in languages other than English; b) recruiting patients with diagnosis other than MDD or whose MDD was secondary to drugs or general medical conditions; c) conducted in children and adolescents; d) using ^1^H-MRS to investigate brain levels of other cerebral metabolites, such as Glu, Gln, mI, Cho, but not NAA. | Methods, Search strategy and selection criteria |
| 7 | Information sources | *Describe all information sources (e.g., databases with dates of coverage, contact with study authors to identify additional studies) in the search and date last searched.*  Two-step search strategy: 1) a systematic search on Scopus® and Web of Knowledge℠ databases extended until October, 2021; 2) manual search of the reference lists of the retrieved articles. | Methods, Search strategy and selection criteria |
| 8 | Search | *Present full electronic search strategy for at least one database, including any limits used, such that it could be repeated.*  The following terms were used: “*n-acetylaspartate* OR *naa* AND *major depressive disorder* OR *mdd* OR *depression”* | Methods, Search strategy and selection criteria |
| 9 | Study selection | *State the process for selecting studies (i.e., screening, eligibility, included in systematic review, and, if applicable, included in the meta-analysis).*  The identified articles were screened by title and abstract, and the full text of surviving articles were further inspected for eligibility against a priori defined inclusion and exclusion criteria. Study types different from original articles were excluded. | Methods, Search strategy and selection criteria, Fig. 1 |
| 10 | Data collection process | *Describe method of data extraction from reports (e.g., piloted forms, independently, in duplicate) and any processes for obtaining and confirming data from investigators.*  Data were independently extracted by MT and FT. The extracted data were cross-checked, and discrepancies were resolved by discussion between MT, FT and the independent arbiter ^1^. | Methods, Data extraction |
| 11 | Data items | *List and define all variables for which data were sought (e.g., PICOS, funding sources) and any assumptions and simplifications made.*  Extracted variables: author, publication year, sample size, brain region, NAA mean, NAA standard deviation or standard error, p value, socio-demographic moderators (age, % female), methodological moderators (^1^H-MRS field strength and acquisition sequence, NAA quantification, cefalo spinal fluid (CSF) correction, time echo, time relaxation) and clinical moderators (illness duration, depression symptom severity, antidepressant treatment). | Methods, Data extraction |
| 12 | Risk of bias in individual studies | *Describe methods used for assessing risk of bias of individual studies (including specification of whether this was done at the study or outcome level), and how this information is to be used in any data synthesis.*  Risk of bias was assessed with the Newcastle-Ottawa Quality Assessment Scale ^2^. | Methods, Data analysis |
| 13 | Summary measures | *State the principal summary measures (e.g., risk ratio, difference in means).*  Hedges’ g (standardized mean difference) and relative standard error. | Methods, Data analysis |
| 14 | Synthesis of results | *Describe the methods of handling data and combining results of studies, if done, including measures of consistency (e.g., I^2^) for each meta-analysis.*  Effect sizes were pooled using a random-effect model; heterogeneity and bias were assessed by performing influence analyses with the leave-one-out method and the Graphic Display of Heterogeneity (GOSH) plot analysis; moderators were tested with subgroup analyses and meta-regressions for categorical and continuous moderators, respectively. | Methods, Data analysis |
| 15 | Risk of bias across studies | *Specify any assessment of risk of bias that may affect the cumulative evidence (e.g., publication bias, selective reporting within studies).*  Publication bias was assessed by plotting funnel plots and p-curves, with the Egger’s test to quantify funnel plot asymmetry. | Methods, Data analysis |
| 16 | Additional analyses | *Describe methods of additional analyses (e.g., sensitivity or subgroup analyses, metaregression), if done, indicating which were pre-specified.*  Outlier analysis. Sensitivity analysis with *leave-one-out* method. Subgroup analysis for categorical moderators: ^1^H-MRS acquisition Sequence; NAA quantification (Cr scaling vs absolute concentration); CSF quantification (yes vs no); antidepressant treatment (yes vs no); field strength (1.5 vs > 1.5). Meta-regression models to investigate the influence of pre-specified continuous predictors: year of publication, age, % female, ¹H-MRS field strength, time echo, time relaxation, illness duration and Ham-D score. | Methods, Data analysis, supplementary influence diagnostics |
| **RESULTS** | | | |
| 17 | Study selection | *Give numbers of studies screened, assessed for eligibility, and included in the review, with reasons for exclusions at each stage, ideally with a flow diagram.*  All details are depicted in the PRISMA flow-chart and described in the main text. | Results; Fig. 1 |
| 18 | Study characteristics | *For each study, present characteristics for which data were extracted (e.g., study size, PICOS, follow-up period) and provide the citations.*  Characteristics of the included studies are presented in Table 1. | Results; Table 1 |
| 19 | Risk of bias within studies | *Present data on risk of bias of each study and, if available, any outcome level assessment (see item 12).*  The results of quality assessment with NOS are reported in table 9. | Results; Table 9 |
| 20 | Results of individual studies | *For all outcomes considered (benefits or harms), present, for each study: (a) simple summary data for each intervention group (b) effect estimates and confidence intervals, ideally with a forest plot.*  Results of individual studies, in terms of Hedges’ g, standard error, 95%CI, and weight, are represented in the forest plots of each brain area section and described in the Result section. | Results |
| 21 | Synthesis of results | *Present results of each meta-analysis done, including confidence intervals and measures of consistency.*  Results of the meta-analysis, in terms of overall Hedges’ g, 95%CI, prediction interval, and measures of consistency, are represented in Table.2 and Fig.2 and described in the Result section. Results of the meta-analyses conducted for each illness phase and brain region, in terms of Hedges’ g, 95%CI, prediction interval, and measures of consistency, are reported in each brain area section. | Results; Table.2; Fig. 2 |
| 22 | Risk of bias across studies | *Present results of any assessment of risk of bias across studies (see Item 15).*  Funnel plots for publication bias are shown in each brain area section. Results of Egger’s test for publication bias, when present, are reported in each brain area section. | Results |
| 23 | Additional analysis | *Give results of additional analyses, if done (e.g., sensitivity or subgroup analyses, meta-regression [see Item 16]).*  Results of outlier and sensitivity analysis are presented in each brain area section. Results of meta-regressions are reported in each brain area section. | Results |
| **DISCUSSION** | | | |
| 24 | Summary of evidence | *Summarize the main findings including the strength of evidence for each main outcome; consider their relevance to key groups (e.g., healthcare providers, users, and policy makers).* | Discussion |
| 25 | Limitations | *Discuss limitations at study and outcome level (e.g., risk of bias), and at review-level (e.g., incomplete retrieval of identified research, reporting bias).* | Discussion |
| 26 | Conclusions | *Provide a general interpretation of the results in the context of other evidence, and implications for future research.* | Discussion |
| **FUNDING** | | | |
| 27 | Funding | This work was supported by the University of Pisa, Fondi di Ateneo 2019 to G.R. G.R. is supported by the European Union’s Horizon 2020 research and innovation program under the MSC grant agreement n° 101026235. | Acknowledgements |

Supplementary Table 2. Search strategy according to the Population, Intervention, Comparison, Outcomes and Study Design (PICOS) model

| Parameter | Inclusion criteria | Exclusion criteria |
| --- | --- | --- |
| Population | Patients aged 18 years or older  with a diagnosis of MDD as assessed by DSM, ICD, or consensus expert evaluation. | Children or adolescents;  diagnosis of any other mental disorder, particularly bipolar disorder, schizophrenia, schizoaffective disorder. |
| Interventions | Brain levels of NAA measured with ^1^H-MRS. | Brain levels of other cerebral metabolites, such as glutamate, glutamine, myo-inositol, choline. |
| Comparison | Comparison between patients with MDD and healthy control subjects. Controls can also be subjects with a non-psychiatric illness. | Comparison between patients with a diagnosis of MDD and patients with any other mental disorder, i.e. the absence of a healthy control group. |
| Outcomes | Measures of NAA concentrations in different brain regions (both absolute or scaled to creatine). | Measures of NAA T2 relaxation times. |
| Study design model | Cross-sectional and randomized controlled trials (RCT). In the case of RCT, we used the NAA measures prior to treatment allocation. | Case studies, case series, pilot studies and reviews. |

#### Data extraction

We extracted sample size, mean NAA concentration and standard deviation or standard error of the mean for patients with MDD patient and healthy controls. If the normality assumption allowed parametric statistics in the original paper, t-test or p-value were extracted alongside with direction of the effect size. In this case, we adopted the following a priori rules:

- Statistically significant differences in p-value were implied as 0.05, where not directly specified.
- Not statistically significant differences in p-value were implied as 0.90 where not directly specified.
- Null hypothesis significance tests were considered two-tailed if not otherwise specified.

If non-parametric statistic were used we computed the effect size from the z-value provided in the paper as follow ^3^:

$$r=\frac{z}{\sqrt{N}}$$

Where *r* is the effect size, *z* is the z-score and *N* is the sample size.

Supplementary Table 3. Merging of continuous variables

|  | **Subgroup 1** | **Subgroup 2** | **Combined group** |
| --- | --- | --- | --- |
| **Sample size** | N1 | N2 | *N1+ N2* |
| **Mean** | M1 | M2 | $\frac{N1M1+N2M2}{N1+N2}$ |
| **SD** | SD1 | SD2 | $\sqrt[2]{\frac{\left( N\text{1}-1 \right){SD1}^{2}+\left( N2-1 \right){SD2}^{2}+ \frac{N1N2}{N1+N2}({M1}^{2}+{M2}^{2}-2M1M2)}{N1+N2-1}}$ |

Abbreviations: M, mean; N, sample size; SD, standard deviation.

### **Supplementary Results**

#### 1. Frontal lobe

##### Main results

We included 26 studies: 517 patients and 452 controls.

Supplementary Figure 1. Forest plot of all studies examining NAA levels in the frontal lobe of patients with cMDD compared to healthy controls.

The meta-analysis revealed a significant difference between patients and controls (n = 26, Hedges’ g = -0.33, 95% CI -0.598 to -0.062; p = 0.018; Q = 102.84, I² = 75.7%, p < 0.0001). Positive values favour cMDD, while negative values favour controls.

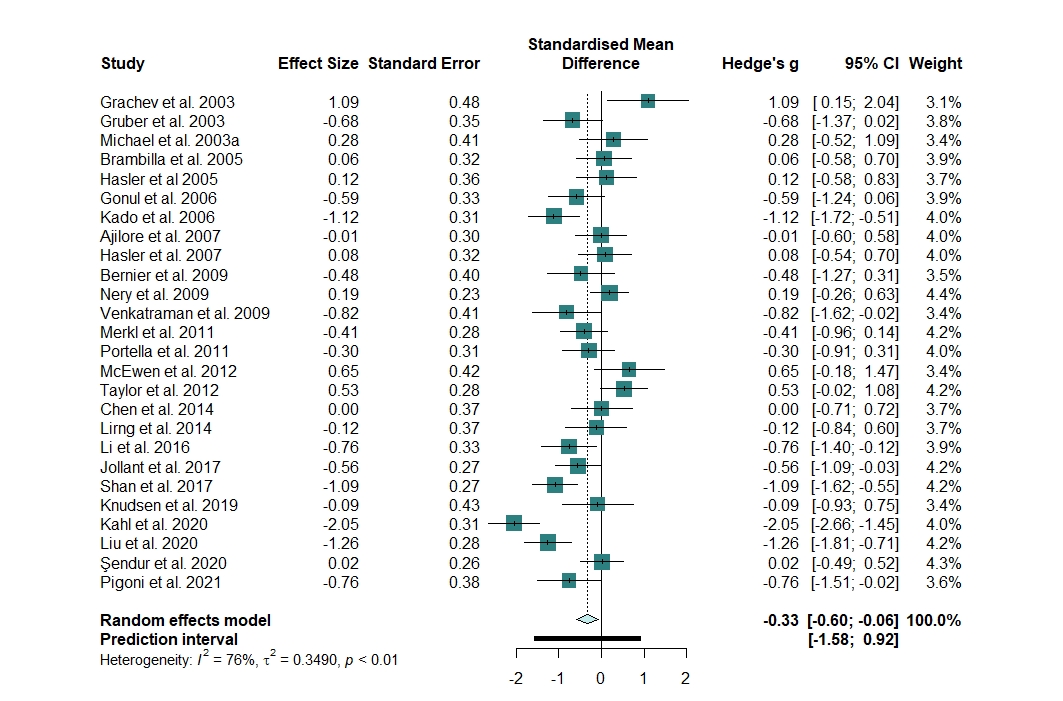

##### Heterogeneity

We identified and removed four outliers ^4-7^.

The updated meta-analysis confirmed a significant difference between patients and controls (n = 22, Hedges’ g = -0.300, 95% CI -0.505 to -0.094; p = 0.006), with significant moderate heterogeneity (Q = 42.84, *I²* = 51%, p = 0.003).

Supplementary Figure 2. Heterogeneity assessment in the frontal lobe meta-analysis

**A.** Baujat plot of the contribution of each study to the overall heterogeneity (as measured by Cochran’s Q) and its influence on the pooled effect size. **B.** Influence analysis with the *leave-one-out* method of the meta-analysis of all studies of cMDD examining NAA levels in the frontal lobe. Parameters of the influence analysis: standardized residuals, dffits, Cook’s distance, covariance ratio, tau^2^, Q, hat, and weight. No studies were identified as influential according to the cut-offs proposed by Viechtbauer and Cheung ^8^. **C.** Forest plot of the overall effect sizes of the meta-analyses of all studies of cMDD examining NAA levels in the frontal lobe, recalculated with the *leave-one-out* method, ordered by effect size. **D.** Forest plot of the overall effect sizes of the meta-analyses of all studies of cMDD examining NAA levels in the frontal lobe, recalculated with the *leave-one-out* method, ordered by heterogeneity.

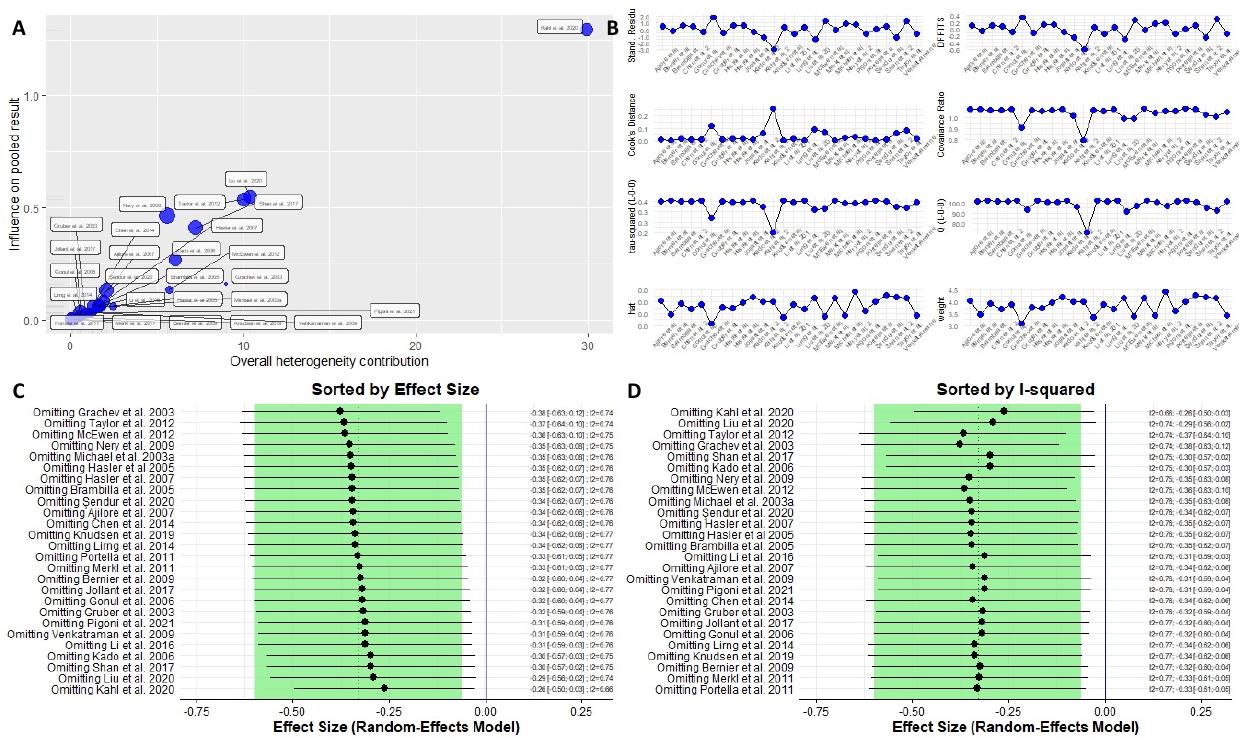

Supplementary Figure 3. GOSH diagnostics in the frontal lobe meta-analysis

**A**. GOSH plot showing the meta-analysis models of all studies of cMDD examining NAA levels in the frontal lobe, fitted to all 2^k-1^ possible combinations of the included studies (x-axis, pooled effect size; y-axis, between-study heterogeneity). The GOSH plot highlighted two distinct clusters, one with greater heterogeneity and more negative effect size, and the other with lower heterogeneity and smaller effect size **B.** DBSCAN Algorithm. **C.** K.means Algorithm. The GOSH diagnostics confirmed the presence of 4 studies with a large influence on the effect size and heterogeneity, the same previously identified as outliers (2-4, 7) **D.** GOSH plot fitted to all 2^k-1^ possible combinations of the included studies with and without the outlier with most extreme effect size ^5^.

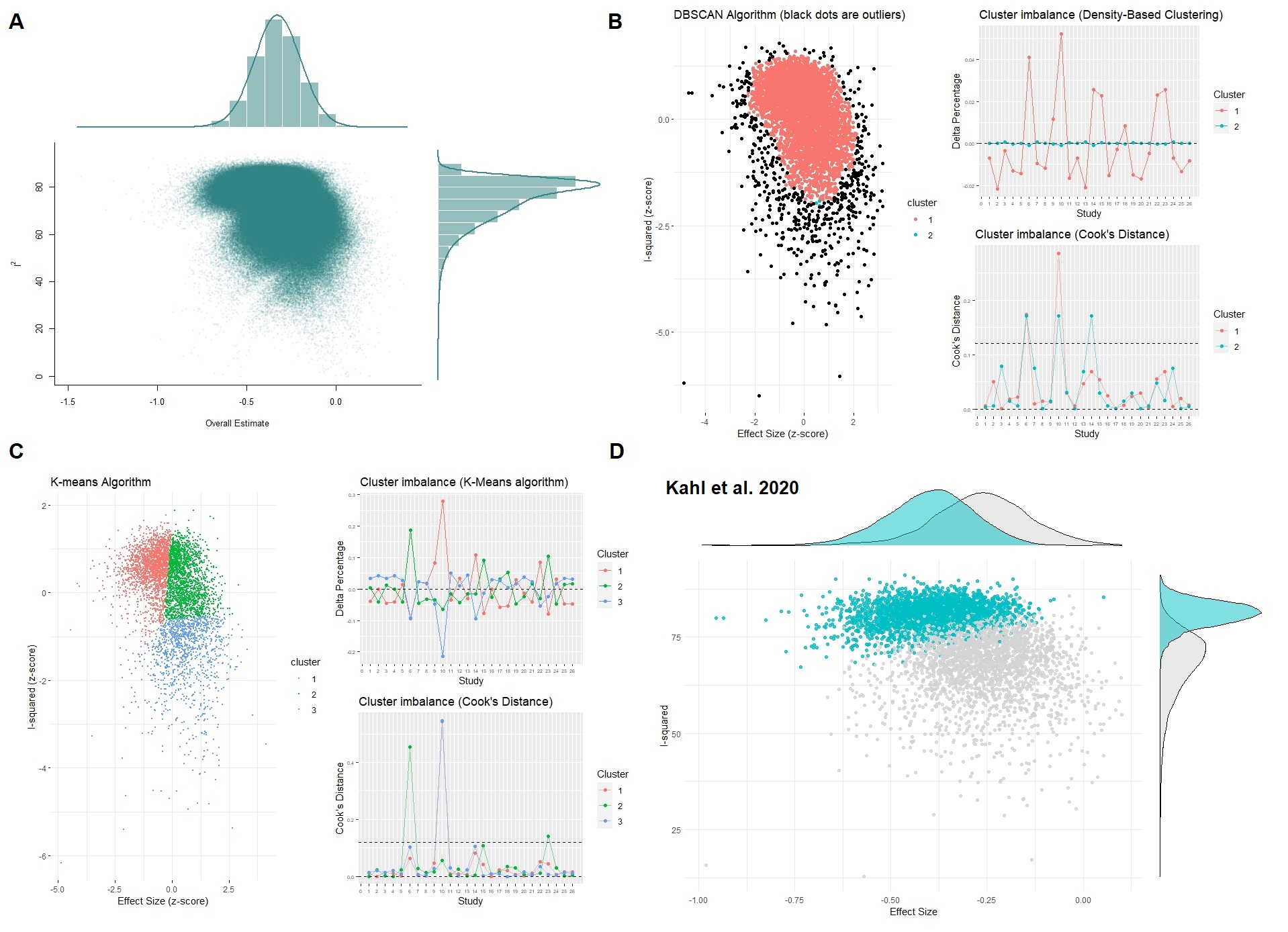

##### Subgroup Analyses

Supplementary Table 4. Subgroup analysis of the meta-analyses of all studies examining NAA levels in the frontal lobe of patients with cMDD compared to healthy controls.

|  | Subgroup sample size | Hedges’ g | 95% CI | p | I² | 95% CI | p subgroups |
| --- | --- | --- | --- | --- | --- | --- | --- |
| NAA quantification method | | | | | | | 0.717 |
| Cr scaling | 11 | -0.384 | -0.803, 0.034 | 0.072 | 78% | 62, 88% |  |
| Absolute | 15 | -0.286 | -0.616, 0.044 | 0.089 | 74% | 57, 85% |  |
| CSF correction | | | | | | | 0.757 |
| Yes | 11 | -0.380 | -0.840, 0.080 | 0.105 | 83% | 70, 90% |  |
| No | 15 | -0.294 | -0.592, 0.005 | 0.054 | 69% | 46, 82% |  |
| Field strength | | | | | | | 0.297 |
| 1.5 T | 12 | -0.185 | -0.504, 0.134 | 0.255 | 61% | 26, 79% |  |
| > 1.5 T | 14 | -0.450 | -0.833, -0.068 | 0.021 | 81% | 69, 88% |  |

##### Metaregressions

Supplementary Table 5. Results of the meta-regressions in the frontal lobe.

|  | beta | SE | p |
| --- | --- | --- | --- |
| Age | -0.0003 | 0.0114 | 0.982 |
| Female % | 0.005 | 0.0093 | 0.587 |
| Illness duration | 0.0563 | 0.0379 | 0.172 |
| Ham-D scale | -0.0012 | 0.0167 | 0.9421 |
| ^1^H-MRS field strength | -0.1807 | 0.174 | 0.3094 |
| TE | -0.0009 | 0.0023 | 0.7160 |
| TR | 0.0001 | 0.0001 | 0.5253 |

Supplementary Figure 4. Meta-regression assessing the relationship between publication year and Hedges’ g of the meta-analysis of all studies examining NAA levels in the frontal lobe of patients with cMDD as compared to healthy controls.

Intercept = 99.009, β = -0.049, p = 0.025; R² = 15.92%

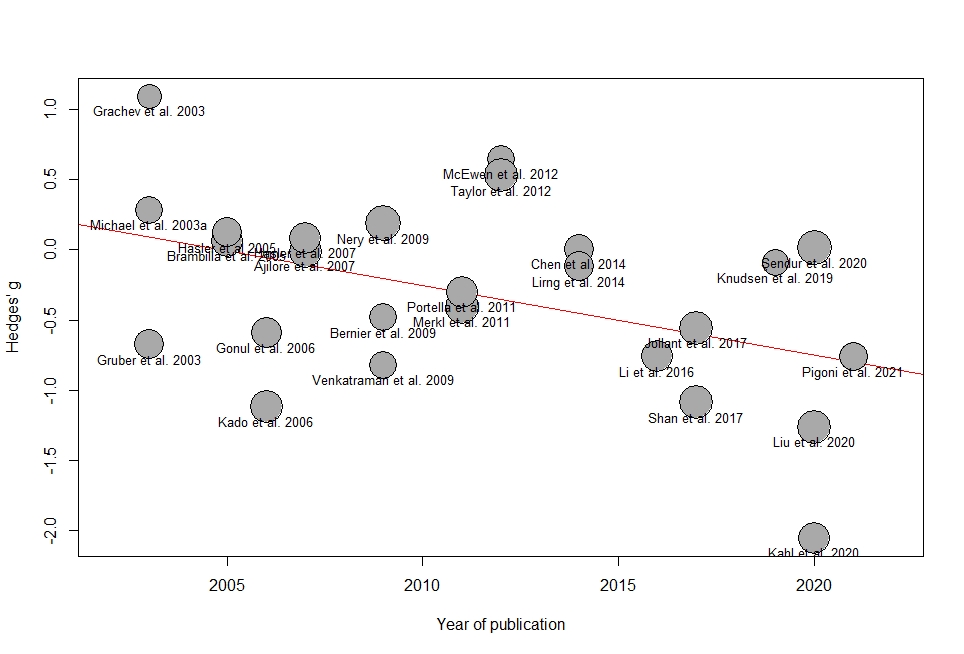

##### Publication bias

Supplementary Figure 5. Assessment of small sample publication bias with the contour-enhanced funnel plot for the studies examining NAA levels in the frontal lobe of cMDD.

The Egger’s test yielded non-significant results (Intercept = 1.651, p = 0.479).

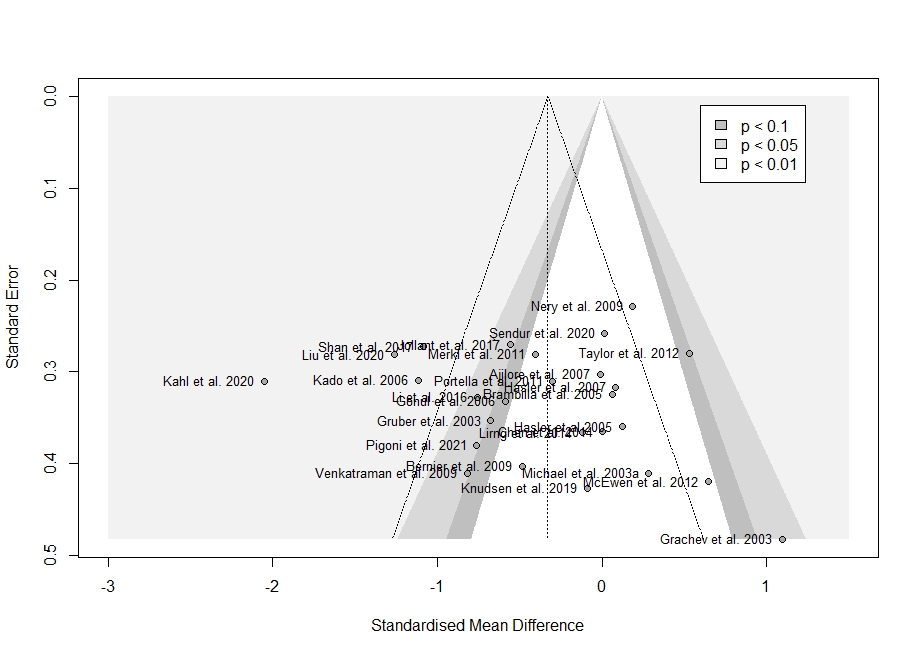

Supplementary Figure 6. P-curve for the analysis of cMDD in the frontal lobe.

The analysis of the p-curve confirmed the presence of evidential value: the right-skewness test was significant for both the full and half curve, while the flatness test was not significant for both the full and half curve. Overall, the p-curve appears right-skewed, since highly significant results (p-values = 0.01) are over-represented. This can be interpreted as evidence of no p-hacking nor selective reporting.

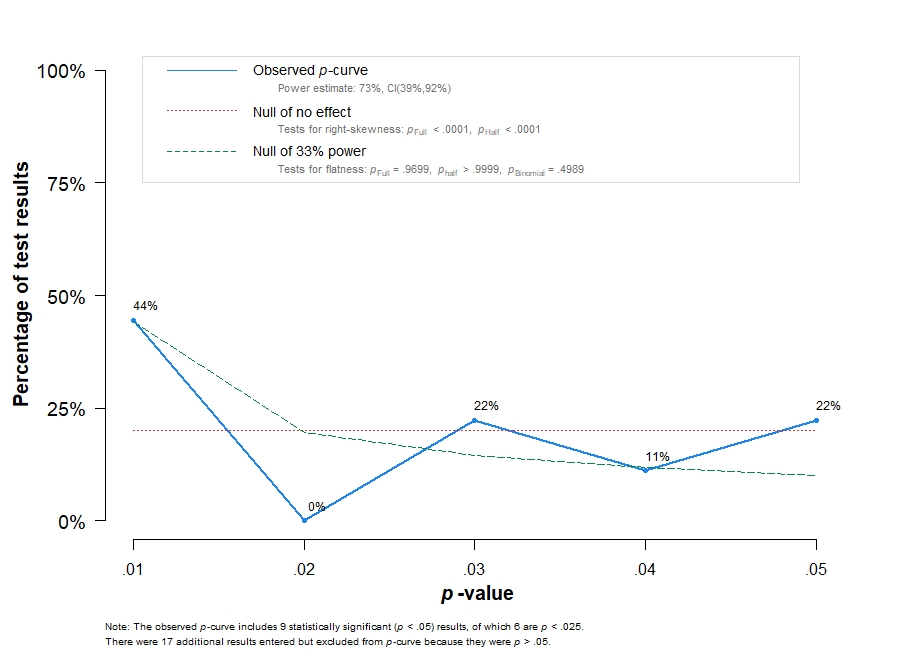

#### 2. Dorsolateral prefrontal region

##### Main results

We included 13 studies: 259 patients and 275 controls.

Supplementary Figure 7. Forest plot of all studies examining NAA levels in the dorsolateral prefrontal region of patients with cMDD compared to healthy controls.

The meta-analysis revealed no significant difference between patients and controls (n = 13, Hedges’ g = -0.024, 95% CI -0.274 to 0.225; p = 0.836; Q = 17.63, I² = 31.9%, p = 0.127). Positive values favour cMDD, while negative values favour controls.

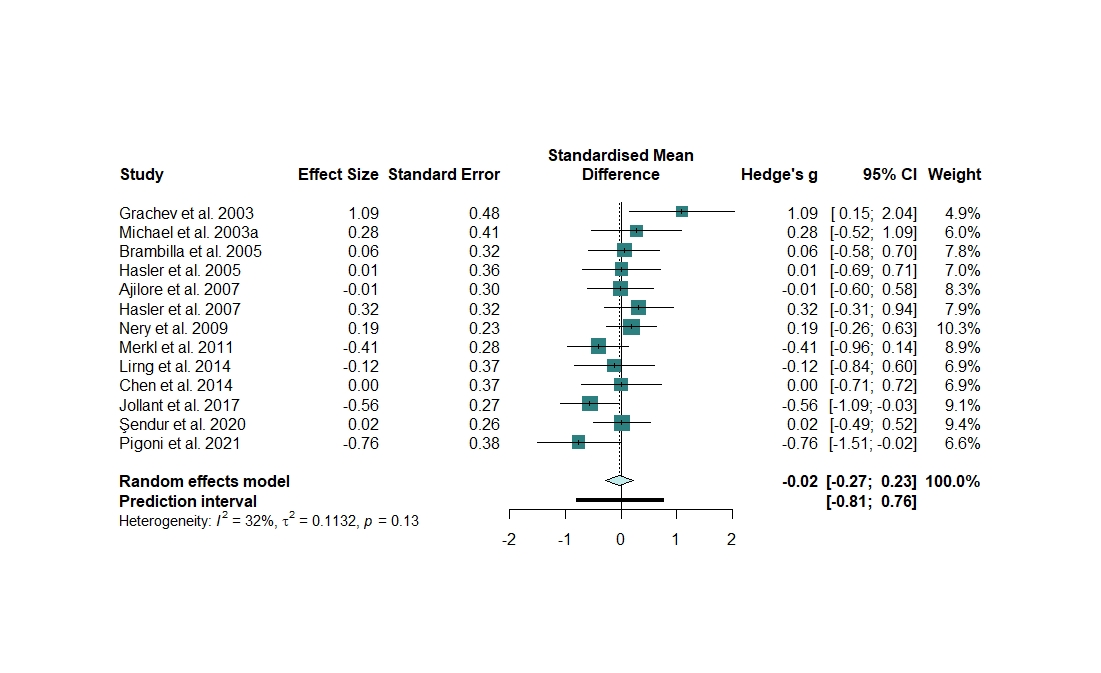

##### Heterogeneity

No outliers could be found and removed.

Between-study heterogeneity was not significant (Q = 17.63, I² = 31.9%, p = 0.127).

Supplementary Figure 8. Heterogeneity assessment in the dorsolateral prefrontal region meta-analysis

**A.** Influence analysis with the *leave-one-out* method of the meta-analysis of all studies of cMDD examining NAA levels in the dorsolateral prefrontal region. Parameters of the influence analysis: standardized residuals, dffits, Cook’s distance, covariance ratio, tau^2^, Q, hat, and weight. No studies were identified as influential according to the cut-offs proposed by Viechtbauer and Cheung ^8^. **C.** Forest plot of the overall effect sizes of the meta-analyses of all studies of cMDD examining NAA levels in the dorsolateral prefrontal region, recalculated with the *leave-one-out* method, ordered by effect size. **D.** Forest plot of the overall effect sizes of the meta-analyses of all studies of cMDD examining NAA levels in the dorsolateral prefrontal region, recalculated with the *leave-one-out* method, ordered by heterogeneity.

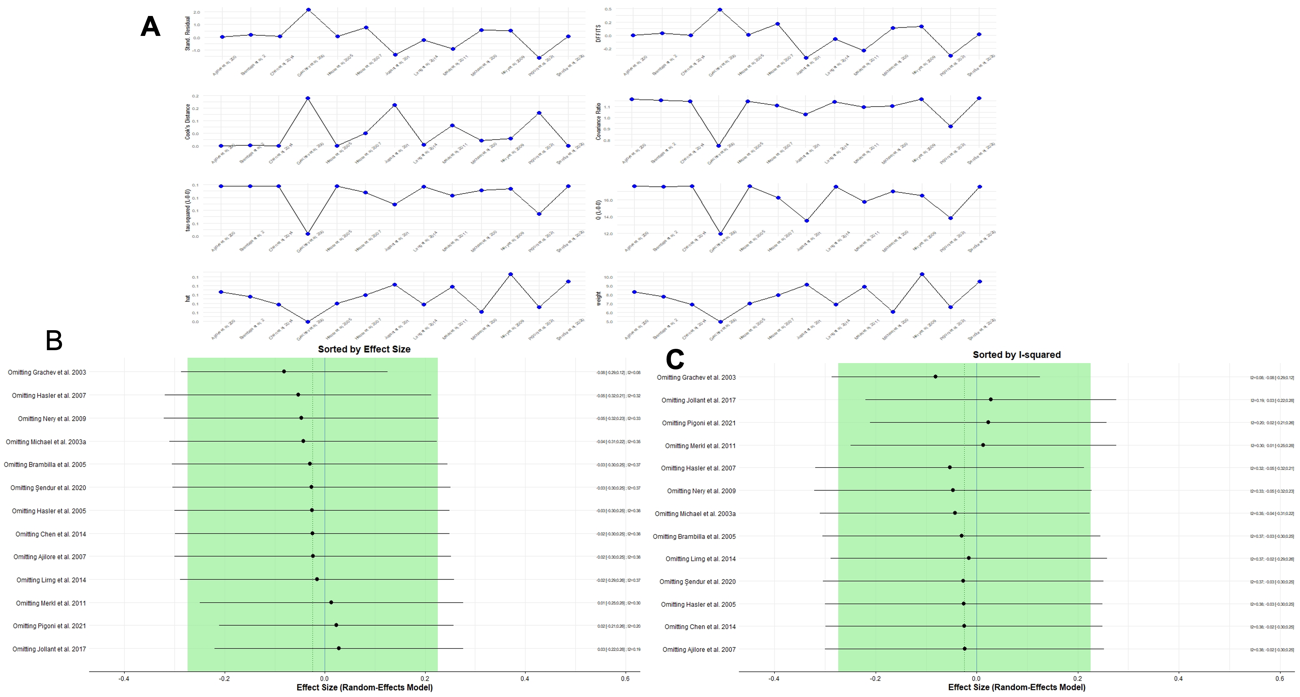

Supplementary Figure 9. GOSH diagnostics in the dorsolateral prefrontal region meta-analysis

**A**. GOSH plot showing the meta-analysis models of all studies of cMDD examining NAA levels in the dorsolateral prefrontal region, fitted to all 2^k-1^ possible combinations of the included studies (x-axis, pooled effect size; y-axis, between-study heterogeneity). The GOSH plot highlighted two distinct clusters, one with higher heterogeneity and slightly positive effect sizes, and the other one with lower heterogeneity and slightly negative effect sizes. **B.** K.means Algorithm. **C.** Gaussian Mixture Model. The GOSH diagnostics detected the presence of Grachev et al., 2003 ^4^ as a study with an influence on the effect size and heterogeneity. **D.** GOSH plot fitted to all 2^k-1^ possible combinations of the included studies with and without the outlier with most extreme effect size ^4^.

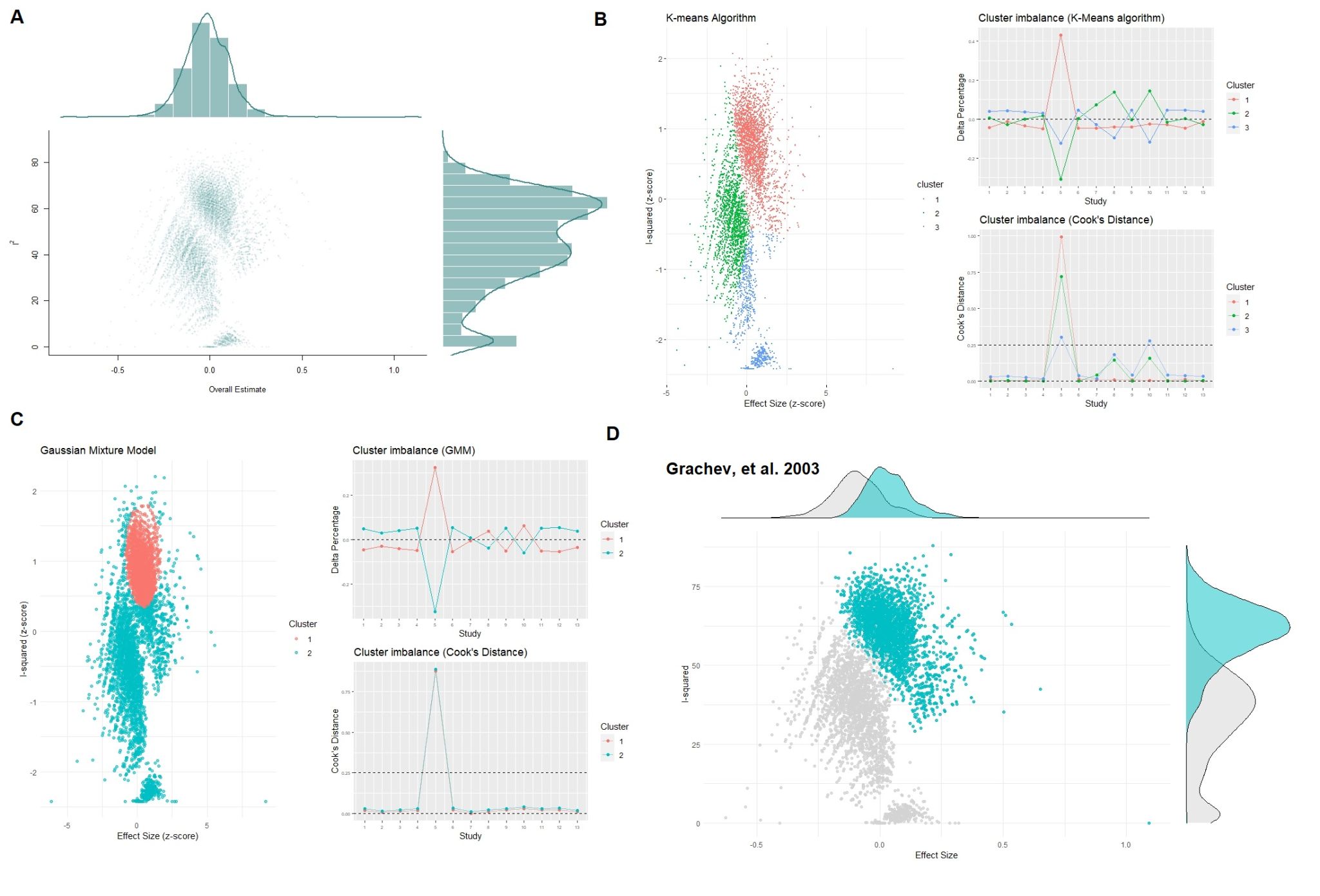

##### Subgroup Analyses

Supplementary Table 6. Subgroup analysis of the meta-analyses of all studies examining NAA levels in the dorsolateral prefrontal region of patients with cMDD compared to healthy controls.

|  | Subgroup sample size | Hedges’ g | 95% CI | p | I² | 95% CI | p subgroups |
| --- | --- | --- | --- | --- | --- | --- | --- |
| NAA quantification method | | | | | | | 0.716 |
| Cr scaling | 5 | 0.019 | -0.475, 0.513 | 0.939 | 57% | 0, 84% |  |
| Absolute | 7 | -0.062 | -0.325, 0.201 | 0.643 | 22% | 0, 65% |  |
| CSF correction | | | | | | | 0.901 |
| Yes | 5 | -0.003 | -0.239, 0.233 | 0.980 | 0% | 0, 74% |  |
| No | 8 | -0.030 | -0.390, 0.329 | 0.869 | 51% | 0, 78% |  |
| Field strength | | | | | | | 0.248 |
| 1.5 T | 8 | 0.077 | -0.233, 0.387 | 0.626 | 30% | 0, 69% |  |
| > 1.5 T | 5 | -0.182 | -0.495, 0.131 | 0.253 | 25% | 0, 70% |  |

##### Metaregressions

Supplementary Table 7. Results of the meta-regressions in the dorsolateral prefrontal region.

|  | beta | SE | p |
| --- | --- | --- | --- |
| Age | 0.0121 | 0.0180 | 0.5218 |
| Female % | -0.0243 | 0.014 | 0.1413 |
| Illness duration | -0.0144 | 0.0518 | 0.794 |
| Ham-D scale | -0.0504 | 0.0149 | 0.28 |
| ^1^H-MRS field strength | -0.2902 | 0.1888 | 0.1628 |
| TE | -0.0015 | 0.0037 | 0.6975 |
| TR | -0.0001 | -0.0001 | 0.6092 |

Supplementary Figure 10. Meta-regression assessing the relationship between publication year and Hedges’ g of the meta-analysis of all studies examining NAA levels of all studies examining NAA levels in the dorsolateral prefrontal region of patients with cMDD as compared to healthy controls.

Intercept = 89.156, β = -0.044, p = 0.014; R² = 41.60%

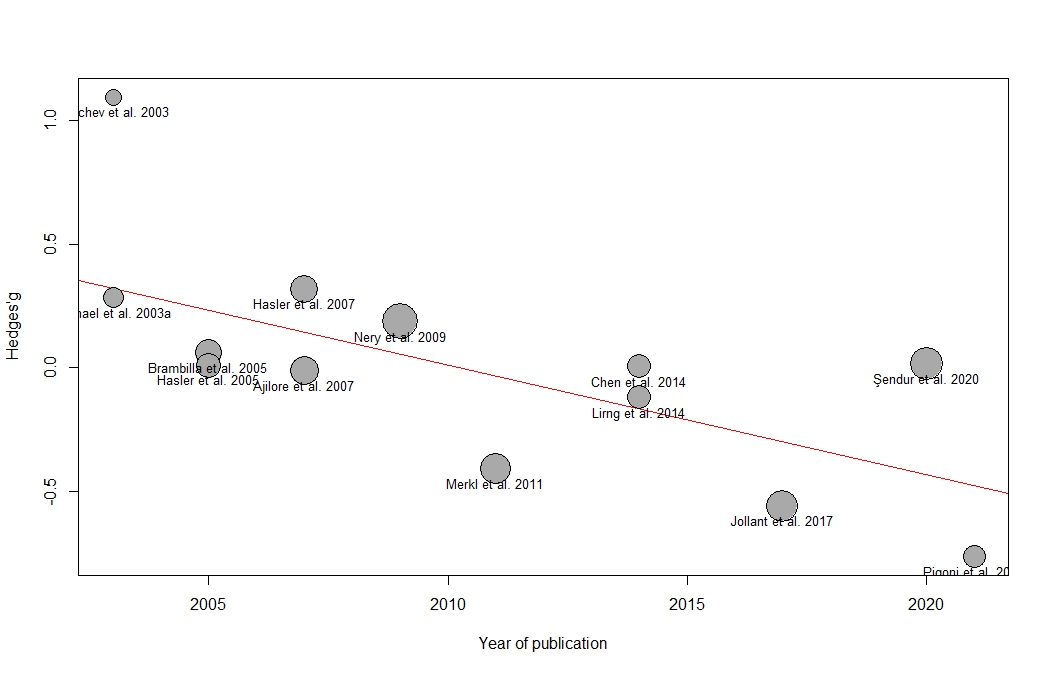

##### Publication bias

Supplementary Figure 11. Assessment of small sample publication bias with the contour-enhanced funnel plot for the studies examining NAA levels in the dorsolateral prefrontal region of cMDD.

The Egger’s test yielded non-significant results (Intercept = 1.534, p = 0.399).

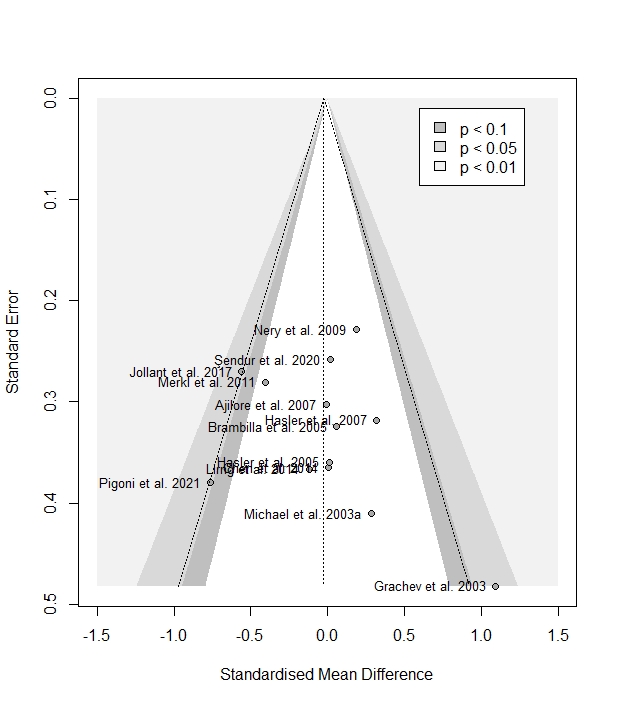

Supplementary Figure 12. P-curve for the analysis of cMDD in the dorsolateral prefrontal region.

The analysis of the p-curve confirmed that evidential value was absent: the right-skewness test was not significant for neither the full and half curve, while the flatness test was significant with p < 0.05 for the full curve. Overall, the p-curve appears left-skewed, since p-values close to 0.05 are over-represented, while highly significant results (p-values < 0.03) are poorly represented. This can be interpreted as evidence of p-hacking and selective reporting.

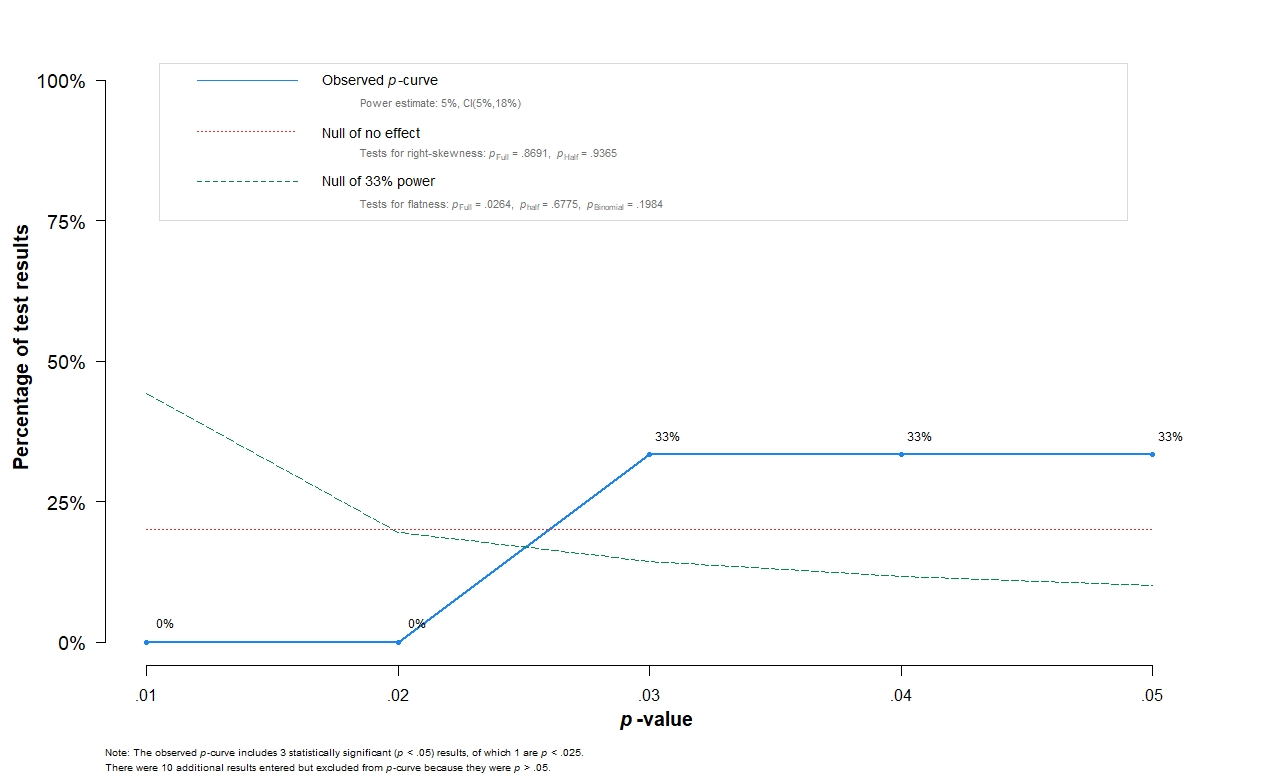

##### Sensitivity analysis on the influence of geriatric populations

The sensitivity analysis omitting one study including geriatric subjects ^9^ revealed no difference between patients and controls (n = 6, Hedges’ g = 0.007, 95% CI -0.560 to 0.573; p = 0.977), with a significant between-study heterogeneity (Q = 13.25, I² = 62.3%, p = 0.021).

#### 3. Prefrontal cortex

##### Main results

We included 4 studies: 82 patients and 89 controls.

Supplementary Figure 13. Forest plot of all studies examining NAA levels in the prefrontal cortex of patients with cMDD compared to healthy controls.

The meta-analysis revealed no difference between patients and controls (n = 4, Hedges’ g = -0.801, 95% CI -1.644 to -0.043; p = 0.057; Q = 6.79, I² = 55.8%, p = 0.079). Positive values favour cMDD, while negative values favour controls.

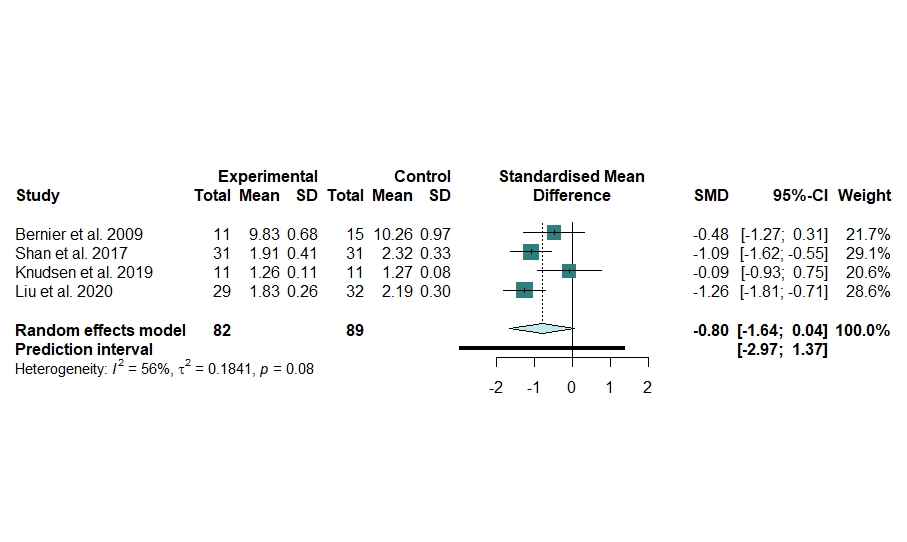

##### Heterogeneity

No outlier was identified.

The between-study heterogeneity was not significant (*Q* = 6.79, *I²* = 55.8%, p = 0.079).

##### Publication bias

Supplementary Figure 14. Assessment of small sample publication bias with the contour-enhanced funnel plot for the studies examining NAA levels in the prefrontal cortex of cMDD.

Egger’s test was not performed because there were not enough studies.

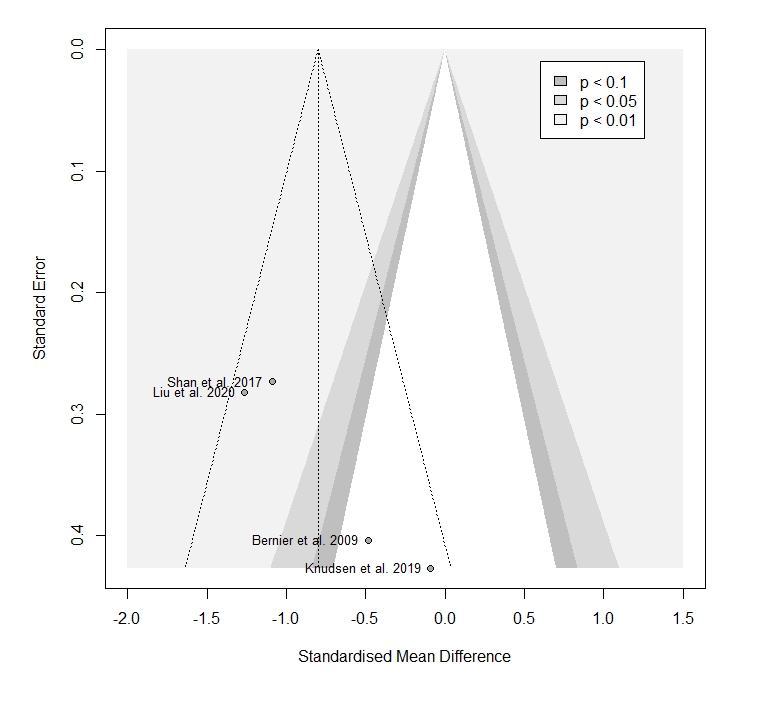

#### 4. Medial prefrontal cortex

##### Main results

We included 7 studies: 156 patients and 114 controls.

Supplementary Figure 15. Forest plot of all studies examining NAA levels in the medial prefrontal cortex of patients with cMDD compared to healthy controls.

The meta-analysis revealed no difference between patients and controls (n = 7, Hedges’ g = -0.096, 95% CI -0.632 to 0.440; p = 0.677; Q = 16.81, I^2^ = 64.3%, p = 0.01). Positive values favour cMDD, while negative values favour controls.

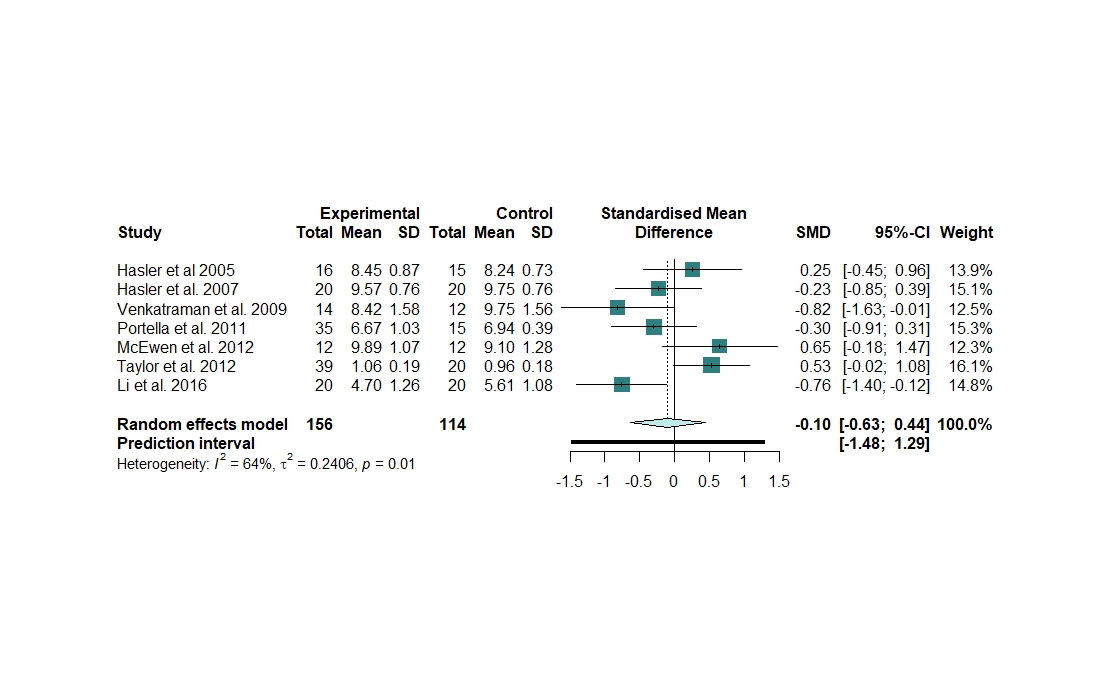

##### Heterogeneity

No outlier was identified.

The between-study heterogeneity was significant and moderate (Q = 16.81, I^2^ = 64.3%, p = 0.01).

Supplementary Figure 16. Heterogeneity assessment in the medial prefrontal cortex meta-analysis

**A.** Forest plot of the overall effect sizes of the meta-analyses of all studies of cMDD examining NAA levels in the medial prefrontal cortex, recalculated with the *leave-one-out* method, ordered by effect size. **B.** Forest plot of the overall effect sizes of the meta-analyses of all studies of cMDD examining NAA levels in the medial prefrontal cortex, recalculated with the *leave-one-out* method, ordered by heterogeneity.

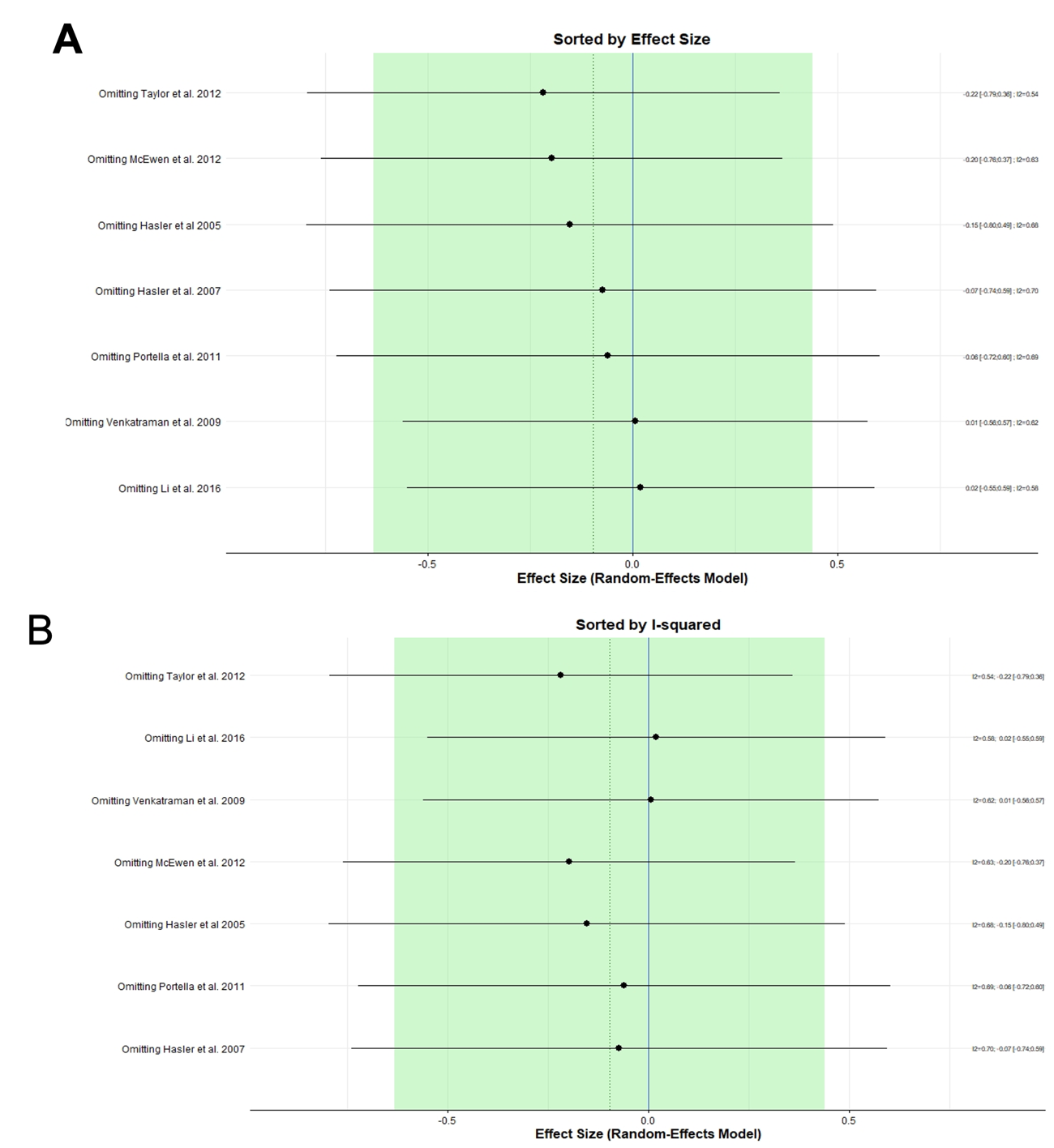

##### Publication bias

Supplementary Figure 17. Assessment of small sample publication bias with the contour-enhanced funnel plot for the studies examining NAA levels in the medial prefrontal cortex of cMDD.

Egger’s test was not performed because there were not enough studies.

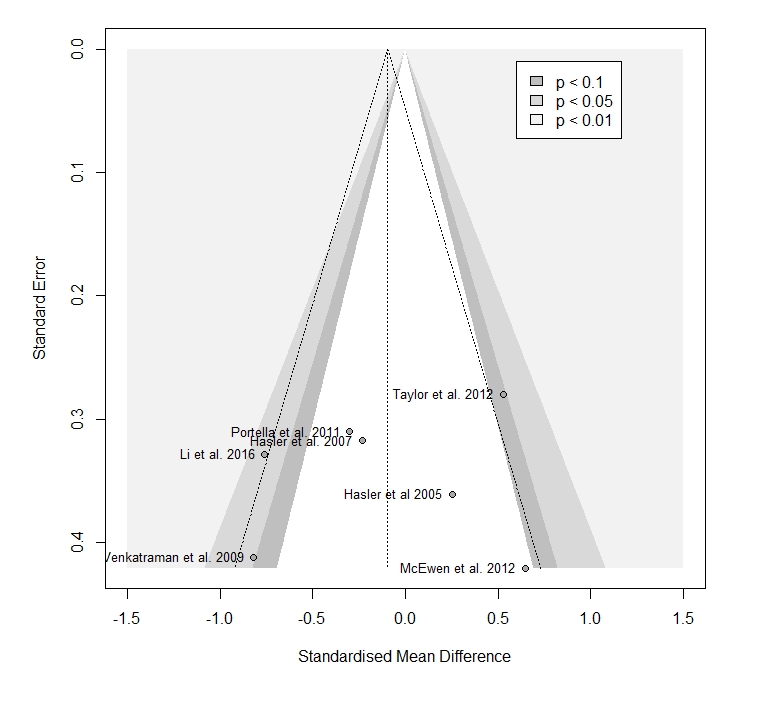

#### 5. Parietal lobe

##### Main results

We included 2 studies: 52 patients and 52 controls.

Supplementary Figure 18. Forest plot of all studies examining NAA levels in the parietal lobe of patients with cMDD compared to healthy controls.

The meta-analysis revealed no significant difference between patients and controls (n = 2, Hedges’ g = -1.264, 95% CI -12.481 to 9.953; p = 0.388; Q = 15.37, *I^2^=* 93.5%, p < 0.0001). Positive values favour cMDD, while negative values favour controls.

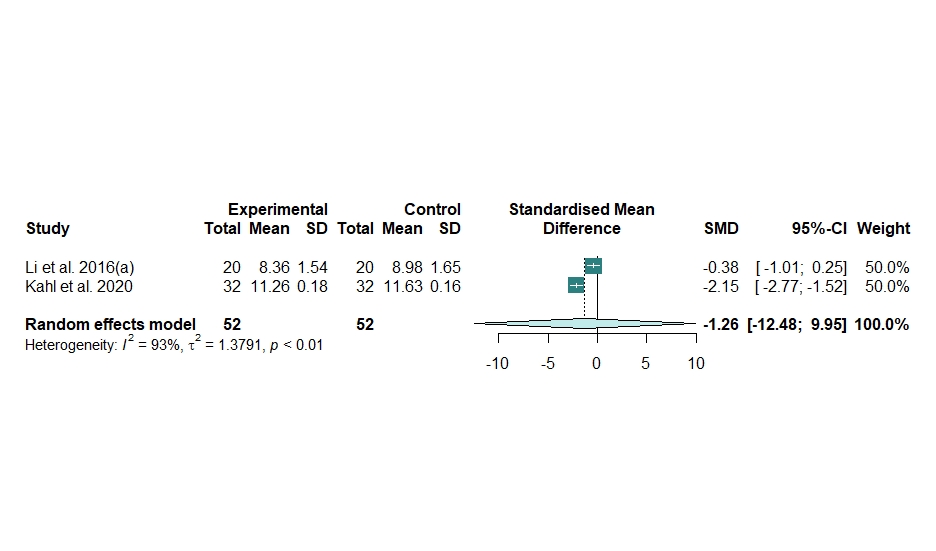

#### 6. Temporal lobe

##### Main results

We included 3 studies: 66 patients and 60 controls.

Supplementary Figure 19. Forest plot of all studies examining NAA levels in the temporal lobe of patients with cMDD compared to healthy controls.

The meta-analysis revealed no difference between patients and controls (n = 3, Hedges’ g = -0.217, 95% CI -3.042 to 2.608; p = 0.772; Q = 23.63, *I^2^=* 91.5%, p < 0.0001). Positive values favour cMDD, while negative values favour controls.

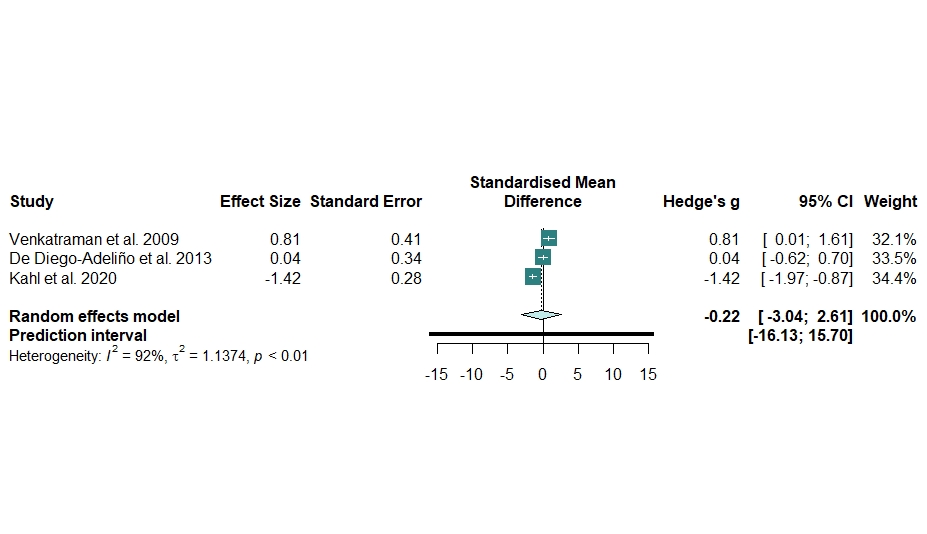

##### Heterogeneity

No outlier was identified.

The between-study heterogeneity was significant and substantial (Q = 23.63, *I^2^=* 91.5%, p < 0.0001).

##### Publication bias

Supplementary Figure 20. Assessment of small sample publication bias with the contour-enhanced funnel plot for the studies examining NAA levels in the temporal lobe of cMDD.

No evidence of publication bias emerged from the funnel plot. Egger’s test was not performed because there were not enough studies.

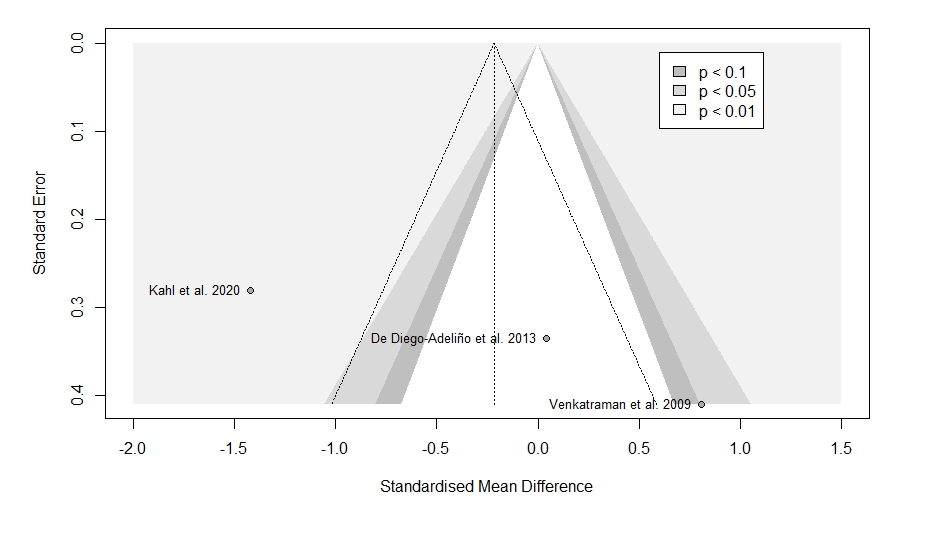

#### 7. Occipital lobe

##### Main results

We included 4 studies: 87 patients and 89 controls.

Supplementary Figure 21. Forest plot of all studies examining NAA levels in the occipital lobe of patients with cMDD compared to healthy controls.

The meta-analysis revealed a significant difference between patients and controls (n = 4, Hedges’ g = -0.677, 95% CI -1.013 to -0.341; p = 0.007; Q = 1.39, *I^2^=* 0%, p < 0.708). Positive values favour cMDD, while negative values favour controls.

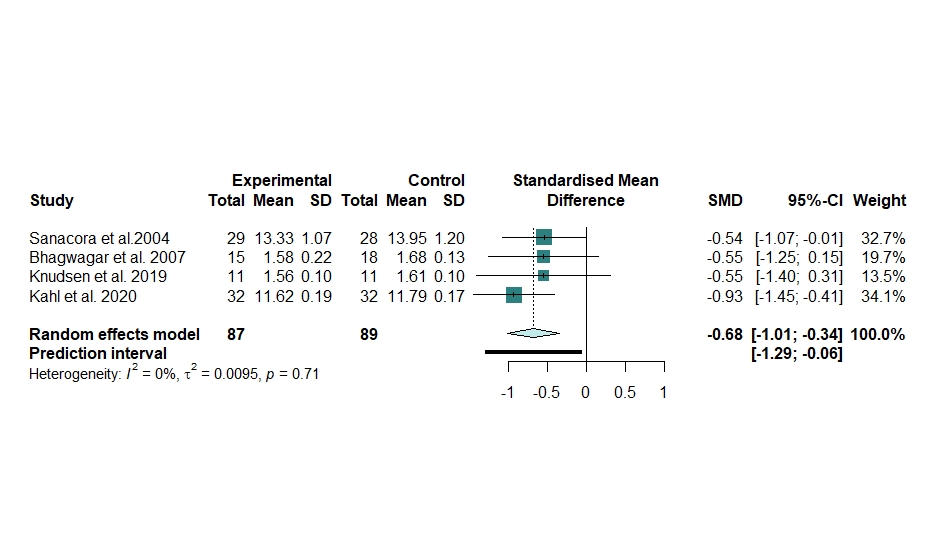

##### Heterogeneity

No outliers were detected.

The between-study heterogeneity was not significant (Q = 1.39, *I^2^=* 0%, p < 0.708).

Supplementary Figure 22. Heterogeneity assessment in the occipital lobe meta-analysis

**A**. Baujat plot of the contribution of each study to the overall heterogeneity (as measured by Cochran’s Q) and its influence on the pooled effect size. **B.** Influence analysis with the *leave-one-out* method of the meta-analysis of all studies of cMDD examining NAA levels in the occipital lobe. Parameters of the influence analysis: standardized residuals, dffits, Cook’s distance, covariance ratio, tau^2^, Q, hat, and weight. One study ^5^ was identified as influential according to the cut-offs proposed by Viechtbauer and Cheung, 2010 ^8^. **C.** Forest plot of the overall effect sizes of the meta-analyses of all studies of cMDD examining NAA levels in the occipital lobe, recalculated with the *leave-one-out* method, ordered by effect size. **D**. Forest plot of the overall effect sizes of the meta-analyses of all studies of cMDD examining NAA levels in the occipital lobe, recalculated with the *leave-one-out* method, ordered by heterogeneity.

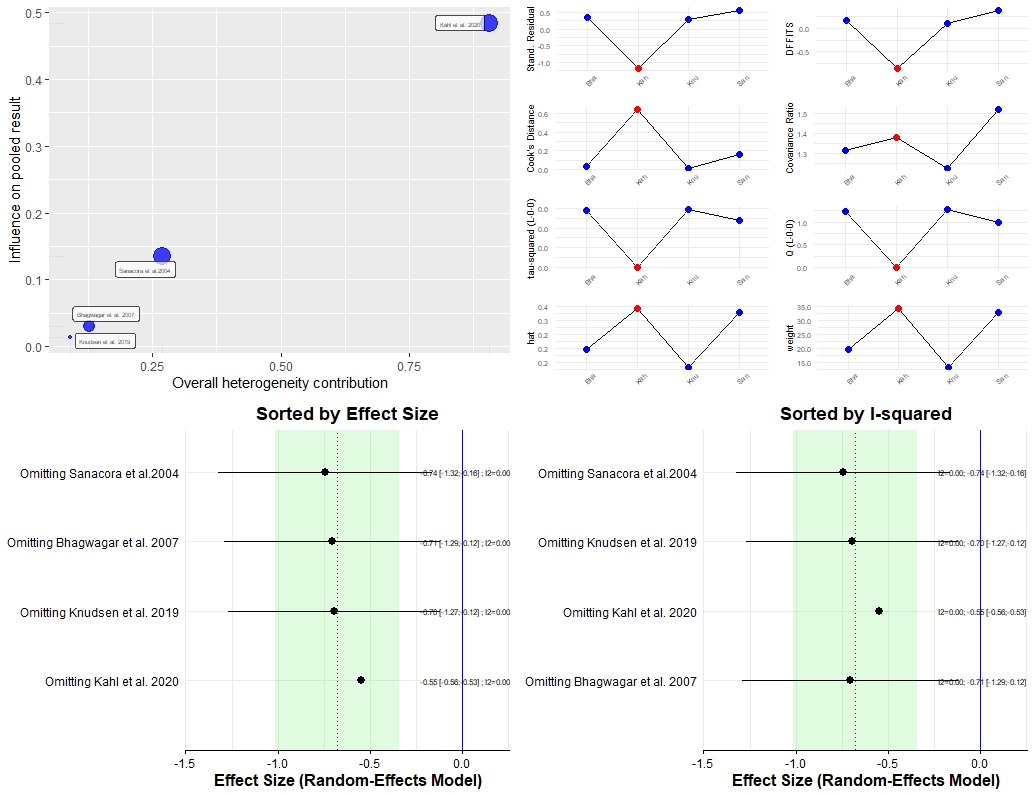

##### Publication bias

Supplementary Figure 23. Assessment of small sample publication bias with the contour-enhanced funnel plot for the studies examining NAA levels in the occipital lobe of cMDD.

No evidence of publication bias seemed to emerge from the funnel plot. Egger’s test was not performed because there were not enough studies.

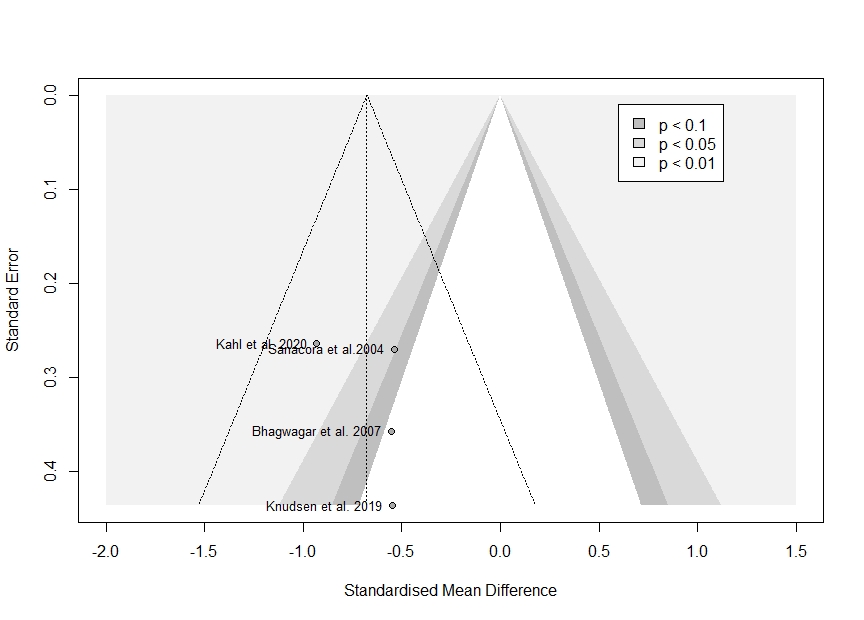

#### 8. Anterior cingulate cortex

##### Main results

We included 17 studies: 388 patients and 314 controls.

Supplementary Figure 24. Forest plot of all studies examining NAA levels in the anterior cingulate cortex of patients with cMDD compared to healthy controls.

The meta-analysis revealed no significant difference between patients and controls (n = 17, Hedges’ g = -0.141, 95% CI -0.4174; 0.1358; p = 0.297; Q = 41.45, *I^2^=* 61.4%, p = 0.0005). Positive values favour cMDD, while negative values favour controls.

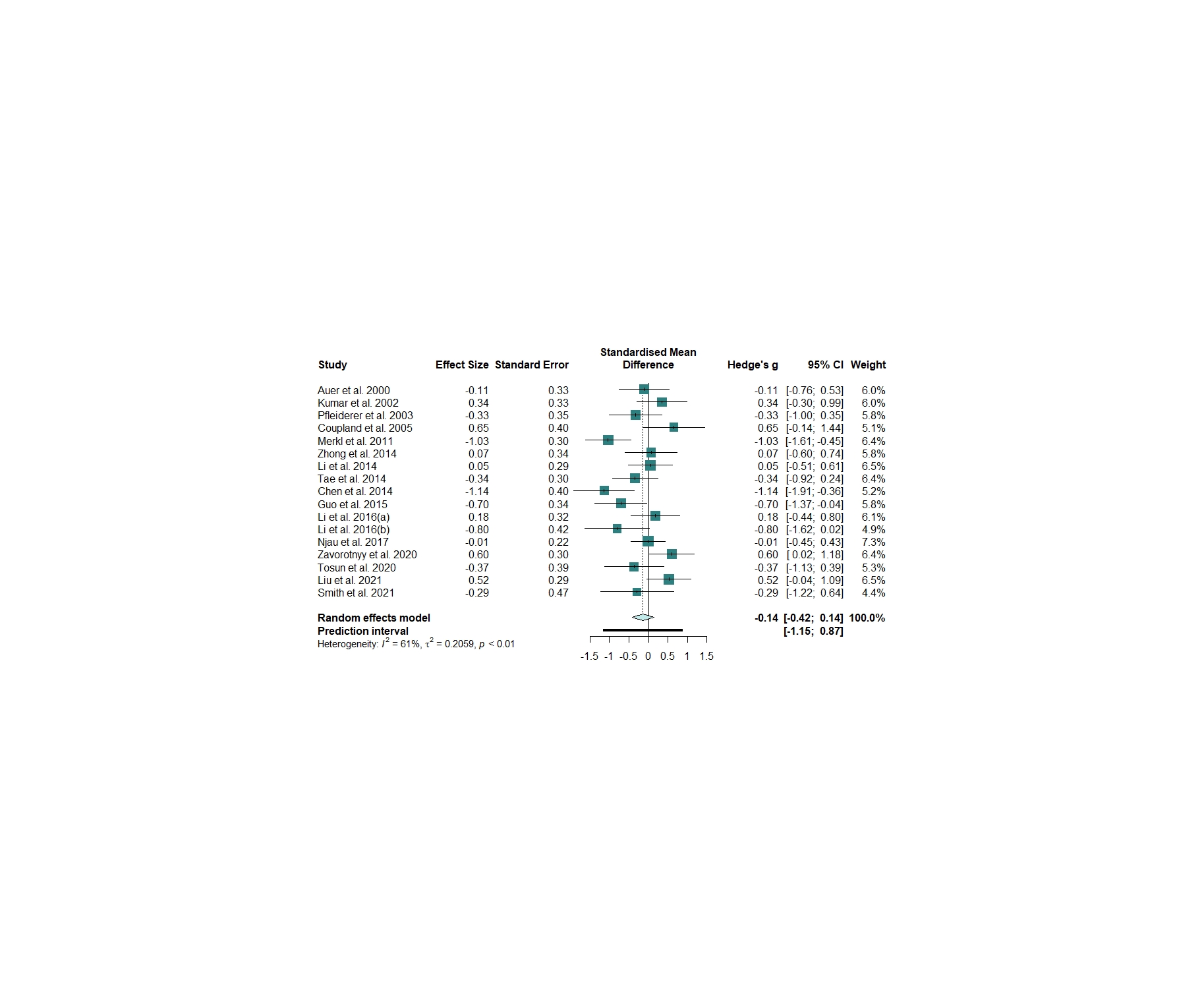

##### Heterogeneity

We identified and removed one outlier ^10^.

The updated meta-analysis confirmed no significant difference between patients and controls (n = 16, Hedges’ g = --0.077; p = 0.5444), with significant moderate heterogeneity (Q = 31.07, *I^2^=* 51.7%, p = 0.008).

Supplementary Figure 25. Heterogeneity assessment in the anterior cingulate cortex meta-analysis

**A.** Baujat plot of the contribution of each study to the overall heterogeneity (as measured by Cochran’s Q) and its influence on the pooled effect size. **B.** Influence analysis with the *leave-one-out* method of the meta-analysis of all studies of cMDD examining NAA levels in the anterior cingulate cortex. Parameters of the influence analysis: standardized residuals, dffits, Cook’s distance, covariance ratio, tau^2^, Q, hat, and weight. No studies were identified as influential according to the cut-offs proposed by Viechtbauer and Cheung ^8^. **C.** Forest plot of the overall effect sizes of the meta-analyses of all studies of cMDD examining NAA levels in the anterior cingulate cortex, recalculated with the *leave-one-out* method, ordered by effect size. **D.** Forest plot of the overall effect sizes of the meta-analyses of all studies of cMDD examining NAA levels in the anterior cingulate cortex, recalculated with the *leave-one-out* method, ordered by heterogeneity.

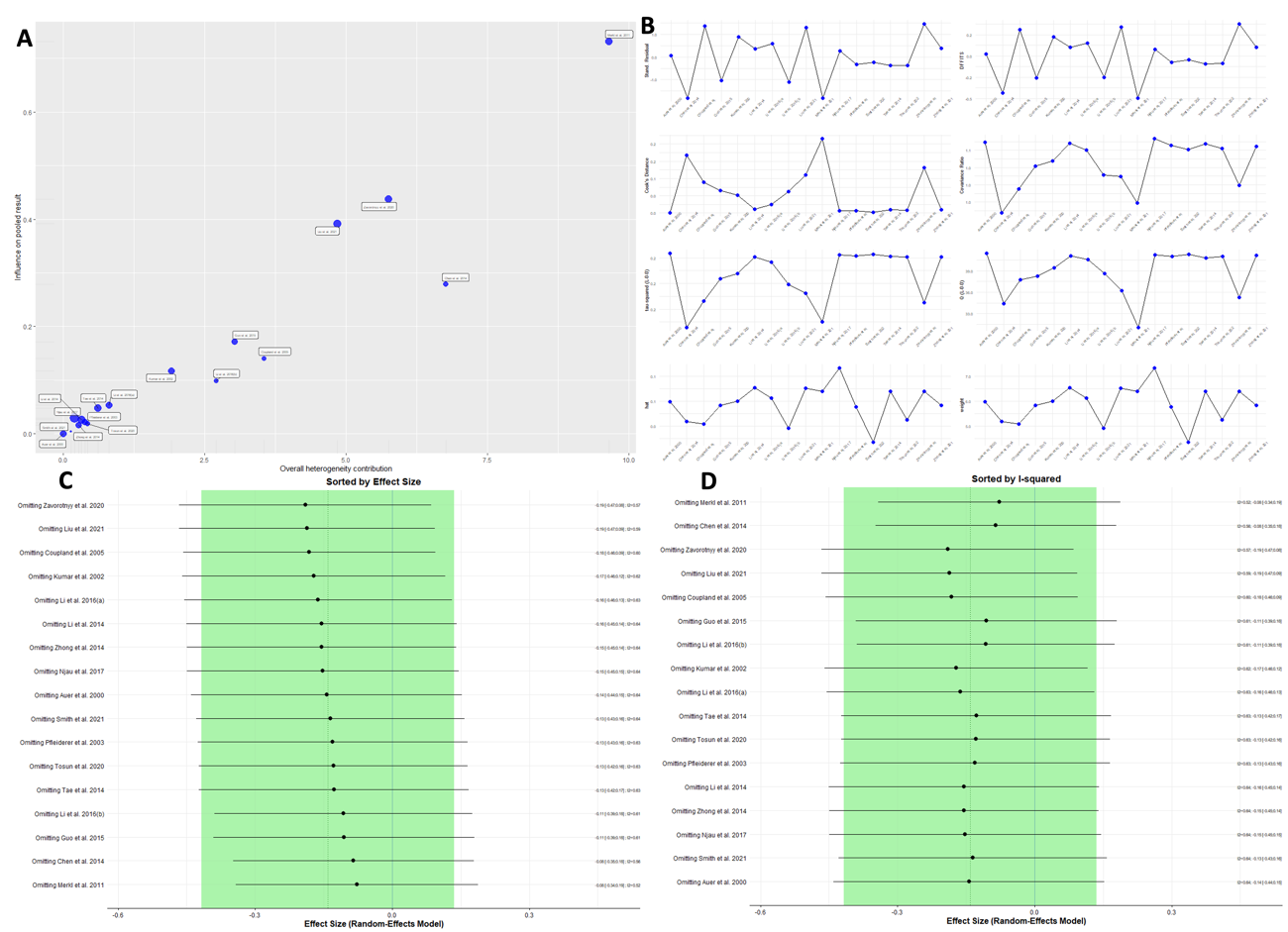

Supplementary Figure 26. GOSH diagnostics in the anterior cingulate cortex meta-analysis

**A**. GOSH plot showing the meta-analysis models of all studies of cMDD examining NAA levels in the anterior cingulate cortex, fitted to all 2^k-1^ possible combinations of the included studies (x-axis, pooled effect anterior cingulate cortex; y-axis, between-study heterogeneity). The GOSH plot did not highlight distinct clusters of heterogeneity. **B.** DBSCAN Algorithm. **C.** Gaussian mixture model. **D.** GOSH plot fitted to all 2^k-1^ possible combinations of the included studies with and without the outlier with most extreme effect size (Merkl et al., 2011) ^10^.

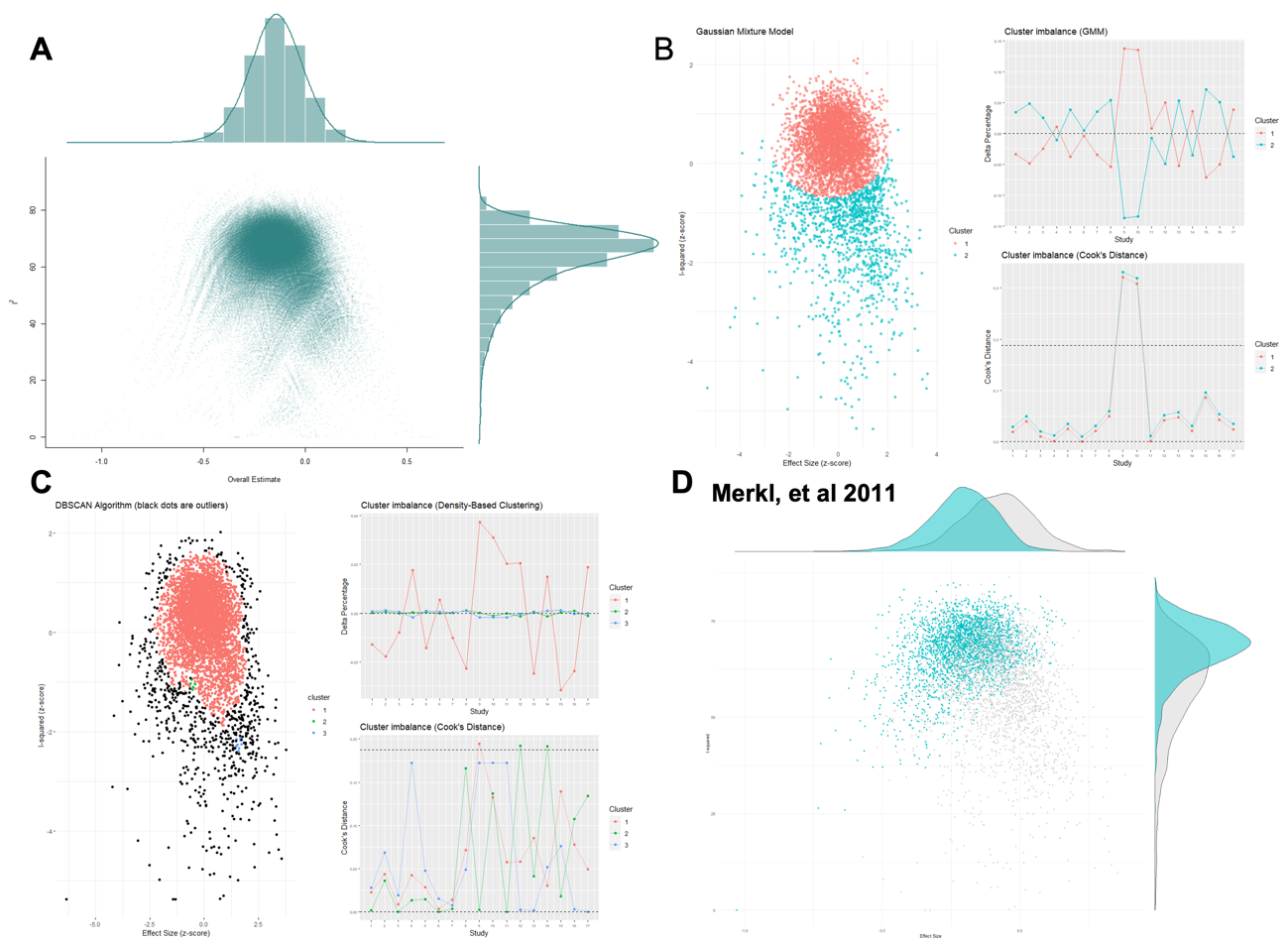

##### Subgroup Analyses

Supplementary Table 8. Subgroup analysis of the meta-analyses of all studies examining NAA levels in the anterior cingulate cortex of patients with cMDD compared to healthy controls.

|  | Subgroup sample size | Hedges’ g | 95% CI | p | I² | 95% CI | p subgroups |
| --- | --- | --- | --- | --- | --- | --- | --- |
| NAA quantification method | | | | | | | 0.110 |
| Cr scaling | 8 | 0.019 | -0.345, 0.382 | 0.920 | 57% | 6, 81% |  |
| Absolute | 8 | -0.368 | -0.688, -0.05 | 0.023 | 53% | 0, 79% |  |
| CSF correction | | | | | | | 0.261 |
| Yes | 8 | -0.290 | -0.722, 0.14 | 0.186 | 70% | 38, 86% |  |
| No | 9 | 0.007 | -0.283, 0.297 | 0.962 | 42% | 0, 73% |  |
| Field strength | | | | | | | 0.435 |
| 1.5 T | 6 | -0.229 | -0.618, 0.159 | 0.247 | 47% | 0, 79% |  |
| > 1.5 T | 11 | -0.093 | -0.436, 0.250 | 0.596 | 68% | 40, 83% |  |

##### Metaregressions

Supplementary Table 9. Results of the meta-regressions in the anterior cingulate cortex.

|  | beta | SE | p |
| --- | --- | --- | --- |
| Age | -0.0040 | 0.0092 | 0.668 |
| Female % | -0.0149 | 0.0086 | 0.1046 |
| Illness duration | 0.0076 | 0.0422 | 0.866 |
| Ham-D scale | -0.0008 | 0.0263 | 0.976 |
| ^1^H-MRS field strength | -0.0587 | 0.0881 | 0.515 |
| TE | 0.0027 | 0.0028 | 0.35 |
| TR | -0.0003 | 0.0002 | 0.125 |

##### Publication bias

Supplementary Figure 27. Assessment of small sample publication bias with the contour-enhanced funnel plot for the studies examining NAA levels in the anterior cingulate cortex of cMDD.

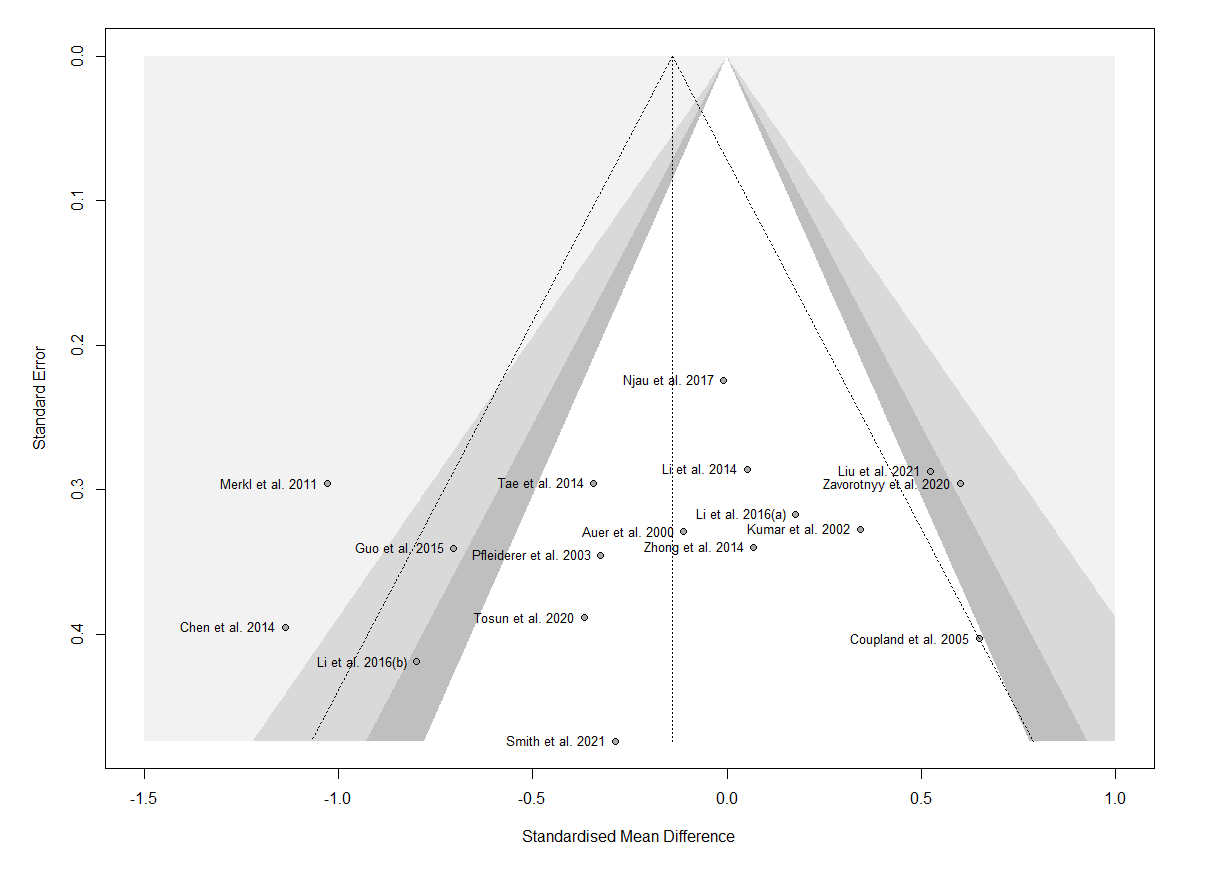

Supplementary Figure 28. P-curve for the analysis of cMDD in the anterior cingulate cortex.

The analysis of the p-curve confirmed the presence of evidential value: the right-skewness test was significant for the half p-curve (p<0.05), while the flatness test was not significant for both the full and half curve. Overall, the p-curve appears right-skewed, with highly significant results (p-values = 0.01) representing 50% of all p-values. This can be interpreted as evidence of no p-hacking nor selective reporting.

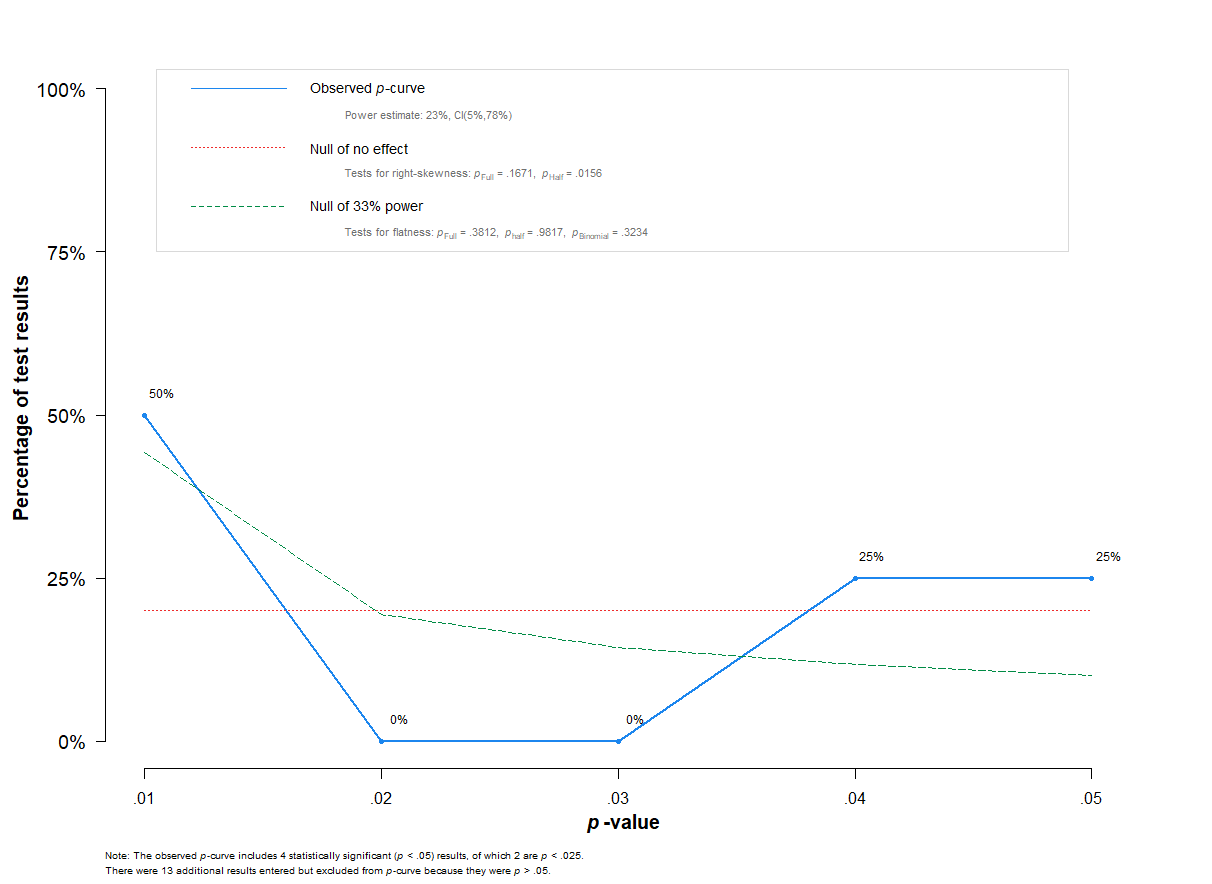

##### Sensitivity analysis on the influence of geriatric populations

We proceeded to make a sensitivity analysis omitting three studies analyzing geriatric subjects ^11-13^. No difference was found between patients and controls (n = 15, Hedges’ g = -0.140, 95% CI -0.435 to 0.155; p = 0.326). Between-study heterogeneity remained significant and moderate (Q = 36.76, *I²* = 61.9%, p < 0.001).

#### 9. Posterior cingulate cortex

##### Main results

We included 3 studies: 47 patients and 38 controls.

Supplementary Figure 29. Forest plot of all studies examining NAA levels in the posterior cingulate cortex of patients with cMDD compared to healthy controls.

The meta-analysis revealed no significant difference between patients and controls (n = 3, Hedges’ g = -0.503, 95% CI -2.321 to 1.262; p = 0.331; Q = 6.19, *I^2^=* 67.7%, p = 0.045). Positive values favour cMDD, while negative values favour controls.

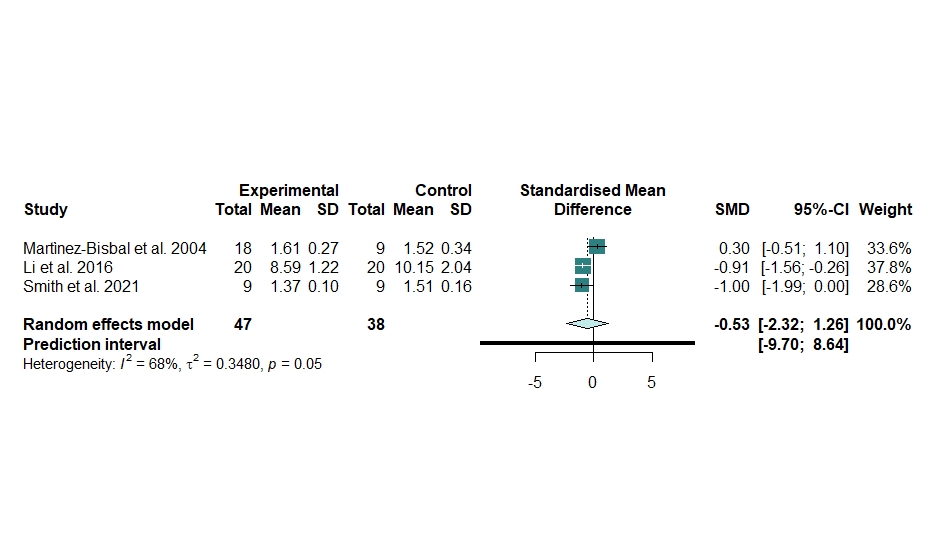

##### Heterogeneity

No outlier was identified.

The between-study heterogeneity was significant and moderate (Q = 6.19, *I^2^=* 67.7%, p = 0.045), but no other analyses were performed because of the shortage of the studies (n = 3).

##### Publication bias

Supplementary Figure 30. Assessment of small sample publication bias with the contour-enhanced funnel plot for the studies examining NAA levels in the posterior cingulate cortex of cMDD.

We found no evidence of publication bias. Egger’s test was not performed because there were not enough studies.

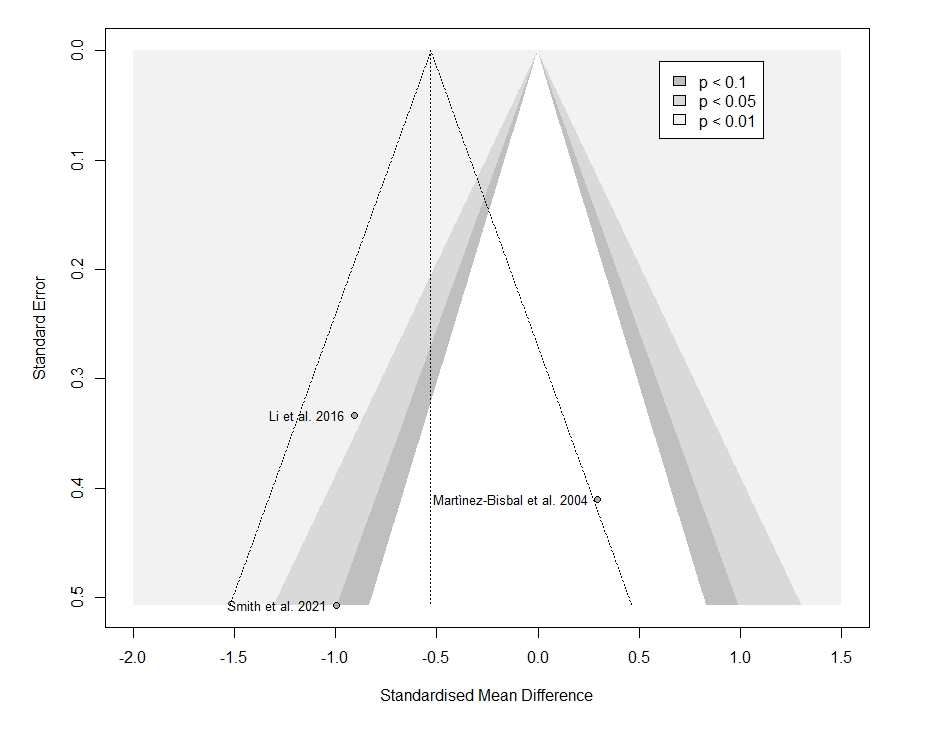

#### 10. Hippocampus

##### Main results

We included 9 studies: 211 patients and 157 controls.

Supplementary Figure 31. Forest plot of all studies examining NAA levels in the hippocampus of patients with cMDD compared to healthy controls.

The meta-analysis revealed no significant difference between patients and controls (n = 9, Hedges’ g = -0.093, 95% CI -1.628 to 1.442; p = 0.892; Q = 150.85, *I^2^=* 94.7%, p < 0.0001). Positive values favour cMDD, while negative values favour controls.

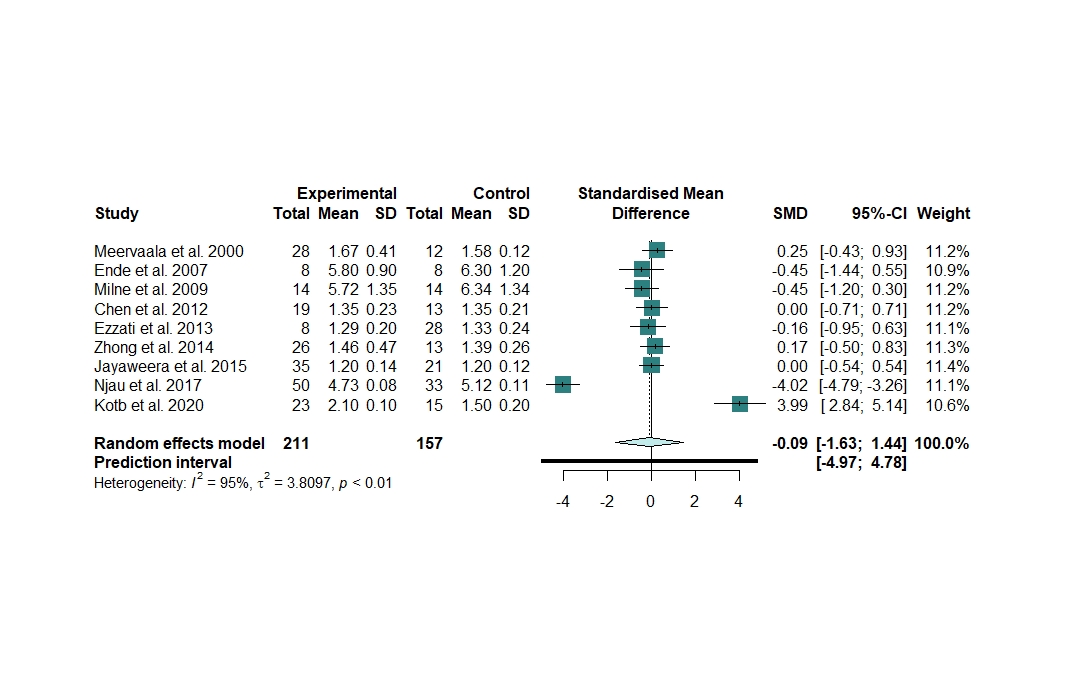

##### Heterogeneity

We identified and removed 2 outliers ^14, 15^.

The updated meta-analysis did not find a significant difference between patients and controls (n = 7, Hedges’ g = -0.047, 95% CI -0.283 to 0.190; p = 0.647). However, the between-study heterogeneity became null (Q = 2.96, *I²* = 0%, p = 0.814), meaning that these two outliers explained the whole heterogeneity.

Supplementary Figure 32. Forest plot of studies examining NAA levels in the hippocampus of patients with cMDD compared to healthy controls after removing the outliers.

Positive values favour MDD, while negative values favour controls.

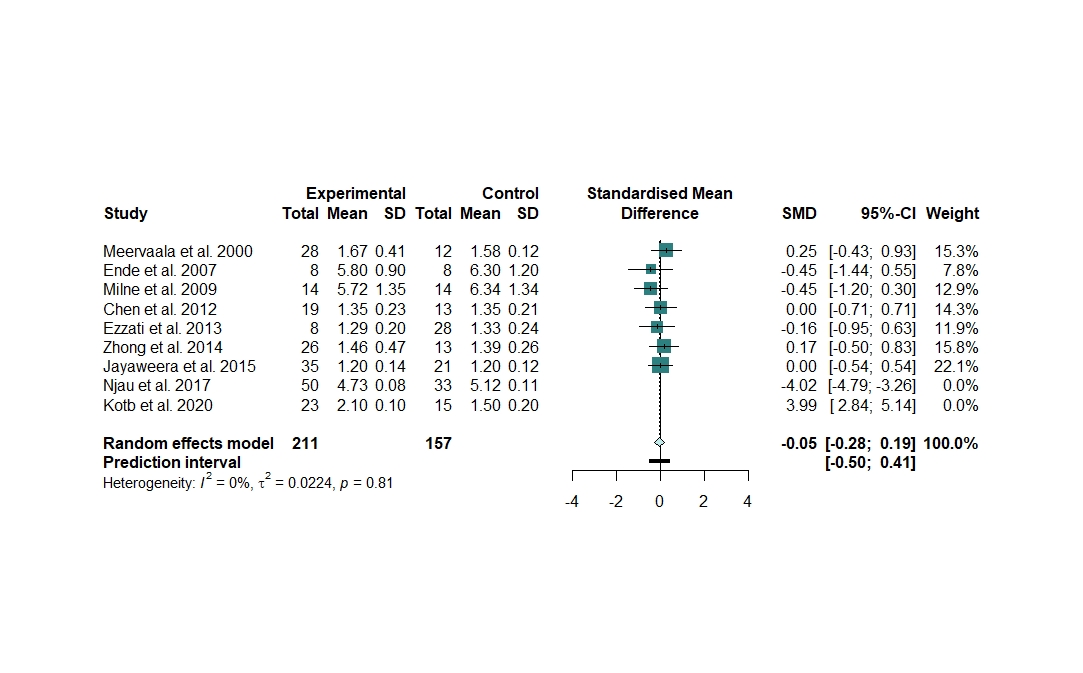

Supplementary Figure 33. Heterogeneity assessment in the hippocampus meta-analysis

**A.** Baujat plot of the contribution of each study to the overall heterogeneity (as measured by Cochran’s Q) and its influence on the pooled effect size. **B**. Influence analysis with the *leave-one-out* method of the meta-analysis of all studies of chronic MDD examining NAA levels in the hippocampus. Parameters of the influence analysis: standardized residuals, dffits, Cook’s distance, covariance ratio, tau^2^, Q, hat, and weight. Two studies ^14, 15^ were identified as influential according to the cut-offs proposed by Viechtbauer and Cheung, 2010 ^8^ . **C**. Forest plot of the overall effect sizes of the meta-analyses of all studies of cMDD examining NAA levels in the hippocampus, recalculated with the *leave-one-out* method, ordered by effect size. **D**. Forest plot of the overall effect sizes of the meta-analyses of all studies of cMDD examining NAA levels in the hippocampus, recalculated with the *leave-one-out* method, ordered by heterogeneity.

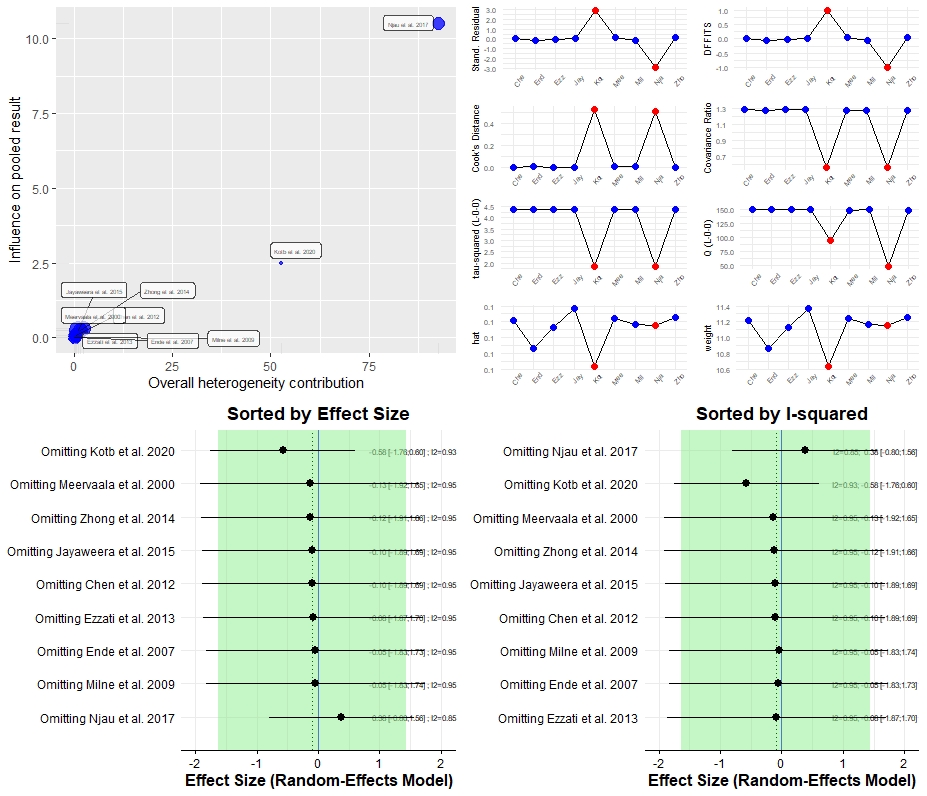

Supplementary Figure 34. GOSH diagnostics in the hippocampus meta-analysis

**A**. GOSH plot showing the meta-analysis models of all studies of cMDD examining NAA levels in the hippocampus, fitted to all 2^k-1^ possible combinations of the included studies (x-axis, pooled effect size; y-axis, between-study heterogeneity). The GOSH plot highlighted 4 clusters: 3 with high heterogeneity and with negative, null and positive effect size, respectively; the 4^th^ one with low heterogeneity and effect size around 0. GOSH diagnostics confirmed the presence of two studies with a large influence on the heterogeneity, corresponding to the two identified outliers ^14, 15^. **B.** DBSCAN Algorithm. **C**. GOSH plot fitted to all 2^k-1^ possible combinations of the included studies with and without Njau et al., 2017 ^15^. **D**. GOSH plot fitted to all 2^k-1^ possible combinations of the included studies with and without Kotb et al., 2020 ^14^.

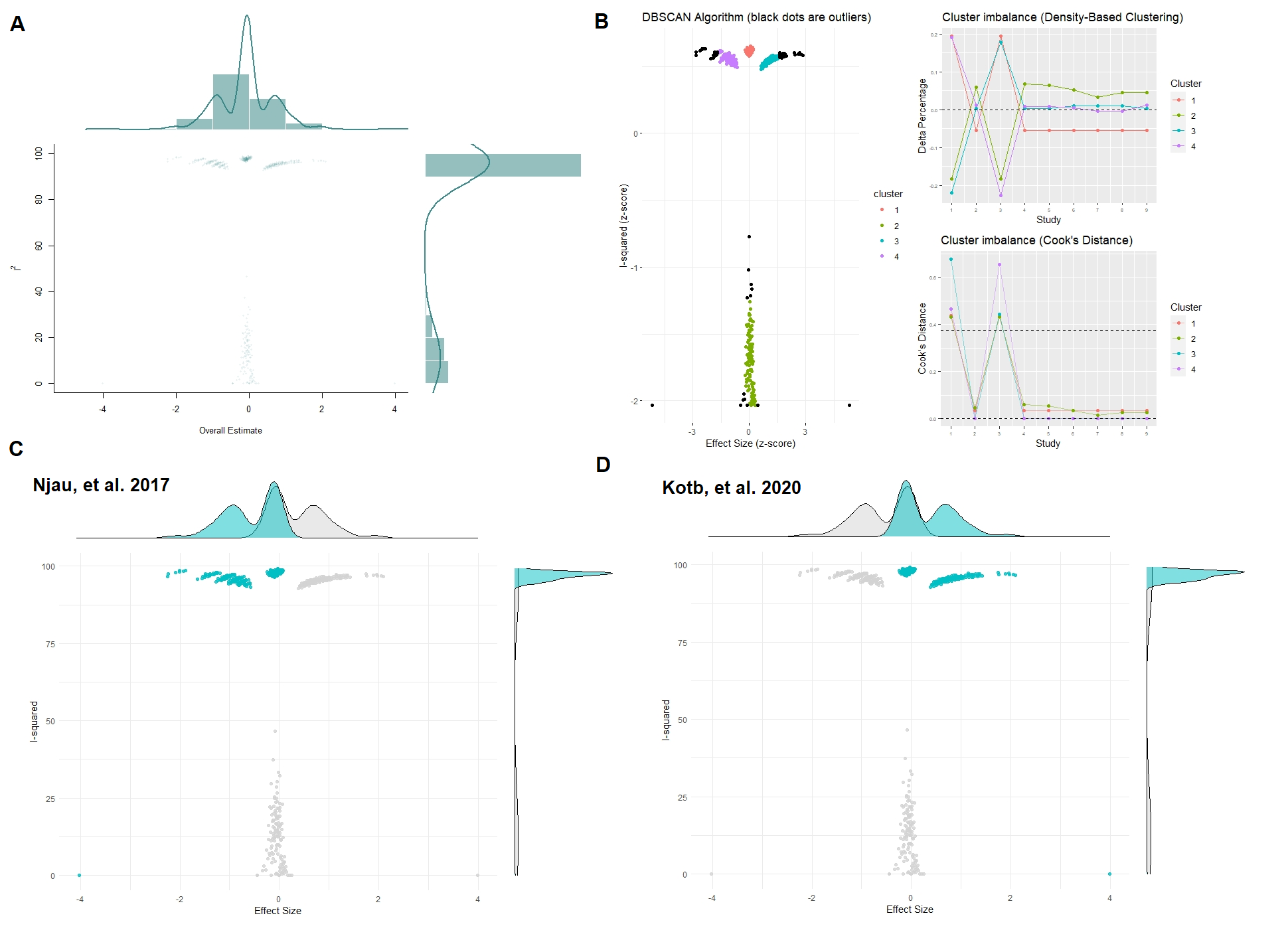

##### Subgroup Analyses

Supplementary Table 10. Subgroup analysis of the meta-analyses of all studies examining NAA levels in the hippocampus of patients with cMDD compared to healthy controls.

|  | Subgroup sample size | Hedges’ g | 95% CI | p | I² | 95% CI | p subgroups |
| --- | --- | --- | --- | --- | --- | --- | --- |
| Field strength | | | | | | | 0.125 |
| 1.5 T | 5 | 0.759 | -0.796, 2.314 | 0.339 | 91% | 81, 95% |  |
| > 1.5 T | 4 | -1.152 | -3.029, 0.726 | 0.229 | 96% | 93, 98% |  |

##### Publication bias

Supplementary Figure 35. Assessment of small sample publication bias with the contour-enhanced funnel plot for the studies examining NAA levels in the hippocampus of cMDD.

We found no evidence of publication bias. The two outliers are extremely out of the funnel: Njau et al 2017 toward very low effect size and low standard error, Kotb et al., 2020 towards very high effect size and standard error.

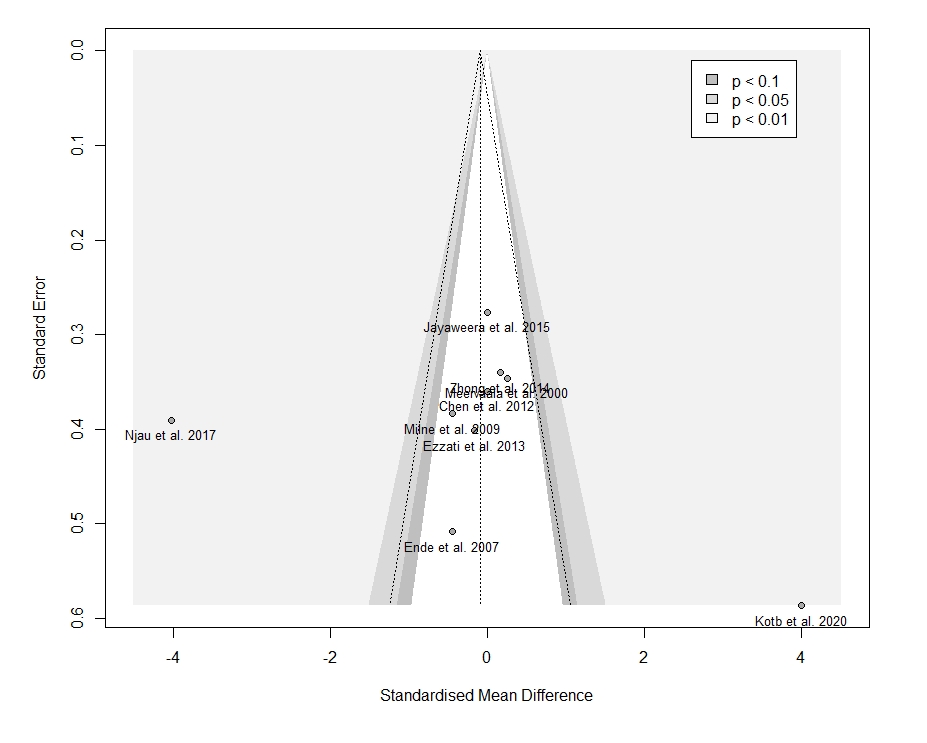

##### Sensitivity analysis on the influence of geriatric populations

We proceeded to make a sensitivity analysis omitting two studies analyzing geriatric subjects ^16, 17^. The meta-analysis revealed still no difference between patients and controls (n = 7, Hedges’ g = -0.091, 95% CI -2.232 to 2.049; p = 0.920), with a significant between-study heterogeneity (Q = 149.49, *I²* = 96%, p < 0.001).

#### 11. Thalamus

##### Main results

We included 5 studies: 91 patients and 71 controls.

Supplementary Figure 36. Forest plot of all studies examining NAA levels in the thalamus of patients with cMDD compared to healthy controls.

The meta-analysis revealed no significant difference between patients and controls (n = 5, Hedges’ g = -0.4234, 95% CI -1.085 to 0.239; p = 0.15; Q = 8.95, *I^2^=* 55.3%, p = 0.062). Positive values favour cMDD, while negative values favour controls.

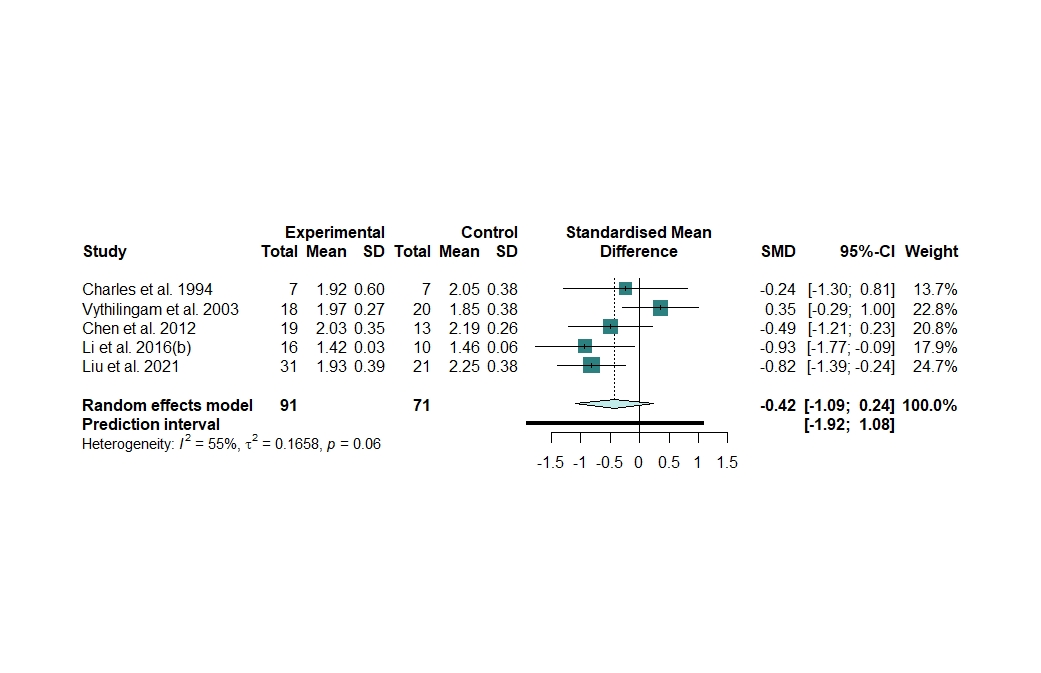

##### Heterogeneity

We identified no outliers^4-7^.

The between-study heterogeneity had a trend toward significance (Q = 8.95, *I^2^=* 55.3%, p = 0.062).

Supplementary Figure 37. Heterogeneity assessment in the thalamus meta-analysis

**A**. Baujat plot of the contribution of each study to the overall heterogeneity (as measured by Cochran’s Q) and its influence on the pooled effect size. **B.** Influence analysis with the *leave-one-out* method of the meta-analysis of all studies of cMDD examining NAA levels in the thalamus. Parameters of the influence analysis: standardized residuals, dffits, Cook’s distance, covariance ratio, tau^2^, Q, hat, and weight. One study ^18^ was identified as influential according to the cut-offs proposed by Viechtbauer and Cheung, 2010. **C.** Forest plot of the overall effect sizes of the meta-analyses of all studies of cMDD examining NAA levels in the thalamus, recalculated with the *leave-one-out* method, ordered by effect size. **D**. Forest plot of the overall effect sizes of the meta-analyses of all studies of cMDD examining NAA levels in the thalamus, recalculated with the *leave-one-out* method, ordered by heterogeneity.

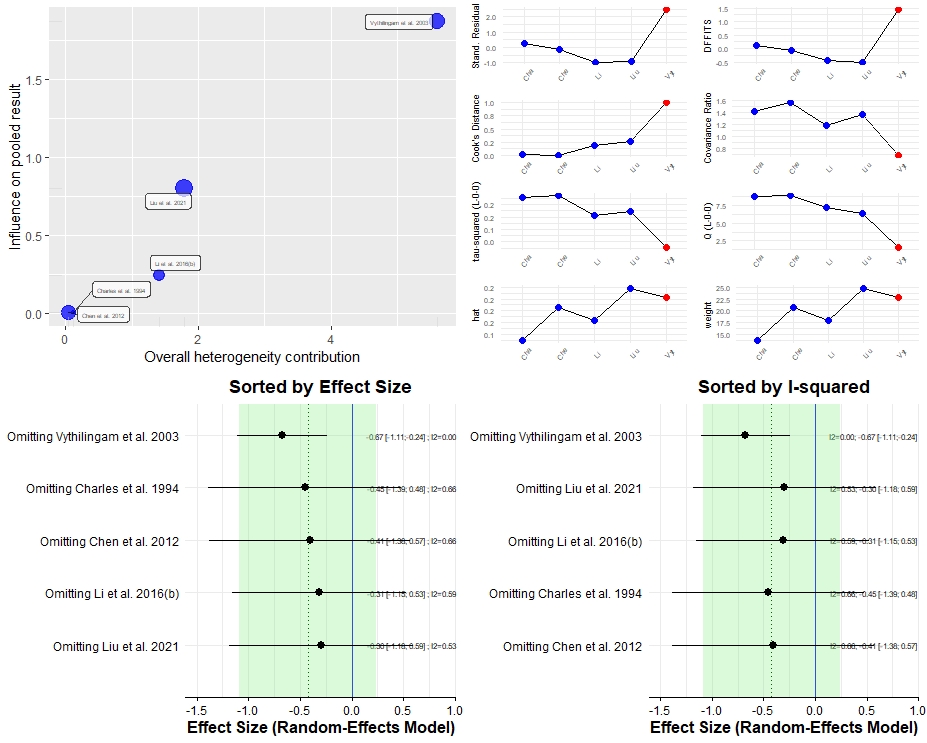

Supplementary Figure 38. Forest plot of studies of cMDD examining NAA levels in the thalamus after removing one influential study.

The updated meta-analysis found significant results: NAA levels in the thalamus of cMDD patients were significantly lower than controls (n = 4, Hedges’ g = -0.673, 95% CI -1.108 to -0.238; p = 0.016; Q = 1.49, *I²* = 0%, p = 0.686). Positive values favour MDD, while negative values favour controls.

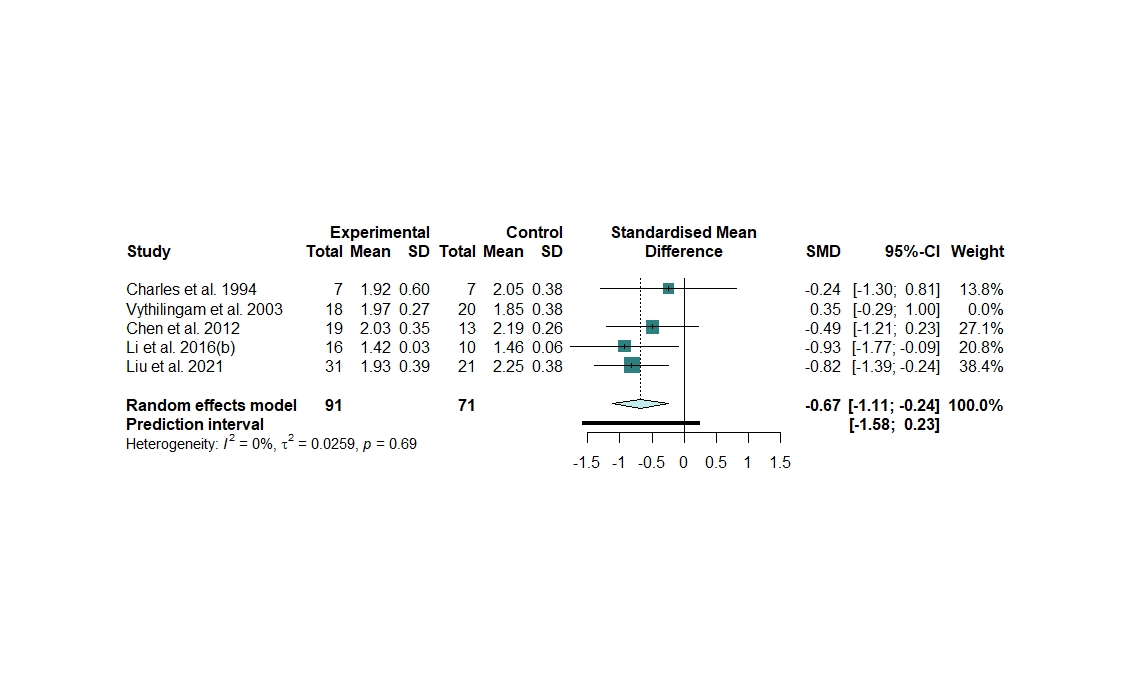

##### Publication bias

Supplementary Figure 39. Assessment of small sample publication bias with the contour-enhanced funnel plot for the studies examining NAA levels in the frontal lobe of cMDD.

We found no evidence of publication bias. Egger’s test was not performed because there were not enough studies.

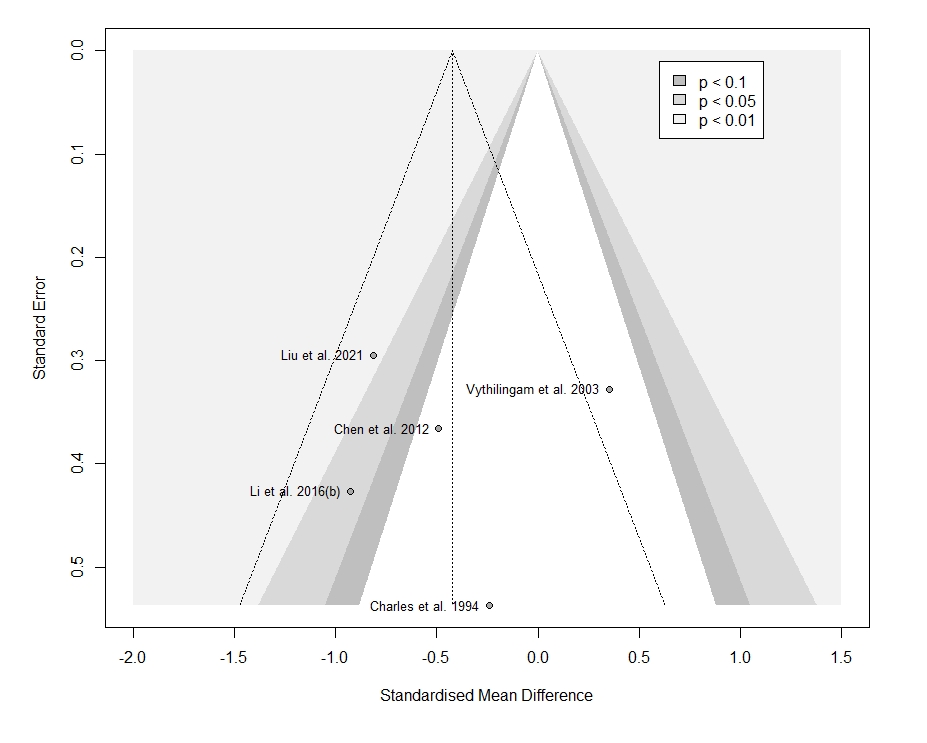

#### 12. Basal ganglia

##### Main results

We included 10 studies: 252 patients and 219 controls.

Supplementary Figure 40. Forest plot of all studies examining NAA levels in the basal ganglia of patients with cMDD compared to healthy controls.

The meta-analysis revealed no significant difference between patients and controls (n = 10, Hedges’ g = -0.1684, 95% CI -0.495 to 0.159; p = 0.274; Q = 18.52, *I^2^=* 51.4%, p = 0.03). Positive values favour cMDD, while negative values favour controls.

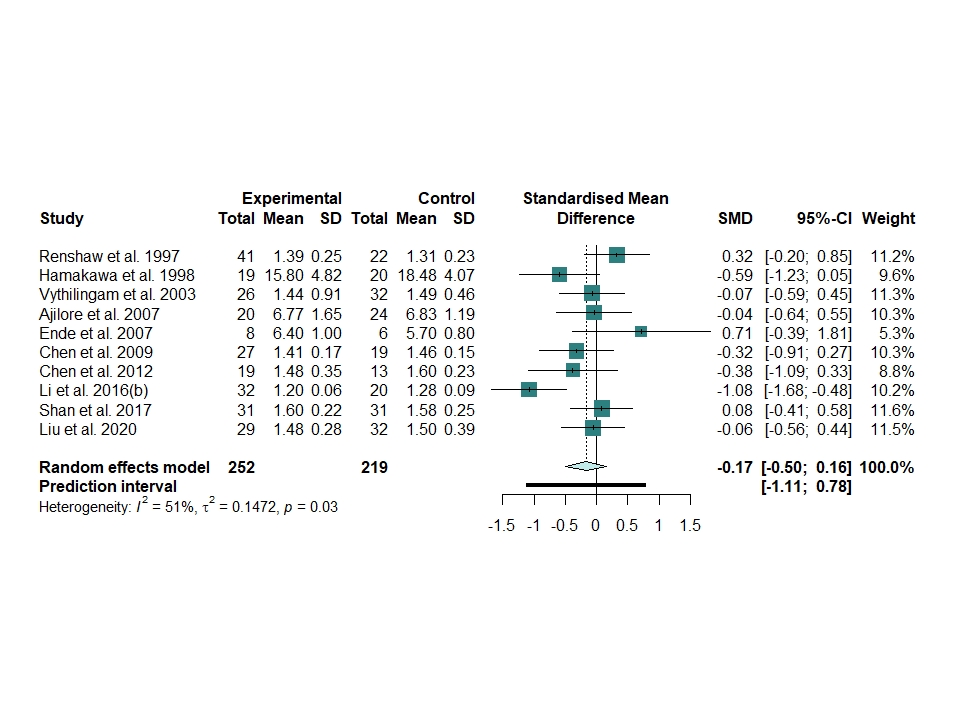

##### Heterogeneity

The between-study heterogeneity was significant and moderate (Q = 18.52, *I^2^=* 51.4%, p = 0.03), but no outliers were detected.

Supplementary Figure 41. Heterogeneity assessment in the basal ganglia meta-analysis

**A**. Baujat plot of the contribution of each study to the overall heterogeneity (as measured by Cochran’s Q) and its influence on the pooled effect size. **B**. Influence analysis with the *leave-one-out* method of the meta-analysis of all studies of cMDD examining NAA levels in the basal ganglia. Parameters of the influence analysis: standardized residuals, dffits, Cook’s distance, covariance ratio, tau^2^, Q, hat, and weight. No studies were identified as influential according to the cut-offs proposed by Viechtbauer and Cheung, 2010 ^8^ **C**. Forest plot of the overall effect sizes of the meta-analyses of all studies of cMDD examining NAA levels in the basal ganglia, recalculated with the *leave-one-out* method, ordered by effect size. **D**. Forest plot of the overall effect sizes of the meta-analyses of all studies of cMDD examining NAA levels in the basal ganglia, recalculated with the *leave-one-out* method, ordered by heterogeneity.

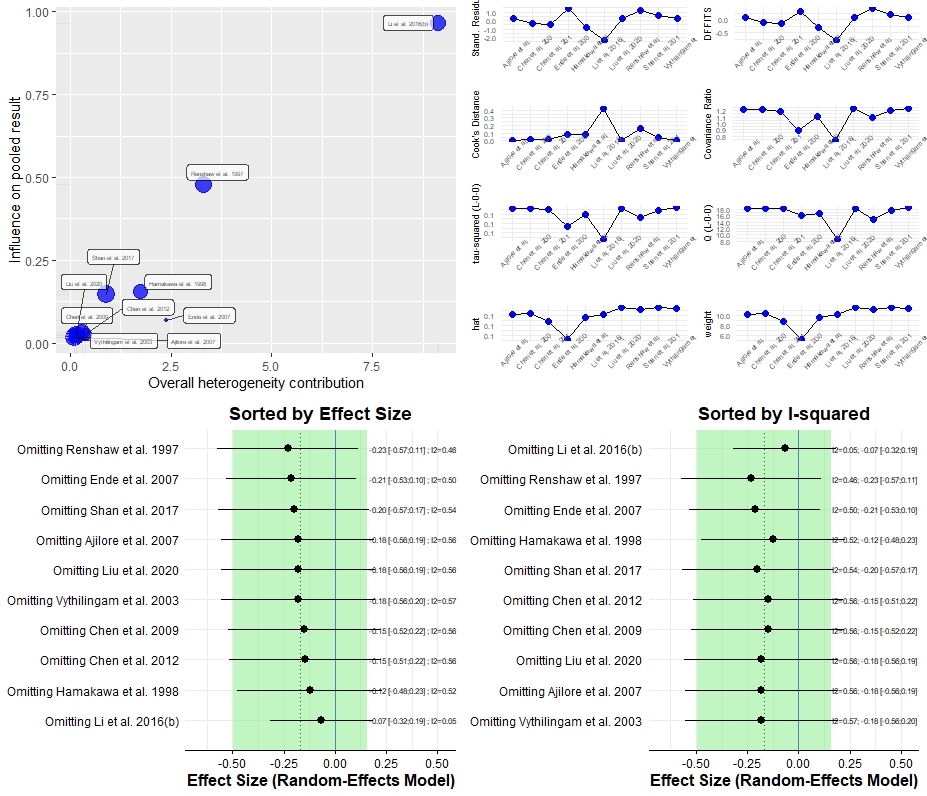

Supplementary Figure 42. GOSH diagnostics in the basal ganglia meta-analysis

**A**. GOSH plot showing the meta-analysis models of all studies of cMDD examining NAA levels in the basal ganglia, fitted to all 2^k-1^ possible combinations of the included studies (x-axis, pooled effect size; y-axis, between-study heterogeneity). The GOSH plot highlighted 3 clusters: one with high heterogeneity, one with moderate heterogeneity and the other with low heterogeneity. The cluster with high heterogeneity was also the one with the largest negative effect size, although there were no substantial effect size variations across clusters. **B**. K-means algorithm. **C**. Gaussian Mixture Model. The GOSH diagnostic function confirmed the presence of one study ^19^ with a quite large influence on the heterogeneity. **D.** GOSH plot fitted to all 2^k-1^ possible combinations of the included studies with and without this study ^19^.

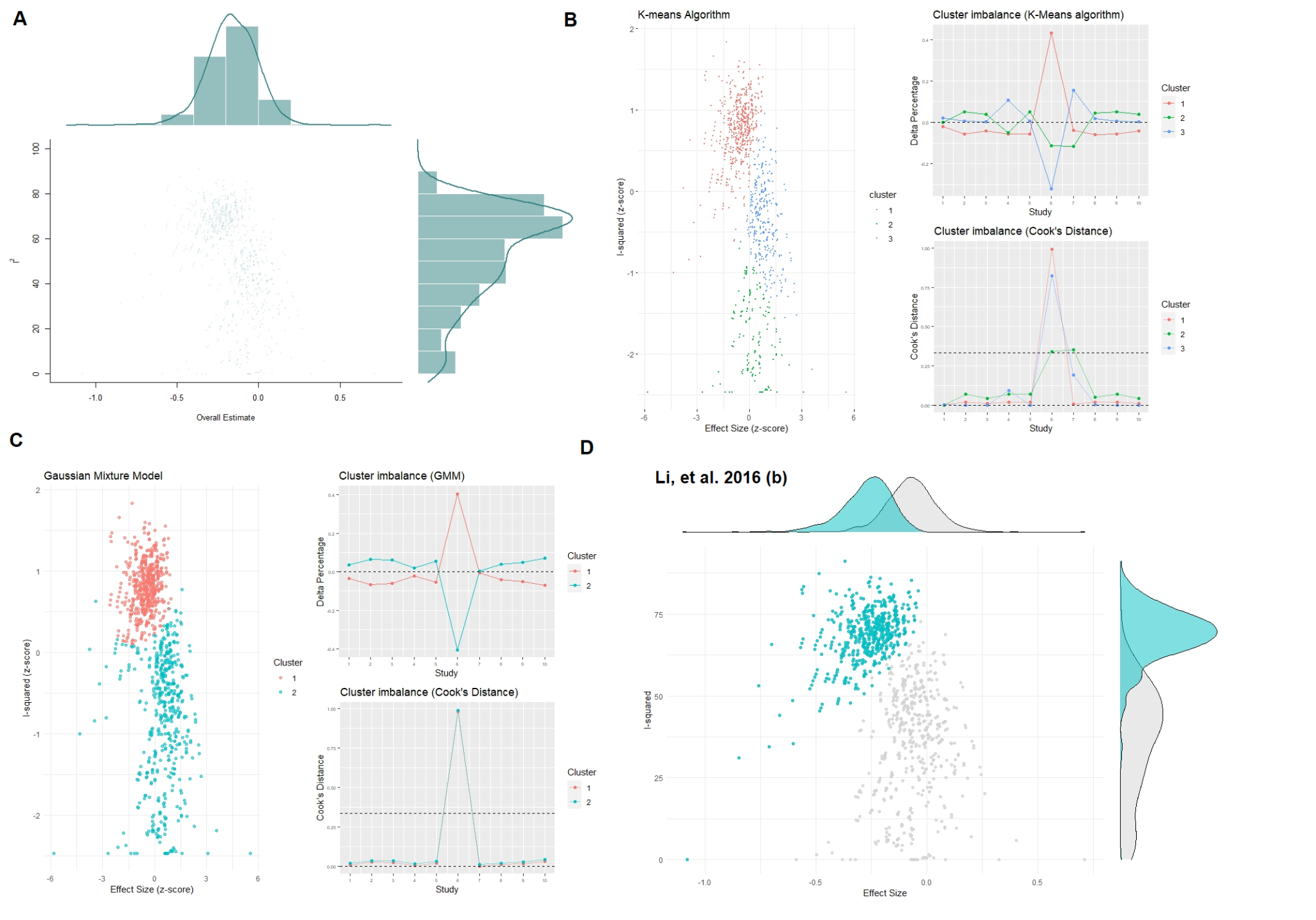

##### Subgroup Analyses

Supplementary Table 11. Subgroup analysis of the meta-analyses of all studies examining NAA levels in the basal ganglia of patients with cMDD compared to healthy controls.

|  | Subgroup sample size | Hedges’ g | 95% CI | p | I² | 95% CI | p subgroups |
| --- | --- | --- | --- | --- | --- | --- | --- |
| NAA quantification method | | | | | | | 0.753 |
| Cr scaling | 7 | -0.193 | -0.523, 0.138 | 0.253 | 58% | 3, 82% |  |
| Absolute | 3 | -0.071 | -0.755, 0.614 | 0.840 | 53% | 0, 87% |  |
| CSF correction | | | | | | | 0.093 |
| Yes | 5 | 0.030 | -0.160, 0.220 | 0.756 | 0% | 0, 54% |  |
| No | 5 | -0.395 | -0.853, 0.063 | 0.091 | 69% | 19, 88% |  |
| Field strength | | | | | | | 0.375 |
| 1.5 T | 6 | -0.052 | -0.386, 0.282 | 0.760 | 32% | 0, 73% |  |
| > 1.5 T | 4 | -0.325 | -0.829, 0.178 | 0.205 | 69% | 11, 89% |  |

##### Metaregressions

Supplementary Table 12. Results of the meta-regressions in the basal ganglia.

|  | beta | SE | p |
| --- | --- | --- | --- |
| Age | -0.9195 | 1.4103 | 0.5327 |
| Female % | -1.4563 | 2.5372 | 0.5818 |
| Illness duration | 0.0058 | 0.0271 | 0.835 |
| Ham-D scale | 15.5835 | 9.3816 | 0.156 |
| ^1^H-MRS field strength | -4.8517 | 11.8114 | 0.692 |
| TE | 0.0020 | 0.0027 | 0.468 |
| TR | 0.0058 | 0.0271 | 0.835 |

##### Publication bias

Supplementary Figure 43. Assessment of small sample publication bias with the contour-enhanced funnel plot for the studies examining NAA levels in the basal ganglia of cMDD.

The funnel plot showed an asymmetric pattern, with Ende et al. 2007 ^20^, the study having the smallest sample size (largest standard error), deviating toward large positive - albeit non-significant - effect sizes. There was a single study with highly significant effect size (p < 0.01), that is Li et al. 2016(b), which was indeed found to distort the pooled effect through influence analysis. Nonetheless, the Egger’s test yielded non-significant results (Intercept = -0.197, p = 0.941).

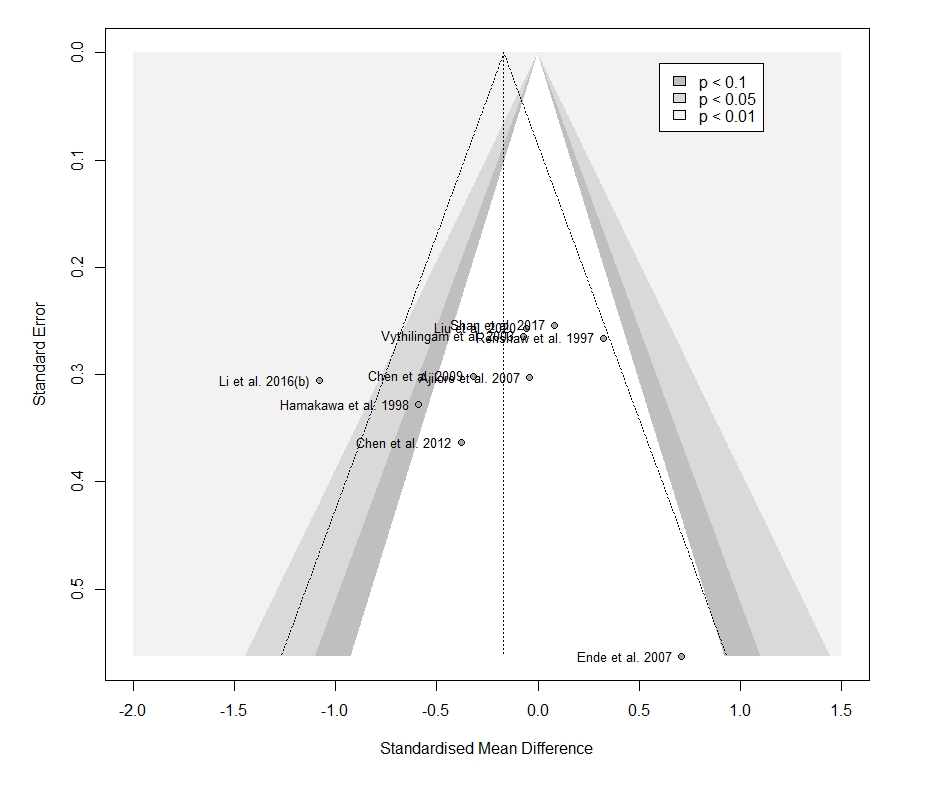

##### Sensitivity analysis on the influence of geriatric populations

The sensitivity analysis upon omission of the study assessing geriatric subjects ^21^ did not change the overall results (n = 9, Hedges’ g = -0.150, 95% CI -0.522 to 0.222; p = 0.379; Q = 18.19, *I²* = 56%, p = 0.020).

#### 13. Cerebellum

##### Main results

We included 3 studies: 78 patients and 68 controls.

Supplementary Figure 44. Forest plot of all studies examining NAA levels in the cerebellum of patients with cMDD compared to healthy controls.

The meta-analysis revealed no significant difference between patients and controls (n = 3, Hedges’ g = -0.033, 95% CI -0.655 to 0.588; p = 0.839; Q = 1.52, *I^2^=* 0%, p = 0.467). Positive values favour cMDD, while negative values favour controls.

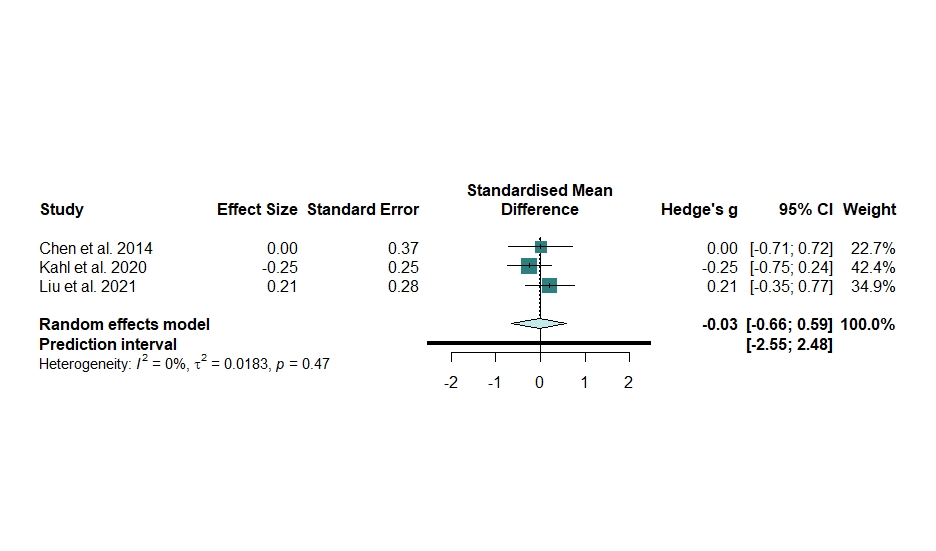

##### Heterogeneity

The between-study heterogeneity was not significant (Q = 1.52, *I^2^=* 0%, p = 0.467) without outliers, and no other analyses were performed because of the shortage of the studies (n = 3).

##### Publication bias

Supplementary Figure 45. Assessment of small sample publication bias with the contour-enhanced funnel plot for the studies examining NAA levels in the cerebellum of cMDD.

We found no evidence of publication bias. Egger’s test was not performed because there were not enough studies.

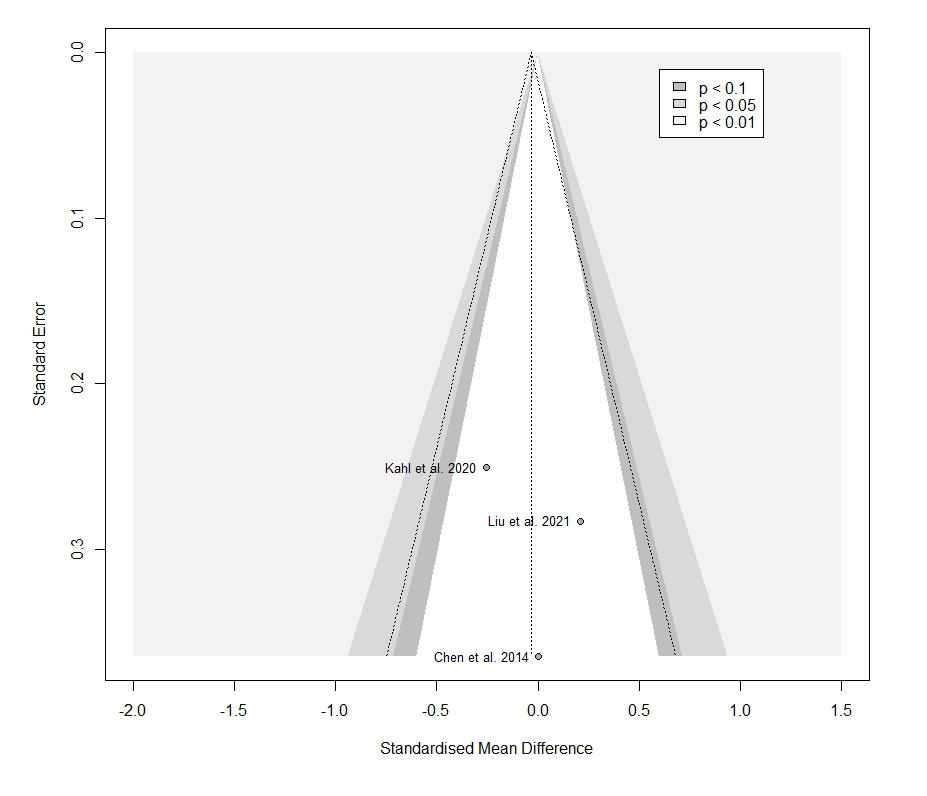

#### 14. White matter: Frontal white matter

##### Main results

We included 6 studies: 144 patients and 105 controls.

Supplementary Figure 46. Forest plot of all studies examining NAA levels in the frontal white matter of patients with cMDD compared to healthy controls.

The meta-analysis revealed a significant difference between patients and controls (n = 6, Hedges’ g = -0.471, 95% CI -0.891 to -0.052; p = 0.034; Q = 7.71, *I^2^=* 35.1%, p = 0.173). Positive values favour cMDD, while negative values favour controls.

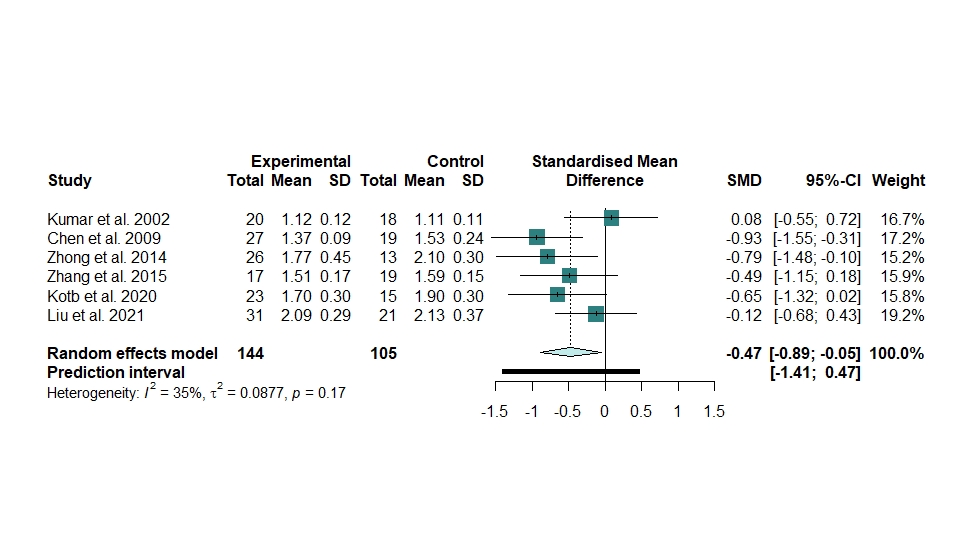

##### Heterogeneity

The between-study was not significant (Q = 7.71, *I^2^=* 35.1%, p = 0.173. No outliers were detected.

Supplementary Figure 47. Heterogeneity assessment in the frontal white matter meta-analysis

**A**. Baujat plot of the contribution of each study to the overall heterogeneity (as measured by Cochran’s Q) and its influence on the pooled effect size. **B**. Influence analysis with the *leave-one-out* method of the meta-analysis of all studies of cMDD examining NAA levels in the frontal white matter. Parameters of the influence analysis: standardized residuals, dffits, Cook’s distance, covariance ratio, tau^2^, Q, hat, and weight. No studies were identified as influential according to the cut-offs proposed by Viechtbauer and Cheung, 2010 ^8^. **C.** Forest plot of the overall effect sizes of the meta-analyses of all studies of cMDD examining NAA levels in the frontal white matter, recalculated with the *leave-one-out* method, ordered by effect size. **D.** Forest plot of the overall effect sizes of the meta-analyses of all studies of cMDD examining NAA levels in the frontal white matter, recalculated with the *leave-one-out* method, ordered by heterogeneity.

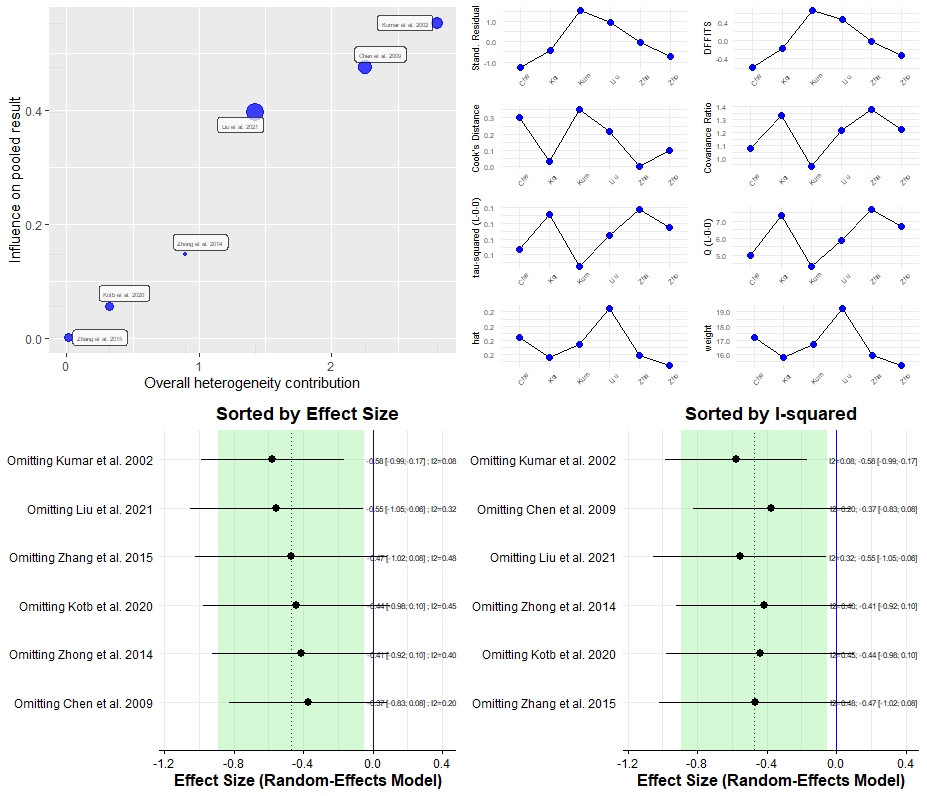

##### Publication bias

Supplementary Figure 48. Assessment of small sample publication bias with the contour-enhanced funnel plot for the studies examining NAA levels in the frontal white matter of cMDD.

Egger’s test could not be performed because there were not enough studies.

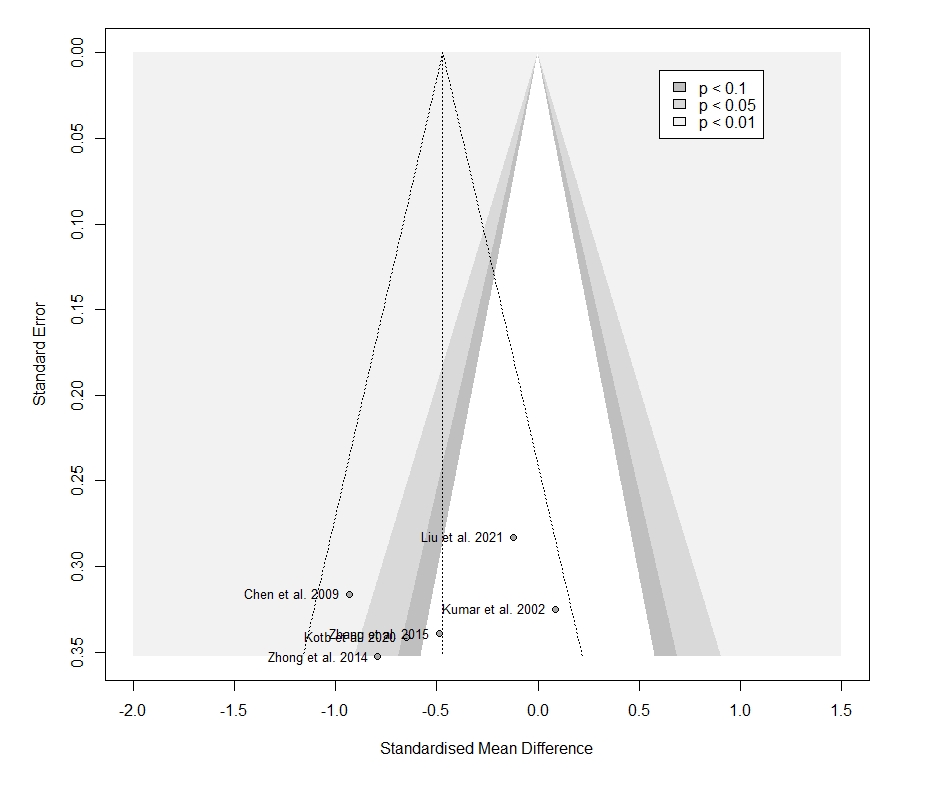

##### Sensitivity analysis on the influence of geriatric populations

The difference between cMDD patients and healthy controls in the NAA levels in the frontal white matter remained significant one reanalysed after the removal of studies conducted on geriatric populations ^12, 21^, with unchanged effect size (n = 4, Hedges’ g = -0.482, 95% CI -0.962 to -0.001; p = 0.050) and reduced between-study heterogeneity (Q = 2.65, *I^2^=* 0%, p = 0.449).

#### 15. White matter: Periventricular white matter

##### Main results

We included 3 studies: 53 patients and 39 controls.

Supplementary Figure 49. Forest plot of all studies examining NAA levels in the periventricular white matter of patients with cMDD compared to healthy controls.

The meta-analysis revealed a significant difference between patients and controls (n = 3, Hedges’ g = -0.4781, 95% CI -0.938 to -0.018; p = 0.047; Q = 0.49, *I^2^=* 0%, p = 0.784). Positive values favour cMDD, while negative values favour controls.

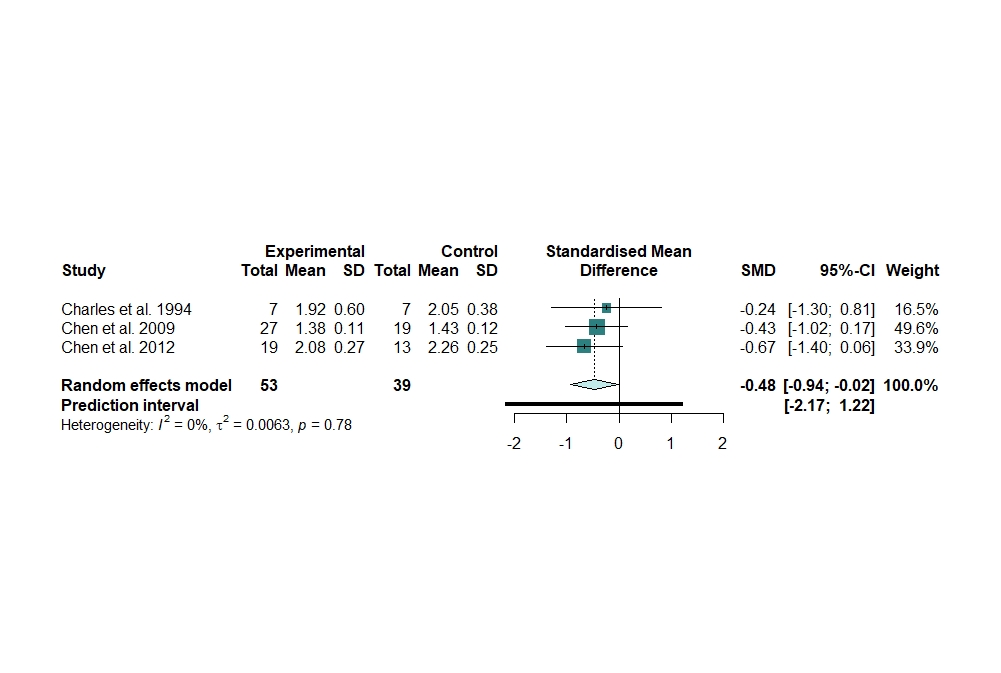

##### Publication bias

Supplementary Figure 50. Assessment of small sample publication bias with the contour-enhanced funnel plot for the studies examining NAA levels in the periventricular white matter of cMDD.

We found no evidence of publication bias. Egger’s test was not performed because there were not enough studies.

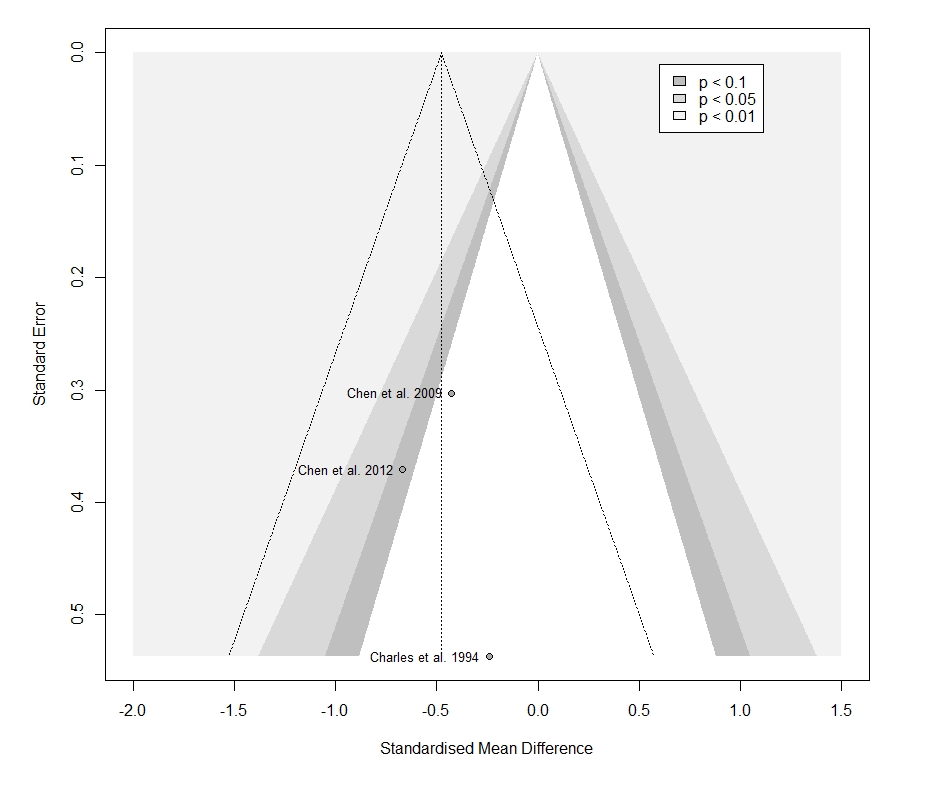

#### Sensitivity Analysis for the effect of Antidepressants in frontal lobe and basal ganglia

Supplementary Table 13. Sensitivity Analysis for the effect of Antidepressants in frontal lobe and basal ganglia in cMDD patients.

|  |  | Hedges’ g | 95% CI | p | I² | 95% CI | p subgroups |
| --- | --- | --- | --- | --- | --- | --- | --- |
| Frontal lobe | Antidepressant | | | | | | 0.900 |
|  | Yes | -0.323 | -1.068, 0.423 | 0.396 | 88% | 78, 94% |  |
|  | No | -0.373 | -0.636, -0.110 | 0.005 | 67% | 45, 80% |  |
| Basal ganglia | Antidepressant | | | | | | 0.990 |
|  | Yes | -0.163 | -0.630, 0.304 | 0.495 | 32 | 0, 76% |  |
|  | No | -0.168 | -0.765, 0.429 | 0.582 | 78 | 39, 92 |  |

Supplementary Figure 51. Forest plot of subgroup analyses for the effect of antidepressants.

Subgroup analyses in frontal lobe (A) and basal ganglia (B).

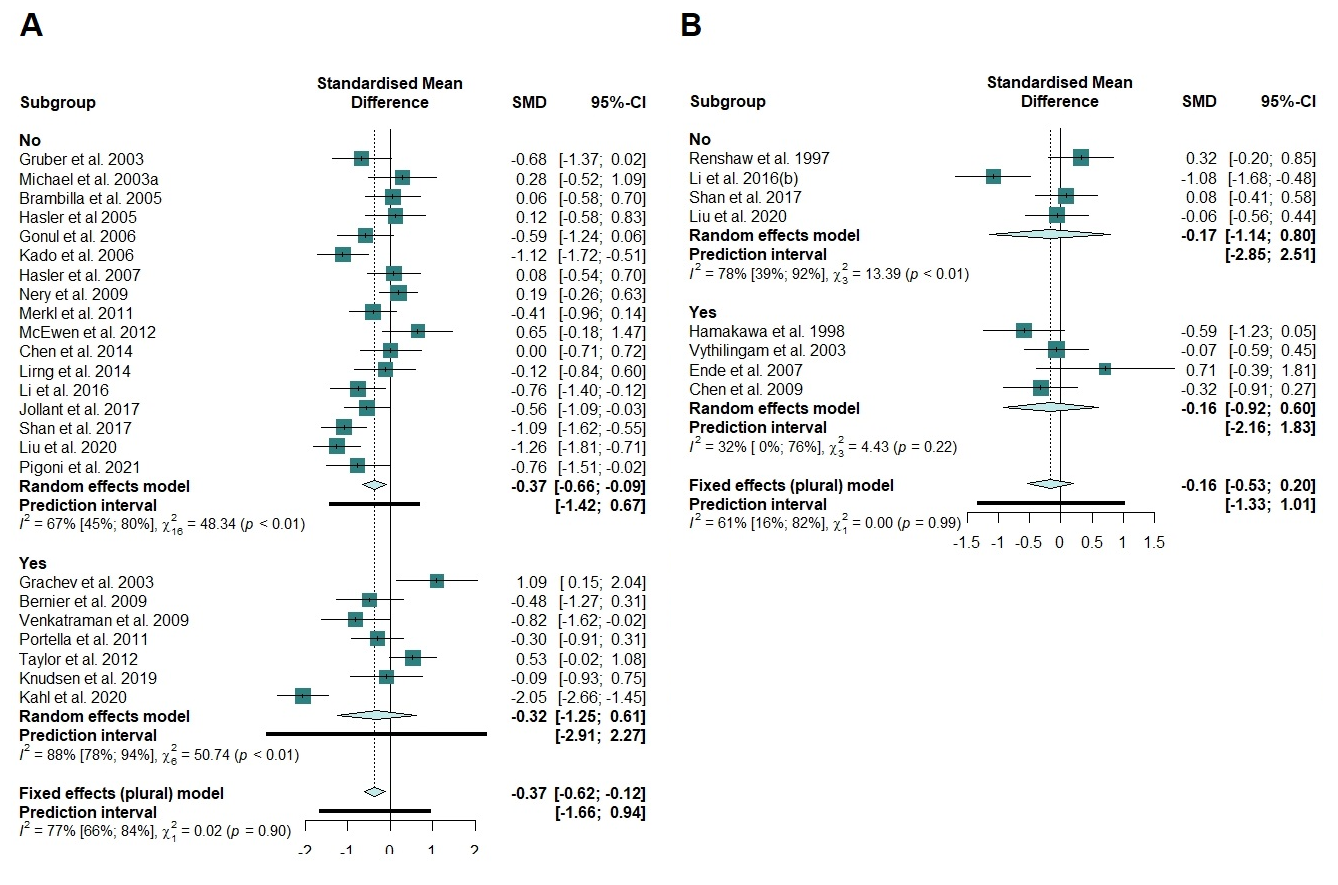

Supplementary Table 14. Quality assessment of the included studies with the Newcastle-Ottawa Quality Assessment Scale.

| **Study** | Case definition | Representativeness of cases | Selection of controls | Definition of controls | Comparability of cases and controls | Total score |
| --- | --- | --- | --- | --- | --- | --- |
| **^22^** | * | * | * | * | ** | 6 |
| **^23^** | * | * | * | * | ** | 6 |
| **^24^** | * | * | _ | * | ** | 5 |
| **^25^** | * | * | * | * | ** | 6 |
| **^26^** | * | * | * | * | ** | 6 |
| **^27^** | * | * | - | - | * | 3 |
| **^21^** | * | * | * | * | ** | 6 |
| **^28^** | * | * | _ | _ | * | 3 |
| **^29^** | * | * | * | _ | ** | 5 |
| **^30^** | * | * | * | * | ** | 6 |
| **^31^** | * | * | * | * | ** | 6 |
| **^20^** | * | * | _ | _ | ** | 4 |
| **^16^** | * | * | * | _ | * | 4 |
| **^32^** | * | * | * | * | ** | 6 |
| **^4^** | * | * | * | _ | ** | 5 |
| **^33^** | * | * | _ | * | ** | 5 |
| **^11^** | * | * | * | * | ** | 6 |
| **^34^** | * | * | * | * | * | 5 |
| **^35^** | * | * | * | * | ** | 6 |
| **^36^** | * | * | * | * | ** | 6 |
| **^17^** | * | * | _ | * | ** | 5 |
| **^37^** | * | * | * | * | ** | 6 |
| **^38^** | * | * | * | * | * | 5 |
| **^39^** | * | * | * | * | * | 5 |
| **^5^** | * | * | * | * | ** | 6 |
| **^40^** | * | * | * | * | ** | 6 |
| **^41^** | * | * | * | * | ** | 6 |
| **^14^** | * | * | _ | * | * | 4 |
| **^12^** | * | * | * | * | * | 5 |
| **^42^** | * | * | * | * | ** | 6 |
| **^43^** | * | * | * | * | ** | 6 |
| **^19^** | * | * | _ | * | ** | 5 |
| **^44^** | * | * | _ | * | ** | 5 |
| **^6^** | * | * | * | * | ** | 6 |
| **^45^** | * | * | * | * | ** | 6 |
| **^46^** | * | * | _ | _ | _ | 2 |
| **^47^** | * | * | * | * | ** | 6 |
| **^10^** | * | * | _ | * | * | 4 |
| **^48^** | * | * | * | * | * | 5 |
| **^49^** | * | * | * | * | ** | 6 |
| **^50^** | * | * | * | * | ** | 6 |
| **^51^** | * | * | * | _ | ** | 5 |
| **^52^** | * | * | * | * | ** | 6 |
| **^15^** | * | * | * | * | ** | 6 |
| **^53^** | * | * | * | * | ** | 6 |
| **^54^** | * | * | * | * | * | 5 |
| **^55^** | * | * | * | * | ** | 6 |
| **^56^** | * | * | * | * | ** | 6 |
| **^57^** | * | * | * | * | ** | 6 |
| **^58^** | * | * | _ | * | * | 4 |
| **^59^** | * | * | * | * | ** | 6 |
| **^13^** | * | * | * | * | ** | 6 |
| **^60^** | * | * | * | * | * | 6 |
| **^61^** | * | * | * | * | ** | 6 |
| **^7^** | * | * | * | * | ** | 6 |
| **^62^** | * | * | * | _ | ** | 4 |
| **^9^** | * | * | * | * | ** | 6 |
| **^18^** | * | * | * | * | ** | 6 |
| **^63^** | * | * | * | * | ** | 6 |
| **^64^** | * | * | * | * | ** | 6 |
| **^65^** | * | * | * | * | ** | 6 |
| **^66^** | * | * | * | * | ** | 6 |
| **^67^** | * | * | * | * | ** | 6 |

### REFERENCES

1.	Moriguchi S, Takamiya A, Noda Y, Horita N, Wada M, Tsugawa S *et al.* Glutamatergic neurometabolite levels in major depressive disorder: a systematic review and meta-analysis of proton magnetic resonance spectroscopy studies. *Mol Psychiatry* 2019; **24**(7)**:** 952-964.

2.	Olkin I, Dahabreh IJ, Trikalinos TA. GOSH - a graphical display of study heterogeneity. *Res Synth Methods* 2012; **3**(3)**:** 214-223.

3.	Fritz CO, Morris PE, Richler JJ. Effect size estimates: current use, calculations, and interpretation. *Journal of experimental psychology General* 2012; **141**(1)**:** 2-18.

4.	Grachev ID, Ramachandran TS, Thomas PS, Szeverenyi NM, Fredrickson BE. Association between dorsolateral prefrontal N-acetyl aspartate and depression in chronic back pain: an in vivo proton magnetic resonance spectroscopy study. *J Neural Transm (Vienna)* 2003; **110**(3)**:** 287-312.

5.	Kahl KG, Atalay S, Maudsley AA, Sheriff S, Cummings A, Frieling H *et al.* Altered neurometabolism in major depressive disorder: A whole brain (1)H-magnetic resonance spectroscopic imaging study at 3T. *Prog Neuropsychopharmacol Biol Psychiatry* 2020; **101:** 109916.

6.	Liu X, Zhong S, Li Z, Chen J, Wang Y, Lai S *et al.* Serum copper and zinc levels correlate with biochemical metabolite ratios in the prefrontal cortex and lentiform nucleus of patients with major depressive disorder. *Prog Neuropsychopharmacol Biol Psychiatry* 2020; **99:** 109828.

7.	Taylor MJ, Godlewska BR, Norbury R, Selvaraj S, Near J, Cowen PJ. Early increase in marker of neuronal integrity with antidepressant treatment of major depression: 1H-magnetic resonance spectroscopy of N-acetyl-aspartate. *Int J Neuropsychopharmacol* 2012; **15**(10)**:** 1541-1546.

8.	Viechtbauer W, Cheung MW. Outlier and influence diagnostics for meta-analysis. *Res Synth Methods* 2010; **1**(2)**:** 112-125.

9.	Venkatraman TN, Krishnan RR, Steffens DC, Song AW, Taylor WD. Biochemical abnormalities of the medial temporal lobe and medial prefrontal cortex in late-life depression. *Psychiatry Res* 2009; **172**(1)**:** 49-54.

10.	Merkl A, Schubert F, Quante A, Luborzewski A, Brakemeier EL, Grimm S *et al.* Abnormal cingulate and prefrontal cortical neurochemistry in major depression after electroconvulsive therapy. *Biol Psychiatry* 2011; **69**(8)**:** 772-779.

11.	Guo Z, Zhang J, Liu X, Hou H, Cao Y, Wei F *et al.* Neurometabolic characteristics in the anterior cingulate gyrus of Alzheimer's disease patients with depression: a (1)H magnetic resonance spectroscopy study. *BMC Psychiatry* 2015; **15:** 306.

12.	Kumar A, Thomas A, Lavretsky H, Yue K, Huda A, Curran J *et al.* Frontal white matter biochemical abnormalities in late-life major depression detected with proton magnetic resonance spectroscopy. *Am J Psychiatry* 2002; **159**(4)**:** 630-636.

13.	Smith GS, Oeltzschner G, Gould NF, Leoutsakos JS, Nassery N, Joo JH *et al.* Neurotransmitters and Neurometabolites in Late-Life Depression: A Preliminary Magnetic Resonance Spectroscopy Study at 7T. *J Affect Disord* 2021; **279:** 417-425.

14.	Kotb MA, Kamal AM, Aldossary NM. Value of magnetic resonance spectroscopy in geriatric patients with cognitive impairment. *Egypt J Neurol Psychiatry Neurosurg* 2020; **56, 10**.

15.	Njau S, Joshi SH, Espinoza R, Leaver AM, Vasavada M, Marquina A *et al.* Neurochemical correlates of rapid treatment response to electroconvulsive therapy in patients with major depression. *J Psychiatry Neurosci* 2017; **42**(1)**:** 6-16.

16.	Ezzati A, Zimmerman ME, Katz MJ, Lipton RB. Hippocampal correlates of depression in healthy elderly adults. *Hippocampus* 2013; **23**(12)**:** 1137-1142.

17.	Jayaweera HK, Lagopoulos J, Duffy SL, Lewis SJ, Hermens DF, Norrie L *et al.* Spectroscopic markers of memory impairment, symptom severity and age of onset in older people with lifetime depression: Discrete roles of N-acetyl aspartate and glutamate. *J Affect Disord* 2015; **183:** 31-38.

18.	Vythilingam M, Charles HC, Tupler LA, Blitchington T, Kelly L, Krishnan KR. Focal and lateralized subcortical abnormalities in unipolar major depressive disorder: an automated multivoxel proton magnetic resonance spectroscopy study. *Biol Psychiatry* 2003; **54**(7)**:** 744-750.

19.	Li Y, Jakary A, Gillung E, Eisendrath S, Nelson SJ, Mukherjee P *et al.* Evaluating metabolites in patients with major depressive disorder who received mindfulness-based cognitive therapy and healthy controls using short echo MRSI at 7 Tesla. *MAGMA* 2016; **29**(3)**:** 523-533.

20.	Ende G, Demirakca T, Walter S, Wokrina T, Sartorius A, Wildgruber D *et al.* Subcortical and medial temporal MR-detectable metabolite abnormalities in unipolar major depression. *Eur Arch Psychiatry Clin Neurosci* 2007; **257**(1)**:** 36-39.

21.	Chen CS, Chiang IC, Li CW, Lin WC, Lu CY, Hsieh TJ *et al.* Proton magnetic resonance spectroscopy of late-life major depressive disorder. *Psychiatry Res* 2009; **172**(3)**:** 210-214.

22.	Ajilore O, Haroon E, Kumaran S, Darwin C, Binesh N, Mintz J *et al.* Measurement of brain metabolites in patients with type 2 diabetes and major depression using proton magnetic resonance spectroscopy. *Neuropsychopharmacology* 2007; **32**(6)**:** 1224-1231.

23.	Auer DP, Putz B, Kraft E, Lipinski B, Schill J, Holsboer F. Reduced glutamate in the anterior cingulate cortex in depression: an in vivo proton magnetic resonance spectroscopy study. *Biol Psychiatry* 2000; **47**(4)**:** 305-313.

24.	Bernier D, Bartha R, Devarajan S, Macmaster FP, Schmidt MH, Rusak B. Effects of overnight sleep restriction on brain chemistry and mood in women with unipolar depression and healthy controls. *J Psychiatry Neurosci* 2009; **34**(5)**:** 352-360.

25.	Bhagwagar Z, Wylezinska M, Jezzard P, Evans J, Ashworth F, Sule A *et al.* Reduction in occipital cortex gamma-aminobutyric acid concentrations in medication-free recovered unipolar depressed and bipolar subjects. *Biol Psychiatry* 2007; **61**(6)**:** 806-812.

26.	Brambilla P, Stanley JA, Nicoletti MA, Sassi RB, Mallinger AG, Frank E *et al.* 1H Magnetic resonance spectroscopy study of dorsolateral prefrontal cortex in unipolar mood disorder patients. *Psychiatry Res* 2005; **138**(2)**:** 131-139.

27.	Charles HC, Lazeyras F, Krishnan KR, Boyko OB, Payne M, Moore D. Brain choline in depression: in vivo detection of potential pharmacodynamic effects of antidepressant therapy using hydrogen localized spectroscopy. *Prog Neuropsychopharmacol Biol Psychiatry* 1994; **18**(7)**:** 1121-1127.

28.	Chen S, Lai L, Kang Z, Luo X, Zhang J, Li J. Imaging changes in neural circuits in patients with depression using (1)H-magnetic resonance spectroscopy and diffusion tensor imaging. *Neural Regen Res* 2012; **7**(24)**:** 1881-1888.

29.	Chen LP, Dai HY, Dai ZZ, Xu CT, Wu RH. Anterior cingulate cortex and cerebellar hemisphere neurometabolite changes in depression treatment: A 1H magnetic resonance spectroscopy study. *Psychiatry Clin Neurosci* 2014; **68**(5)**:** 357-364.

30.	Coupland NJ, Ogilvie CJ, Hegadoren KM, Seres P, Hanstock CC, Allen PS. Decreased prefrontal Myo-inositol in major depressive disorder. *Biol Psychiatry* 2005; **57**(12)**:** 1526-1534.

31.	de Diego-Adelino J, Portella MJ, Gomez-Anson B, Lopez-Moruelo O, Serra-Blasco M, Vives Y *et al.* Hippocampal abnormalities of glutamate/glutamine, N-acetylaspartate and choline in patients with depression are related to past illness burden. *J Psychiatry Neurosci* 2013; **38**(2)**:** 107-116.

32.	Gonul AS, Kitis O, Ozan E, Akdeniz F, Eker C, Eker OD *et al.* The effect of antidepressant treatment on N-acetyl aspartate levels of medial frontal cortex in drug-free depressed patients. *Prog Neuropsychopharmacol Biol Psychiatry* 2006; **30**(1)**:** 120-125.

33.	Gruber S, Frey R, Mlynarik V, Stadlbauer A, Heiden A, Kasper S *et al.* Quantification of metabolic differences in the frontal brain of depressive patients and controls obtained by 1H-MRS at 3 Tesla. *Invest Radiol* 2003; **38**(7)**:** 403-408.

34.	Hamakawa H, Kato T, Murashita J, Kato N. Quantitative proton magnetic resonance spectroscopy of the basal ganglia in patients with affective disorders. *Eur Arch Psychiatry Clin Neurosci* 1998; **248**(1)**:** 53-58.

35.	Hasler G, Neumeister A, van der Veen JW, Tumonis T, Bain EE, Shen J *et al.* Normal prefrontal gamma-aminobutyric acid levels in remitted depressed subjects determined by proton magnetic resonance spectroscopy. *Biol Psychiatry* 2005; **58**(12)**:** 969-973.

36.	Hasler G, van der Veen JW, Tumonis T, Meyers N, Shen J, Drevets WC. Reduced prefrontal glutamate/glutamine and gamma-aminobutyric acid levels in major depression determined using proton magnetic resonance spectroscopy. *Arch Gen Psychiatry* 2007; **64**(2)**:** 193-200.

37.	Jia Y, Zhong S, Wang Y, Liu T, Liao X, Huang L. The correlation between biochemical abnormalities in frontal white matter, hippocampus and serum thyroid hormone levels in first-episode patients with major depressive disorder. *J Affect Disord* 2015; **180:** 162-169.

38.	Jollant F, Near J, Turecki G, Richard-Devantoy S. Spectroscopy markers of suicidal risk and mental pain in depressed patients. *Prog Neuropsychopharmacol Biol Psychiatry* 2016.

39.	Kado H, Kimura H, Murata T, Nagata K, Kanno I. Depressive psychosis: clinical usefulness of MR spectroscopy data in predicting prognosis. *Radiology* 2006; **238**(1)**:** 248-255.

40.	Kaymak SU, Demir B, Oguz KK, Senturk S, Ulug B. Antidepressant effect detected on proton magnetic resonance spectroscopy in drug-naive female patients with first-episode major depression. *Psychiatry Clin Neurosci* 2009; **63**(3)**:** 350-356.

41.	Knudsen MK, Near J, Blicher AB, Videbech P, Blicher JU. Magnetic resonance (MR) spectroscopic measurement of gamma-aminobutyric acid (GABA) in major depression before and after electroconvulsive therapy. *Acta Neuropsychiatr* 2019; **31**(1)**:** 17-26.

42.	Li M, Metzger CD, Li W, Safron A, van Tol MJ, Lord A *et al.* Dissociation of glutamate and cortical thickness is restricted to regions subserving trait but not state markers in major depressive disorder. *J Affect Disord* 2014; **169:** 91-100.

43.	Li H, Xu H, Zhang Y, Guan J, Zhang J, Xu C *et al.* Differential neurometabolite alterations in brains of medication-free individuals with bipolar disorder and those with unipolar depression: a two-dimensional proton magnetic resonance spectroscopy study. *Bipolar Disord* 2016; **18**(7)**:** 583-590.

44.	Lirng JF, Chen HC, Fuh JL, Tsai CF, Liang JF, Wang SJ. Increased myo-inositol level in dorsolateral prefrontal cortex in migraine patients with major depression. *Cephalalgia* 2015; **35**(8)**:** 702-709.

45.	Liu X, Zhong S, Yan L, Zhao H, Wang Y, Hu Y *et al.* Correlations Among mRNA Expression Levels of ATP7A, Serum Ceruloplasmin Levels, and Neuronal Metabolism in Unmedicated Major Depressive Disorder. *Int J Neuropsychopharmacol* 2020; **23**(10)**:** 642-652.

46.	Martinez-Bisbal MC, Arana E, Marti-Bonmati L, Molla E, Celda B. Cognitive impairment: classification by 1H magnetic resonance spectroscopy. *Eur J Neurol* 2004; **11**(3)**:** 187-193.

47.	McEwen AM, Burgess DT, Hanstock CC, Seres P, Khalili P, Newman SC *et al.* Increased glutamate levels in the medial prefrontal cortex in patients with postpartum depression. *Neuropsychopharmacology* 2012; **37**(11)**:** 2428-2435.

48.	Mervaala E, Fohr J, Kononen M, Valkonen-Korhonen M, Vainio P, Partanen K *et al.* Quantitative MRI of the hippocampus and amygdala in severe depression. *Psychol Med* 2000; **30**(1)**:** 117-125.

49.	Michael N, Erfurth A, Ohrmann P, Arolt V, Heindel W, Pfleiderer B. Metabolic changes within the left dorsolateral prefrontal cortex occurring with electroconvulsive therapy in patients with treatment resistant unipolar depression. *Psychol Med* 2003; **33**(7)**:** 1277-1284.

50.	Milne A, MacQueen GM, Yucel K, Soreni N, Hall GB. Hippocampal metabolic abnormalities at first onset and with recurrent episodes of a major depressive disorder: a proton magnetic resonance spectroscopy study. *Neuroimage* 2009; **47**(1)**:** 36-41.

51.	Mohamed MA, Smith MA, Schlund MW, Nestadt G, Barker PB, Hoehn-Saric R. Proton magnetic resonance spectroscopy in obsessive-compulsive disorder: a pilot investigation comparing treatment responders and non-responders. *Psychiatry Res* 2007; **156**(2)**:** 175-179.

52.	Nery FG, Stanley JA, Chen HH, Hatch JP, Nicoletti MA, Monkul ES *et al.* Normal metabolite levels in the left dorsolateral prefrontal cortex of unmedicated major depressive disorder patients: a single voxel (1)H spectroscopy study. *Psychiatry Res* 2009; **174**(3)**:** 177-183.

53.	Pfleiderer B, Michael N, Erfurth A, Ohrmann P, Hohmann U, Wolgast M *et al.* Effective electroconvulsive therapy reverses glutamate/glutamine deficit in the left anterior cingulum of unipolar depressed patients. *Psychiatry Res* 2003; **122**(3)**:** 185-192.

54.	Pigoni A, Delvecchio G, Squarcina L, Bonivento C, Girardi P, Finos L *et al.* Sex differences in brain metabolites in anxiety and mood disorders. *Psychiatry Res Neuroimaging* 2020; **305:** 111196.

55.	Portella MJ, de Diego-Adelino J, Gomez-Anson B, Morgan-Ferrando R, Vives Y, Puigdemont D *et al.* Ventromedial prefrontal spectroscopic abnormalities over the course of depression: a comparison among first episode, remitted recurrent and chronic patients. *J Psychiatr Res* 2011; **45**(4)**:** 427-434.

56.	Renshaw PF, Lafer B, Babb SM, Fava M, Stoll AL, Christensen JD *et al.* Basal ganglia choline levels in depression and response to fluoxetine treatment: an in vivo proton magnetic resonance spectroscopy study. *Biol Psychiatry* 1997; **41**(8)**:** 837-843.

57.	Rosa CE, Soares JC, Figueiredo FP, Cavalli RC, Barbieri MA, Schaufelberger MS *et al.* Glutamatergic and neural dysfunction in postpartum depression using magnetic resonance spectroscopy. *Psychiatry Res Neuroimaging* 2017; **265:** 18-25.

58.	Sanacora G, Gueorguieva R, Epperson CN, Wu YT, Appel M, Rothman DL *et al.* Subtype-specific alterations of gamma-aminobutyric acid and glutamate in patients with major depression. *Arch Gen Psychiatry* 2004; **61**(7)**:** 705-713.

59.	Shan Y, Jia Y, Zhong S, Li X, Zhao H, Chen J *et al.* Correlations between working memory impairment and neurometabolites of prefrontal cortex and lenticular nucleus in patients with major depressive disorder. *J Affect Disord* 2018; **227:** 236-242.

60.	Sozeri-Varma G, Kalkan-Oguzhanoglu N, Efe M, Kiroglu Y, Duman T. Neurochemical metabolites in prefrontal cortex in patients with mild/moderate levels in first-episode depression. *Neuropsychiatr Dis Treat* 2013; **9:** 1053-1059.

61.	Tae WS, Kim SS, Lee KU, Nam EC, Koh SH. Progressive decrease of N-acetylaspartate to total creatine ratio in the pregenual anterior cingulate cortex in patients with major depressive disorder: longitudinal 1H-MR spectroscopy study. *Acta Radiol* 2014; **55**(5)**:** 594-603.

62.	Tosun S, Tosun M, Akansel G, Gokbakan AM, Unver H, Tural U. Proton magnetic resonance spectroscopic analysis of changes in brain metabolites following electroconvulsive therapy in patients with major depressive disorder. *Int J Psychiatry Clin Pract* 2020; **24**(1)**:** 96-101.

63.	Wang Y, Jia Y, Xu G, Ling X, Liu S, Huang L. Frontal white matter biochemical abnormalities in first-episode, treatment-naive patients with major depressive disorder: a proton magnetic resonance spectroscopy study. *J Affect Disord* 2012; **136**(3)**:** 620-626.

64.	Zavorotnyy M, Zollner R, Rekate H, Dietsche P, Bopp M, Sommer J *et al.* Intermittent theta-burst stimulation moderates interaction between increment of N-Acetyl-Aspartate in anterior cingulate and improvement of unipolar depression. *Brain Stimul* 2020; **13**(4)**:** 943-952.

65.	Zhang Y, Han Y, Wang Y, Zhang Y, Li L, Jin E *et al.* A MRS study of metabolic alterations in the frontal white matter of major depressive disorder patients with the treatment of SSRIs. *BMC Psychiatry* 2015; **15:** 99.

66.	Zhong S, Wang Y, Zhao G, Xiang Q, Ling X, Liu S *et al.* Similarities of biochemical abnormalities between major depressive disorder and bipolar depression: a proton magnetic resonance spectroscopy study. *J Affect Disord* 2014; **168:** 380-386.

67.	Sendur I, Kalkan Oguzhanoglu N, Sozeri Varma G. Study on Dorsolateral Prefrontal Cortex Neurochemical Metabolite Levels of Patients with Major Depression Using H-MRS Technique. *Turk Psikiyatri Derg* 2020; **31**(2)**:** 75-83.
